# Aetiology of vaginal discharge, urethral discharge, and genital ulcer in sub-Saharan Africa: systematic review and meta-regression

**DOI:** 10.1101/2023.11.09.23298288

**Authors:** Julia Michalow, Magdalene K Walters, Olanrewaju Edun, Max Wybrant, Bethan Davies, Tendesayi Kufa, Thabitha Mathega, Sungai T Chabata, Frances M Cowan, Anne Cori, Marie-Claude Boily, Jeffrey W Imai-Eaton

## Abstract

**Introduction:** Syndromic management is widely used to treat symptomatic sexually transmitted infections in settings lacking aetiologic diagnostics. However, heterogeneity in underlying aetiologies and consequent treatment suitability are uncertain without regular assessment. This systematic review characterised aetiologies for vaginal discharge, urethral discharge, and genital ulcer in sub-Saharan Africa (SSA).

**Methods:** We searched Embase, MEDLINE, Global Health, and Web of Science until July 25, 2022, and grey literature until August 31, 2022, for studies reporting aetiologic diagnoses among symptomatic populations in SSA. We adjusted observations for diagnostic test performance and used generalised linear mixed-effects meta-regressions to estimate aetiologic distributions, trends, and determinants.

**Results:** Of 4136 identified records, 198 reports were included from 183 studies in 32 countries between 1969 and 2022. In 2015, primary aetiologies for vaginal discharge were candidiasis (69.4% [95% CI:44.1-86.6%], n=50), bacterial vaginosis (50.0% [32.3-67.8%], n=39), chlamydia (16.5% [8.7-29.0%], n=49), and trichomoniasis (12.9% [7.7-20.7%], n=78); for urethral discharge were gonorrhoea (78.8% [70.9-85.1%], n=67) and chlamydia (22.2% [16.0-30.1%], n=48); and for genital ulcer were HSV-2 (56.1% [39.2-71.6%], n=46) and syphilis (7.8% [5.3-11.4%], n=115). Regional variation was marginal. Temporal variation was substantial, particularly for genital ulcer. For each symptom, HIV-status and age were significantly associated with infection diagnoses, although aetiologic hierarchies were largely the same by strata.

**Conclusion:** Syndrome aetiologies in SSA align with WHO guidelines without strong evidence of contextual or demographic variation, supporting broad guideline applicability. Temporal changes underscore the need for aetiologic re-assessment. STI surveillance using syndrome-based assessments is noncomprehensive and requires studies among symptomatic and asymptomatic populations.

**PROSPERO number:** CRD42022348045

## Introduction

In 2020, the World Health Organization (WHO) estimated 374 million new infections worldwide of the four most common curable sexually transmitted infections (STIs): chlamydia, gonorrhoea, syphilis, and trichomoniasis.^1^ However, in sub-Saharan Africa (SSA), which accounts for 40% of the global STI burden,^2^ access to laboratory or point-of-care aetiologic diagnostics is limited. Syndromic case management, in which probable causative infection(s) are treated based on presenting symptoms, was introduced by WHO in 1984 and remains the standard of care.^3^ The approach enables rapid treatment but has several limitations. Many STIs evade treatment due to high rates of asymptomatic infection, particularly in women.^4^ The diagnostic accuracy of these algorithms is suboptimal, despite efforts to incorporate evolving STI epidemiology.^5^ Furthermore, WHO guidance recommends national re-assessment of syndrome aetiologies every 2 years to ensure the relevance of syndromic algorithms,^6,7^ yet approximately 40% of African countries (11/26 in WHO survey) reported including these assessments in STI surveillance.^8^ These factors collectively may lead to unnecessary, incorrect, and missed treatment for STIs, which is particularly concerning amid rising antimicrobial resistance.^5^

In 2021, the WHO released new guidelines for symptomatic STI management, the first update since 2003.^3^ These updates were informed by global systematic reviews of studies assessing the diagnostic performance of different syndromic management algorithms. Although the guidelines accounted for broad changes over time in the underlying causes of each syndrome, they lacked a comprehensive review of the distribution of STIs among symptomatic populations. This information would support future modification of syndromic management protocols to align with local epidemiology.

The objective of our study was to characterise the aetiologies for three prevalent STI symptoms in SSA: vaginal discharge, urethral discharge, and genital ulcer. We performed a systematic review and meta-analysis to estimate the distribution of aetiologies for each symptom, investigate their spatiotemporal changes, and evaluate variation according to population-specific determinants, namely sex, HIV-status, and age.

## Methods

### Data sources and search strategy

We systematically searched for studies assessing the aetiology of vaginal discharge, urethral discharge, and genital ulcer in sub-Saharan Africa. Embase (Ovid), MEDLINE (Ovid), Global Health (Ovid) and Web of Science were searched from inception to 25 July 2022. Search term domains included relevant terms and synonyms for “symptoms”, “infections” and “sub-Saharan Africa” (Table S1). We also performed a comprehensive grey literature search up to 31 August 2022 using the same search term domains. Sources included websites and reports by the WHO, UNAIDS, and Ministries of Health, and conference abstracts published between 2000 and 2022 from the STI & HIV World Congress, International AIDS Society, and International Conference on AIDS and STIs in Africa. We contacted authors of relevant conference abstracts to enquire about potential unpublished data. Sub-Saharan Africa and its sub-regions were defined according to the UN M49 Standard (Table S2).^9^

### Study selection and eligibility criteria

Search results were uploaded and de-duplicated in Covidence systematic review software (Veritas Health Innovation, Melbourne, Australia). Two reviewers independently screened titles and abstract records for eligibility, and then assessed full text reports for inclusion. Any discrepancies were resolved through consensus or by a third reviewer.

We included reports that contained empirical data on the proportion of women with vaginal discharge (VD) diagnosed with bacterial vaginosis (BV), any *Candida* species (CS), *Candida albicans* (CA), *Chlamydia trachomatis* (CT), *Mycoplasma genitalium* (MG), *Neisseria gonorrhoeae* (NG), *Trichomonas vaginalis* (TV), or unknown aetiology (negative aetiologic test results); proportion of men with urethral discharge (UD) diagnosed with CT, MG, NG, TV, or unknown aetiology; and proportion of men and women with genital ulcer (GU) diagnosed with *Haemophilus ducreyi* (HD), herpes simplex virus of unspecified type (HSV) or types 1 or 2 (HSV-1 or HSV-2), lymphogranuloma venereum (LGV) caused by CT serovars L1–L3, *Treponema pallidum* (TP), or unknown aetiology. Since BV and candidiasis are not considered STIs, but are common causes of vaginal discharge, we refer to the broader term reproductive tract infections (RTIs) in this study.

Studies were included if: (1) participants were symptomatic at the time of testing, defined by the presence of either self-reported or clinician-evaluated abnormal vaginal discharge, urethral discharge, or genital ulcer, (2) participants were aged 10 years and older, (3) the sample size was at least 10, and (4) the diagnostic methodology for each infection was described and assessed as valid according to published recommendations.^3,10–13^ Exclusion criteria were: (1) qualitative studies, case reports, commentaries, reviews, mathematical modelling studies, and longitudinal and randomised controlled studies reporting outcomes post baseline only, and (2) studies published in languages other than English, French, or Portuguese.

### Data extraction

Data were independently double extracted from each study (number of studies denoted as N) with discrepancies resolved through consensus or by a third reviewer. A study observation (number of observations denoted as n) was the proportion of symptomatic individuals diagnosed with a given RTI or, if not directly reported, the numerator and denominator to calculate the proportion. In cases of RTI co-infection, we extracted each RTI separately, potentially causing the total infected proportion to exceed 1 when aggregating all observations for a given population. Observations were extracted with stratification by population type (symptomatic clinic attendee; general population; higher-risk general population, such as truck drivers, mineworkers, soldiers, bar workers; or key population), country, year or time-period if not stratified by year, sex, HIV status, and age group as available. We excluded strata subsamples of fewer than 10 participants. Data were also extracted on study characteristics, participant characteristics, and diagnostic methods (Table S3). We also prepared and extracted unpublished data from two databases identified during the search (Text S1, Table S4).

If multiple reports included the same outcome(s) for a study, we preferentially retained observations from the largest sample or, if samples were the same size, observations from the report with the largest number of RTIs tested. If the sample sizes and number of RTIs were equal, the most recent report was retained. Where possible, we preferentially used observations tabulated directly from databases and excluded corresponding published articles.

If a study reported outcomes for self-reported and clinician-evaluated symptoms in the same population, we preferentially extracted observations for the latter. If multiple diagnostic tests were conducted, outcomes based on the most accurate test method for the pathogen and symptom (Tables S5, S6, S7) were preferentially extracted. When multiple diagnostic tests were used for syphilis among those with genital ulcer, we preferentially extracted observations for tests using ulcer swab specimens over serology.^3^ When syphilis serology was used, we prioritised observations from the combination sof non-treponemal and treponemal tests when available. “Unknown aetiology” was only extracted from studies with observations for three or more infectious pathogens, and not from studies testing for fewer pathogens.

### Data analysis

To adjust reported proportions for diagnostic test performance, we classified diagnostic tests into broad categories and compiled sensitivity and specificity estimates for each test category from literature, with priority given to chsaracteristics published by the WHO (Text S2, Tables S5, S6, S7).^10,14^ We used a Bayesian approach to estimate the true proportion of symptomatic individuals with a given RTI.^15,16^ Adjusted numerators and denominators were calculated based on the mean and standard error of the true proportion and used to pool observations (Text S2).

Estimates of regional trends in the diagnosed proportion for each infection used observations among adults of mixed or unmeasured HIV status. We performed meta-regressions for each symptom using generalised linear mixed-effects models.^17^ Models included fixed effects for RTI, the interaction of RTI and year (midpoint date of data collection measured as continuous calendar year), and the interaction of RTI and sex (genital ulcer only). Models included study random intercepts and random intercepts and slopes per year for the crossed interaction of RTI and region (central and western, eastern, or southern Africa). We weighted pooled regional means by sex-matched regional population estimates for adults 15 years and older in 2015 from the UN World Population Prospects 2022.^18^

To assess the effects of HIV status and age, we used observations with appropriate stratification. We extended the meta-regressions described above to include fixed effects for either HIV status (HIV-positive, HIV-negative) or age group (<25 years, ≥25 years), but did not include random slopes due to their low standard deviation in the main analysis.

### Risk of bias assessment and sensitivity analysis

Risk of bias for included studies was assessed by modifying the Joanna Briggs Institute critical appraisal tool for prevalence studies (Table S8).^19^ Each of 10 criteria were independently double assessed, with discrepancies resolved through consensus or by a third reviewer. Each study received a total score based on the number of criteria met, and was considered as higher (4 points and below), moderate (5-7) or lower (8-10) risk of bias. We used the meta-regressions described above to compare trends in the diagnosed proportion for each infection among adults of mixed or unmeasured HIV status when alternatively including studies of lower risk only, lower and moderate risk, and any risk of bias.

To assess sensitivity to the diagnostic test adjustments, we compared trend estimates between models using observations as reported and models using observations adjusted for diagnostic test performance. This comparison was conducted for all diagnostic test types (all RTIs) and specifically for nucleic acid amplification testing (NAAT) (all RTIs, excluding BV, CA, and CS).

Unless stated otherwise, all analyses use observations adjusted for diagnostic test performance from studies of all risk of bias levels. In analyses including studies across multiple regions without stratification, studies were classified using the region where most participants were recruited from. Pooled results are reported as meta-regression model predictions for the year 2015, representing the most recent quinquennium within the timespan of substantial available data. Results are presented as means with 95% confidence intervals (95% CIs). Analyses were conducted in R version 4.2.3, using rstan version 2.26.21 and glmmTMB version 1.1.7.^20,21^

This systematic review and meta-analysis was pre-registered on PROSPERO (CRD: 42022348045)^22^ and reported according to PRISMA guidelines (Table S9).^23^ The study protocol was reviewed and approved by the Imperial College Research Ethics Committee (ICREC #6389606).

## Results

### Search results and scope

We identified 7261 records through the database search, of which 3125 were duplicates and 4136 were screened (Figure 1). Of these, 712 full-text publications were assessed for eligibility and 191 were included. We further identified 40 records through grey literature sources and citation searching, of which 5 full-text publications and two databases were assessed for eligibility and included. The two included databases were from the National Institute for Communicable Diseases (NICD) periodic aetiologic prevalence surveys among symptomatic primary healthcare clinic attendees in South Africa between 2006 and 2022, and the Centre for Sexual Health and HIV/AIDS Research (CeSHHAR) female sex worker population size estimation study in Zimbabwe in 2017.

**Figure 1:**
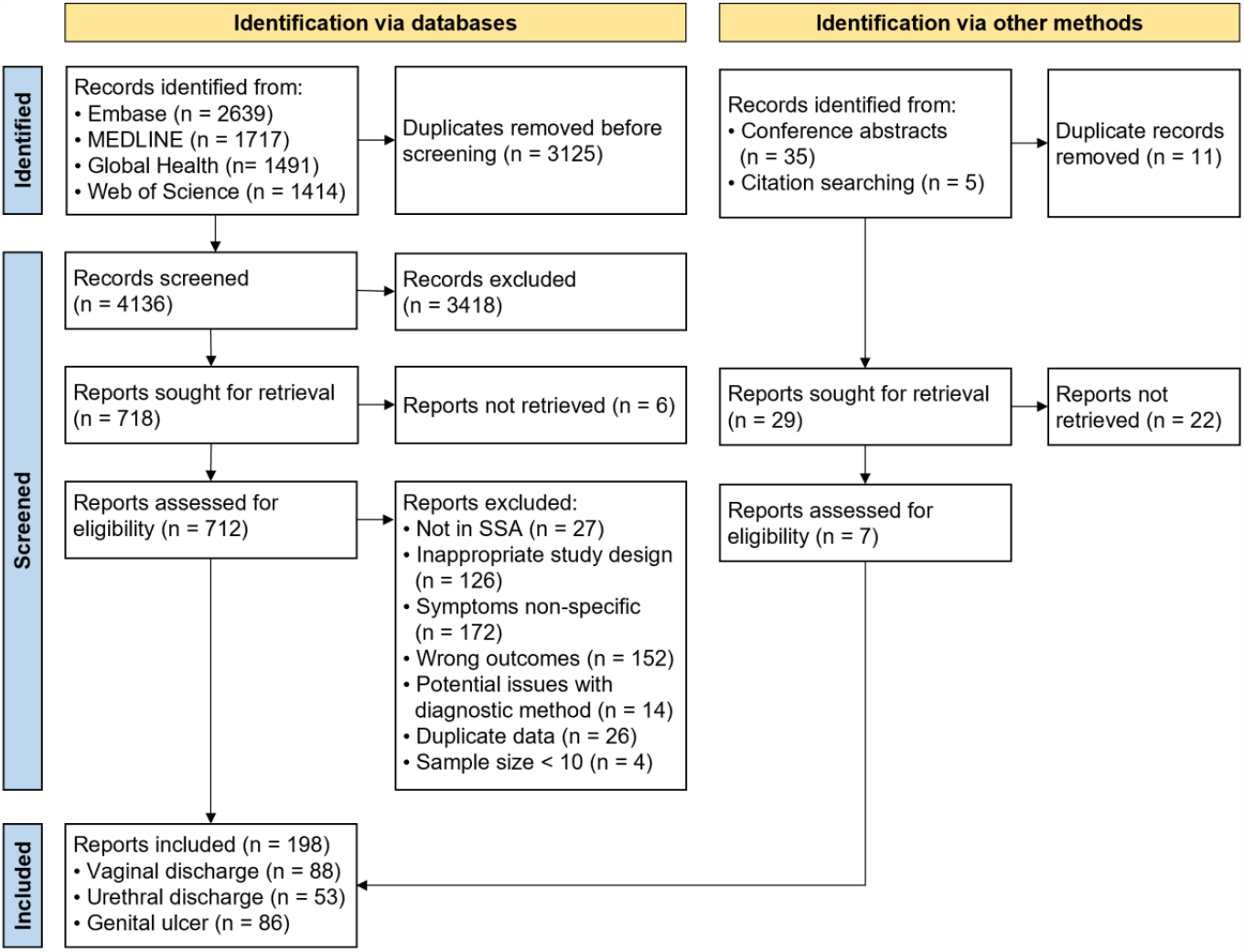
Study selection flowchart. Records include titles and abstracts identified for initial screening. Reports include full text published articles or databases assessed for inclusion.

Overall, 198 reports were included from 183 independent studies (number of studies per symptom (N): N_VD_=83, N_UD_=53, N_GU_=78) spanning 1969 to 2022 (Figure 1, Table 1). Of these, 160 studies focused on a single symptom, 15 studies on two symptoms, and 8 studies on all three symptoms. Less than half of studies aimed to assess the aetiology of genital symptoms (N_VD_=36/83, N_UD_=32/53, N_GU_=38/78). Studies were conducted in 32 of 48 SSA countries included in our search, with a median of 5 studies per country over the full period (Figure S1). Most studies were in eastern Africa (N_VD_=32/83, N_UD_=30/53, N_GU_=49/78) and few were in central Africa (N_VD_=4/83, N_UD_=2/53, N_GU_=2/78). Studies were predominantly in South Africa (N_VD_=12/83, N_UD_=15/53, N_GU_=17/78), and Nigeria (N_VD_=15/83) for vaginal discharge and Kenya (N_GU_=18/78) for genital ulcer. Most studies occurred after 2000 (N_VD_=57/83, N_UD_=26/53) for vaginal discharge and urethral discharge and between 1990 and 1999 (N_GU_=36/78) for genital ulcer. Studies were predominantly among symptomatic clinic attendees (N_VD_=58/83, N_UD_=47/53, N_GU_=61/78) and among individuals of mixed HIV status (N_VD_=31/83, N_UD_=18/53, N_GU_=49/78). Few studies reported the prevalence of HIV (N_VD_=20/83, N_UD_=10/53, N_GU_=42/78) or the mean or median age (N_VD_=25/83, N_UD_=15/53, N_GU_=20/78) among symptomatic participants. Studies were mostly cross-sectional (N_VD_=70/83, N_UD_=45/53, N_GU_=61/78) and with sample sizes of 100 of more (N_VD_=56/83, N_UD_=33/53) for vaginal discharge and urethral discharge, and less than 100 (N_GU_=43/78) for genital ulcer. Most studies tested for more than one pathogen (N_VD_= 56/83, N_UD_=34/53, N_GU_=55/78). The most frequently assessed aetiologies were TV (N_VD_=59/83) and NG (N_VD_=40/83) for vaginal discharge; NG (N_UD_=48/53), CT (N_UD_=30/53) and TV (N_UD_=27/53) for urethral discharge; and TP (N_GU_=66/78) and HD (N_GU_=56/78) for genital ulcer. Study characteristics varied by sub-analysis (Table S10, S11, S12).

**Table 1:** Summary of participant and study characteristics for included studies.

|  |  | Vaginal discharge<br>(N<sub>VD</sub> = 83) | Urethral discharge<br>(N<sub>UD</sub> = 53) | Genital ulcer<br>(N<sub>GU</sub> = 78) | All symptoms<br>(N = 183) |
| --- | --- | --- | --- | --- | --- |
| **Population characteristics** |  |  |  |  |  |
| **Population group<sup>1</sup>** | Symptomatic clinic attendee | 58 (69.9%) | 47 (88.7%) | 61 (78.2%) | 140 (76.5%) |
|  | General | 23 (27.7%) | 2 (3.8%) | 6 (7.7%) | 30 (16.4%) |
|  | Higher-risk general | 2 (2.4%) | 3 (5.7%) | 7 (9%) | 10 (5.5%) |
|  | Key populations | 6 (7.2%) | 1 (1.9%) | 8 (10.3%) | 14 (7.7%) |
|  | Youth | 16 (19.3%) | 6 (11.3%) | 6 (7.7%) | 21 (11.5%) |
| **Sex** | Male | - | 53 (100%) | 20 (25.6%) | 55 (30.1%) |
|  | Female | 83 (100.0%) | - | 21 (26.9%) | 83 (45.4%) |
|  | Mixed | - | - | 37 (47.4%) | 45 (24.6%) |
| **Mean or median age** | < 25 years | 8 (9.6%) | 2 (3.8%) | 1 (1.3%) | 8 (4.4%) |
|  | ≥ 25 years | 17 (20.5%) | 13 (24.5%) | 19 (24.4%) | 45 (24.6%) |
|  | NR | 58 (69.9%) | 38 (71.7%) | 58 (74.4%) | 130 (71%) |
| **HIV status** | Positive | 1 (1.2%) | 0 (0.0%) | 3 (3.8%) | 3 (1.6%) |
|  | Negative | 2 (2.4%) | 1 (1.9%) | 1 (1.3%) | 4 (2.2%) |
|  | Mixed | 31 (37.3%) | 18 (34.0%) | 49 (62.8%) | 78 (42.6%) |
|  | NR | 49 (59%) | 34 (64.2%) | 25 (32.1%) | 98 (53.6%) |
| **HIV prevalence** | < 25% | 10 (12.0%) | 4 (7.5%) | 5 (6.4%) | 17 (9.3%) |
|  | ≥ 25% | 10 (12.0%) | 6 (11.3%) | 37 (47.4%) | 43 (23.5%) |
|  | NR | 63 (75.9%) | 43 (81.1%) | 36 (46.2%) | 123 (67.2%) |
| **Study characteristics** |  |  |  |  |  |
| **Study objective** | Assess symptom aetiology | 36 (43.4%) | 32 (60.4%) | 38 (48.7%) | 87 (47.5%) |
|  | Other | 47 (56.6%) | 21 (39.6%) | 40 (51.3%) | 96 (52.5%) |
| **Region<sup>1</sup>** | Central Africa | 4 (4.8%) | 2 (3.8%) | 2 (2.6%) | 7 (3.8%) |
|  | Eastern Africa | 32 (38.6%) | 30 (56.6%) | 49 (62.8%) | 88 (48.1%) |
|  | Southern Africa | 13 (15.7%) | 11 (20.8%) | 22 (28.2%) | 43 (23.5%) |
|  | Western Africa | 33 (39.8%) | 9 (17%) | 8 (10.3%) | 47 (25.7%) |
|  | Multiple (not stratified) | 1 (1.2%) | 1 (1.9%) | 1 (1.3%) | 2 (1.1%) |
| **Study midpoint year<sup>1,2</sup>** | <1990 | 8 (9.6%) | 10 (18.9%) | 18 (23.1%) | 33 (18%) |
|  | 1990 – 1999 | 21 (25.3%) | 18 (34%) | 36 (46.2%) | 63 (34.4%) |
|  | 2000 – 2009 | 28 (33.7%) | 15 (28.3%) | 19 (24.4%) | 52 (28.4%) |
|  | 2010 – 2022 | 29 (34.9%) | 11 (20.8%) | 9 (11.5%) | 41 (22.4%) |
| **Study design** | Cross sectional | 70 (84.3%) | 45 (84.9%) | 61 (78.2%) | 152 (83.1%) |
|  | Cohort baseline | 10 (12%) | 7 (13.2%) | 12 (15.4%) | 23 (12.6%) |
|  | RCT baseline | 3 (3.6%) | 1 (1.9%) | 5 (6.4%) | 8 (4.4%) |
| **Sampling method** | Non-probability based | 68 (81.9%) | 49 (92.5%) | 71 (91%) | 162 (88.5%) |
|  | Probability based | 9 (10.8%) | 3 (5.7%) | 6 (7.7%) | 13 (7.1%) |
|  | NR | 6 (7.2%) | 1 (1.9%) | 1 (1.3%) | 8 (4.4%) |
| **Sample size<sup>3</sup>** | < 100 | 26 (31.3%) | 20 (37.7%) | 43 (55.1%) | 71 (38.8%) |
|  | ≥ 100 | 57 (68.7%) | 33 (62.3%) | 35 (44.9%) | 112 (61.2%) |
| **Pathogen detected<sup>1</sup>** | BV | 27 (32.5%) | - | - | 27 (14.8%) |
|  | CA | 9 (10.8%) | - | - | 9 (4.9%) |
|  | CS | 33 (39.8%) | - | - | 33 (18.0%) |
|  | CT | 32 (38.6%) | 30 (56.6%) | - | 52 (28.4%) |
|  | HD | - | - | 56 (71.8%) | 56 (30.6%) |
|  | HSV | - | - | 30 (38.5%) | 30 (16.4%) |
|  | HSV-1 | - | - | 14 (17.9%) | 14 (7.7%) |
|  | HSV-2 | - | - | 22 (28.2%) | 22 (12%) |
|  | LGV | - | - | 10 (12.8%) | 10 (5.5%) |
|  | MG | 6 (7.2%) | 8 (15.1%) | - | 12 (6.6%) |
|  | NG | 40 (48.2%) | 48 (90.6%) | - | 75 (41%) |
|  | TP | - | - | 66 (84.6%) | 66 (36.1%) |
|  | TV | 59 (71.1%) | 27 (50.9%) | - | 75 (41.0%) |
|  | None<sup>4</sup> | 14 (16.9%) | 11 (20.8%) | 37 (47.4%) | 55 (30.1%) |
| **Number pathogens extracted per study** | 1 | 27 (32.5%) | 19 (35.8%) | 23 (29.5%) | 54 (29.5%) |
|  | 2 - 3 | 29 (34.9%) | 26 (49.1%) | 39 (50.0%) | 83 (45.4%) |
|  | 4 - 6 | 27 (32.5%) | 8 (15.1%) | 16 (20.5%) | 46 (25.1%) |
| **Risk of bias** | Lower | 17 (20.5%) | 12 (22.6%) | 33 (42.3%) | 48 (26.2%) |
|  | Moderate | 47 (56.6%) | 30 (56.6%) | 33 (42.3%) | 98 (53.6%) |
|  | Higher | 19 (22.9%) | 11 (20.8%) | 12 (15.4%) | 37 (20.3%) |

<sup>1</sup>The same study is included in more than one subcategory when it reports across different variable levels or multiple variable levels are relevant. <sup>2</sup>Mid-point year between start and end dates of data collection. If dates were not reported, year of publication was used as a proxy. <sup>3</sup>Sample size defined as the number of symptomatic individuals tested in the study. If a study used different samples across tests, the largest sample size is reported. <sup>4</sup>Unknown aetiology reported for studies with three or more pathogens extracted. BV: Bacterial vaginosis, CA: *Candida albicans*, CS: *Candida* species (any), CT: *Chlamydia trachomatis*, HD: *Haemophilus ducreyi*, HSV: Herpes simplex virus (unspecified), HSV-1: Herpes simplex virus type 1, HSV-2: Herpes simplex virus type 2, LGV: lymphogranuloma venereum, MG: *Mycoplasma genitalium*, NG: *Neisseria gonorrhoeae*, TP: *Treponema pallidum*, TV: *Trichomonas vaginalis*. None: unknown aetiology. NR: Not reported.

### Regional trends in the aetiology of RTI symptoms

In 2015, CS and BV, rather than STIs, were the primary aetiologies for vaginal discharge (Figure 2A, Table S13A, Table S16A). The proportion of VD cases with CS was 69.4% (95% CI: 44.1-86.6%; number of observations per symptom (n): n_VD_=50), with BV was 50.0% (32.3-67.8%, n_VD_=39), and with CA was 31.5% (12.4-59.9%, n_VD_=9). CT (16.5% [8.7-29.0%], n_VD_=49) and TV (12.9% [7.7-20.7%], n_VD_=78) were the most prominent STIs, while a relatively low proportion of cases were due to NG (6.6% [3.2-13.0%], n_VD_=62) and MG (5.4% [1.8-14.8%], n_VD_=20). A high proportion (25.1% [10.6-48.5%], n_VD_=28) of cases did not have an identified aetiology, despite apparent high levels of co-infection. Diagnosed proportions increased over time for CS (aOR per year: 1.10 [95%CI: 1.05-1.16]), decreased for TV (aOR: 0.94 [0.91-0.97]), and were relatively constant for other RTIs. Temporal trends were difficult to assess for MG, as most studies were conducted after 2005.

**Figure 2:**
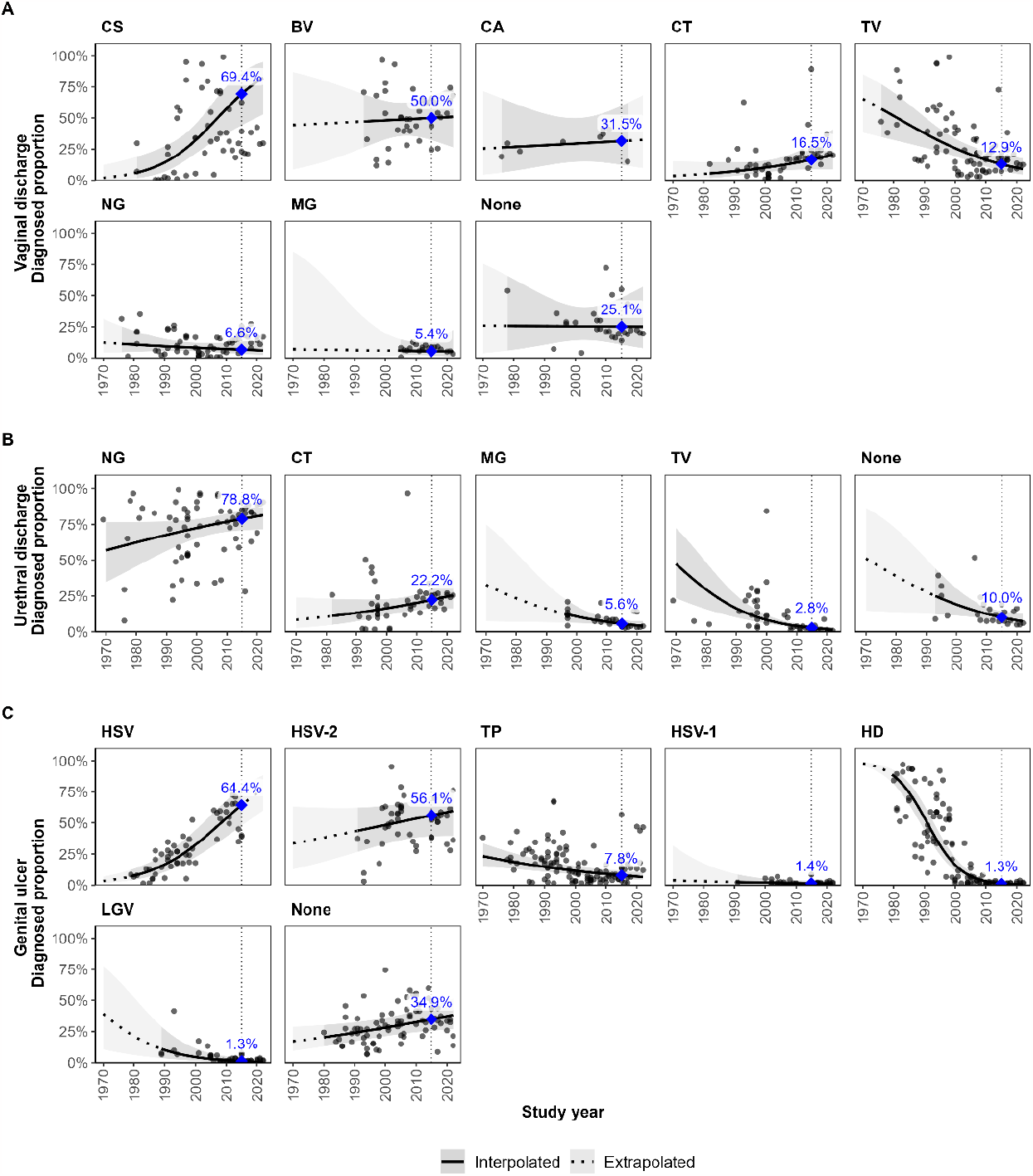
Estimated diagnosed proportion per pathogen over time among symptomatic adults of mixed or unmeasured HIV status in sub-Saharan Africa. Diagnosed proportion per pathogen among adults symptomatic with (A) vaginal discharge, (B) urethral discharge, and (C) genital ulcer. Proportions estimated using generalised linear mixed-effects models for each symptom. Lines and shaded areas represent sex-matched population-weighted mean proportions and 95% confidence intervals. Solid lines and darker shading denote estimates and confidence intervals within the observed data range (interpolated), while dotted lines and lighter shading indicate estimates and confidence intervals beyond that time frame (extrapolated). Blue points and labels represent population-weighted mean proportions in 2015. Vertical dotted lines are through the year 2015. Grey points represent study observations adjusted for diagnostic test performance. BV: Bacterial vaginosis, CA: *Candida albicans*, CS: *Candida* species (any), CT: *Chlamydia trachomatis*, HD: *Haemophilus ducreyi*, HSV: Herpes simplex virus (unspecified), HSV-1: Herpes simplex virus type 1, HSV-2: Herpes simplex virus type 2, LGV: lymphogranuloma venereum, MG: *Mycoplasma genitalium*, NG: *Neisseria gonorrhoeae*, TP: *Treponema pallidum*, TV: *Trichomonas vaginalis*, None: unknown aetiology.

For urethral discharge, NG has persisted over time as the primary aetiology (Figure 2B, Table S13B, Table S16B). In 2015, NG was diagnosed in 78.8% (70.9-85.1%, n_UD_=67) of urethral discharge cases. CT was the predominant (22.2% [16.0-30.1%], n_UD_=48) non-gonococcal cause of urethral discharge, followed by MG (5.6% [3.7-8.2%], n_UD_=29) and TV (2.8% [1.8-4.2%], n_UD_=46). No aetiology was detected in 10.0% (6.9-14.2%, n_UD_=26) of cases. The odds of MG (aOR: 0.95 [0.91-1.00]) and TV (aOR: 0.93 [0.90-0.95]) decreased over time, with no significant change for NG (aOR: 1.02 [1.00-1.04]) or CT (aOR: 1.02 [0.99-1.06]). The proportion per year with unknown aetiology also decreased (aOR: 0.95 [0.91-0.99]).

In 2015, genital ulcer was predominantly caused by HSV-2 (Figure 2C, Table, S13C, Table S16C). The diagnosed proportion of cases for HSV was 64.4% ([47.8-78.1%], n_GU_=54), HSV-2 was 56.1% (39.2-71.6%, n_UD_=46), and HSV-1 was 1.4% (0.5-3.7%, n_UD_=30). TP was the second most prevalent aetiology (7.8% [5.3-11.4%], n_GU_=115), whereas HD (1.3% [0.7-2.1], n_GU_=105) and LGV (1.3% [0.4-3.5], n_GU_=42) were lowest. No aetiology was identified in 34.9% (25.6-45.5%, n_GU_=81) of cases. The distribution of genital ulcer pathogens changed substantially over time. Since most observations for HSV-1 (97%) and HSV-2 (84%) occurred after the year 2000, trends are more reliably assessed for unspecified HSV. In 1980, estimates were highest for HD (87.8% [81.2-92.3%]) and lowest for HSV (7.5% [4.1-13.1%]). The odds of diagnosis increased per year for HSV (aOR: 1.09 [1.07-1.12]) and decreased for HD (aOR: 0.83 [0.82-0.85]), LGV (aOR: 0.92 [0.88-0.95]), and TP (aOR: 0.97 [0.96-0.99]). The odds of having no identified aetiology (aOR: 1.02 [1.00-1.04]) increased over time. Men with genital ulcer had higher odds (aOR: 1.89 [1.24-2.89]) of diagnosis with HD than women, but sex was not a significant predictor for other diagnoses.

Across all three symptoms, there was negligible evidence of regional variation in the estimated diagnosed proportion per pathogen over time (Figure S2, Table S13).

### Population factors influencing the aetiology of each symptom

For each symptom, HIV-status and age were significantly associated with the diagnosis of several RTIs, although the infections with the largest proportions per symptom were generally the same for all sub-groups. For vaginal discharge, the odds were higher among HIV-positive than HIV-negative women for diagnosis with MG (aOR: 2.3 [1.6-3.3], n_VD_=35), TV (aOR: 2.2 [1.6-2.9], n_VD_=44), BV (aOR: 1.8 [1.3-2.4], n_VD_=41) and NG (aOR: 1.6 [1.2-2.2], n_VD_=40), and lower for CS (aOR: 0.6 [0.4-0.8], n_VD_=33; Figure 3A, Table S14A, Table S16A). No HIV-stratified observations were available for CA. The odds were higher among those <25 years than ≥25 years for diagnosis with CT (aOR: 2.4 [1.7-3.3], n_VD_=38), MG (aOR: 2.0 [1.3-3.0], n_VD_=31), CS (aOR: 1.8 [1.3-2.7], n_VD_=37), and NG (aOR: 1.7 [1.2-2.3], n_VD_=43), and lower for those without an identified aetiology (aOR: 0.7 [0.5-0.9], n_VD_=36) (Figure 4A, Table S15A, Table S16A).

For urethral discharge, the odds of TV diagnosis were higher (aOR: 1.8 [1.1-3.1], n_UD_=33) among HIV-positive men than HIV-negative men and lower for CT (aOR: 0.6 [0.4-0.8], n_UD_=36) (Figure 3B, Table S14B, Table S16B). CT was the only RTI associated with age for urethral discharge, with higher odds among <25 years (aOR: 1.6 [1.2-2.2], n_UD_=32) (Figure 4B, Table S15B, Table S16B).

For genital ulcer, the odds of unspecified HSV (aOR: 1.7 [1.1-2.6], n_GU_=51) and HSV-2 (aOR: 1.7 [1.1-2.5], n_GU_=55) were higher among those HIV-positive than HIV-negative, but lower for those with unidentified aetiology (aOR: 0.6 [0.5-0.9], n_GU_=88) (Figure 3C, Table S14C, Table S16C). HD (aOR: 2.4 [1.3-4.4], n_GU_=52) and LGV (aOR: 2.4 [1.3-4.5], n_GU_=43) had higher odds of diagnosis among those <25 years (Figure 4C, Table S15C, Table S16C). In both analyses, men with genital ulcer had lower odds (aOR: 0.5 [0.3-0.7]; aOR: 0.6 [0.4-1.0]) of diagnosis with HSV-2 than women.

### Risk of bias assessment and sensitivity analysis

Most studies (N_VD_=47, N_UD_=30, N_GU_=33) had moderate risk of bias (Table 1, Table S17). Studies with higher risk of bias (N_VD_=19, N_UD_=11, N_GU_=12) were predominantly those with alternate study objectives, insufficient description of study participants and/or settings, only one pathogen assessed, and ambiguous reporting of outcomes. Estimates for the proportion diagnosed per pathogen over time were generally consistent when alternatively including studies of any risk level, or only studies with lower and/or moderate risk of bias (Figure S3).

Estimates derived for RTIs with large diagnostic test performance adjustments (CS, BV, TV, and NG) were higher than those based on reported observations, although trends were relatively similar (Figure S4). Estimates derived using NAAT only generally aligned with trend estimates based on all test types, irrespective of diagnostic test performance adjustment (Figure S5). Notable exceptions in NAAT-based estimates were for vaginal discharge, where TV was stable and NG increased over time, and for genital ulcer, where TP was constant over time. However, NAAT-based estimates were interpolated from shorter time spans than estimates derived using all diagnostic tests.

## Discussion

This systematic review and meta-regression described the distribution, trends, and determinants of vaginal discharge, urethral discharge, and genital ulcer aetiology in sub-Saharan Africa from 1970 to 2022. Our analysis extends previous reviews^24–28^ that informed WHO syndromic management guidelines by incorporating a wider range of studies reporting underlying syndrome aetiologies, specifically in SSA.

The distribution of aetiologies estimated for each symptom were consistent with WHO syndromic management algorithms.^3^ Aligned with guidelines, vaginal discharge management should prioritise candidiasis, bacterial vaginosis, and trichomoniasis. We also identified a notable proportion of cases attributed to chlamydia, emphasising the importance of including speculum examination to improve the vaginal discharge algorithm’s sensitivity to detect cervical infection.^3^ Urethral discharge management should focus on treating gonorrhoea and chlamydia, and genital ulcer management should prioritise herpes and syphilis. Our estimate that 56% of genital ulcer cases in 2015 were caused by HSV-2, with an increasing diagnosed proportion over time, was consistent with another systematic review’s result of 51% during 1990 and 2015.^29^ We did not find evidence of systematic regional variation of aetiologic distributions within SSA. HIV-status and age group were significantly associated with the diagnosis of several RTIs, but the overall hierarchy of aetiologies for each symptom was largely unchanged. Exceptions included higher diagnosed proportions of M. genitalium than chlamydia among younger women with vaginal discharge and HIV-positive men with urethral discharge, and similar diagnosed proportions of chlamydia and gonorrhoea among HIV-positive women with vaginal discharge. Therefore, there was not strong evidence for needing setting or population specific adaptations to the WHO syndromic management algorithms.

Symptom aetiologies have changed over time, particularly for genital ulcer. Candidiasis diagnoses increased among women with vaginal discharge, while the proportion of trichomoniasis diagnoses decreased for both vaginal discharge and urethral discharge. The leading cause of genital ulcer transitioned from chancroid to HSV-2 during 1990 and 2010. These changes underscore the need for periodic aetiologic assessment of syndromes. However, among 32 of 48 SSA countries with studies identified in our review, the publication rate approximated one study every ten years (median of 5 studies per country during 1969 and 2022). Although this falls short of the WHO’s recommended assessment frequency of 2 years,^6^ our findings suggest that an extended assessment interval of 5 years may suffice and that data from neighbouring regions or countries can be used to inform syndromic management protocols.

Aetiologic proportions among symptomatic populations offer insight into the RTI burden, but the two are not equivalent. Diagnosed proportions reflect the prevalence of all possible aetiologies for a particular symptom, conditional on individuals being symptomatic and seeking care. For instance, the proportion of genital ulcer cases attributed to HSV-2 increased over time despite decreasing HSV-2 prevalence in the region, due to more rapid declines in the prevalence of chancroid and syphilis^29–32^ and increased availability of HSV diagnostics.^13^ Nevertheless, aetiologic proportions decreased for syphilis, chancroid, and trichomoniasis, which were consistent with general population prevalence trends in SSA.^14,30–33^ Diagnosed proportions also do not reflect the relative RTI burden among different population groups. Although aetiologic proportions were distributed similarly by HIV-status, age-group, and sex (genital ulcer) for each symptom, the prevalence of RTIs in SSA is higher among women and HIV-positive populations and varies by age.^34,35^ Using our results to estimate overall STI burdens requires additional data on the prevalence of STI syndromes, care seeking, and aetiologies among the general population.

Our analysis had several limitations. We lacked studies in 16 countries, particularly within central Africa. Most studies were not nationally representative and were conducted among a convenience sample of symptomatic individuals seeking healthcare at specific facilities. However, our analysis did not account for factors influencing treatment access, such as urban or rural location,^36–38^ as this information was difficult to attain. We also did not consider symptom severity, recurrence, or treatment history, which could have influenced the aetiologic distributions among study participants but for which data were not consistently available. Furthermore, the populations in our study experiencing abnormal vaginal discharge or urethral discharge were sub-groups of individuals who could potentially be diagnosed with vaginal discharge syndrome (characterised by abnormal discharge, vulval irritation, or itching) or urethral discharge syndrome (abnormal discharge, dysuria, or itching), and may have been presenting with non-infectious aetiologies. Our focus on discharge symptoms rather than syndromes could therefore have influenced estimated aetiologic distributions. As most studies included did not report HIV prevalence or age among symptomatic participants, we were unable to account for this in the overall estimates. Several studies did not differentiate between HSV type or Candida species, yet we considered these data representative of HSV-2 and CA trends, which may have overestimated their contribution. Our estimated diagnosed proportion for HSV-2 was 87% that of unspecified HSV, which was consistent with previous studies.^29^ However, the diagnosed proportion for CA was 45% relative to CS, which was below the expected range of 70-90%.^39,40^ Adjustments to the performance of gram stain and/or wet mount may have overestimated CS proportions, while CA may have been underestimated due to the limited number of observations. Other RTI proportions may also have been over-or under-estimated due to assigned diagnostic test sensitivity and specificity values, despite our efforts to ensure their accuracy. Our modelling approach and estimates did not account for relationships between similar outcomes, such as the contribution of HSV-1 and HSV-2 to unspecified HSV, or the negative correlation between proportions for the same symptom, which depend on co-infection rates. We also did not consider the number of tested pathogens in a study when evaluating proportions with unknown aetiology.

In conclusion, the aetiology of three common STI-related symptoms has evolved over time in SSA, underscoring a changing STI transmission landscape and the need for intermittent re-assessment to inform syndromic management protocols. The observed aetiologic distributions in SSA were consistent with WHO recommended syndromic management algorithms without strong evidence of variation by country, context, or population strata, strengthening the generalisability of our findings to settings lacking data in SSA. Syndrome aetiology assessments are however limited in their utility for infection surveillance. Comprehensive STI surveillance requires prevalence studies among both symptomatic and asymptomatic populations, particularly due to high rates of asymptomatic infection.^1^

## Data Availability

All data produced in the present work are contained in the supplementary materials.

https://github.com/juliamichalow/sti-symptom-aetiology

## Data sharing

Data extracted from included studies and used for analysis are available as supplementary material. Code reproducing the analysis is available from https://github.com/juliamichalow/sti-symptom-aetiology.

## Contributors

JM, JWI-E, AC, and MCB conceptualised the study. JM, OE, MKW, and MW screened titles and abstracts, and full text reports for inclusion. JM, OE, and MKW assessed the risk of bias and extracted data for included studies. JM and BD assessed diagnostic test validity and collated performance characteristics for diagnostic tests. JM performed the data analysis. TK, TM, SC, and FC contributed data, provided input on local epidemiology, and helped interpret results. JM wrote the initial draft of the manuscript, which was then revised with input from all authors. All authors contributed to and approved the final manuscript.

---

**Figure 1:**
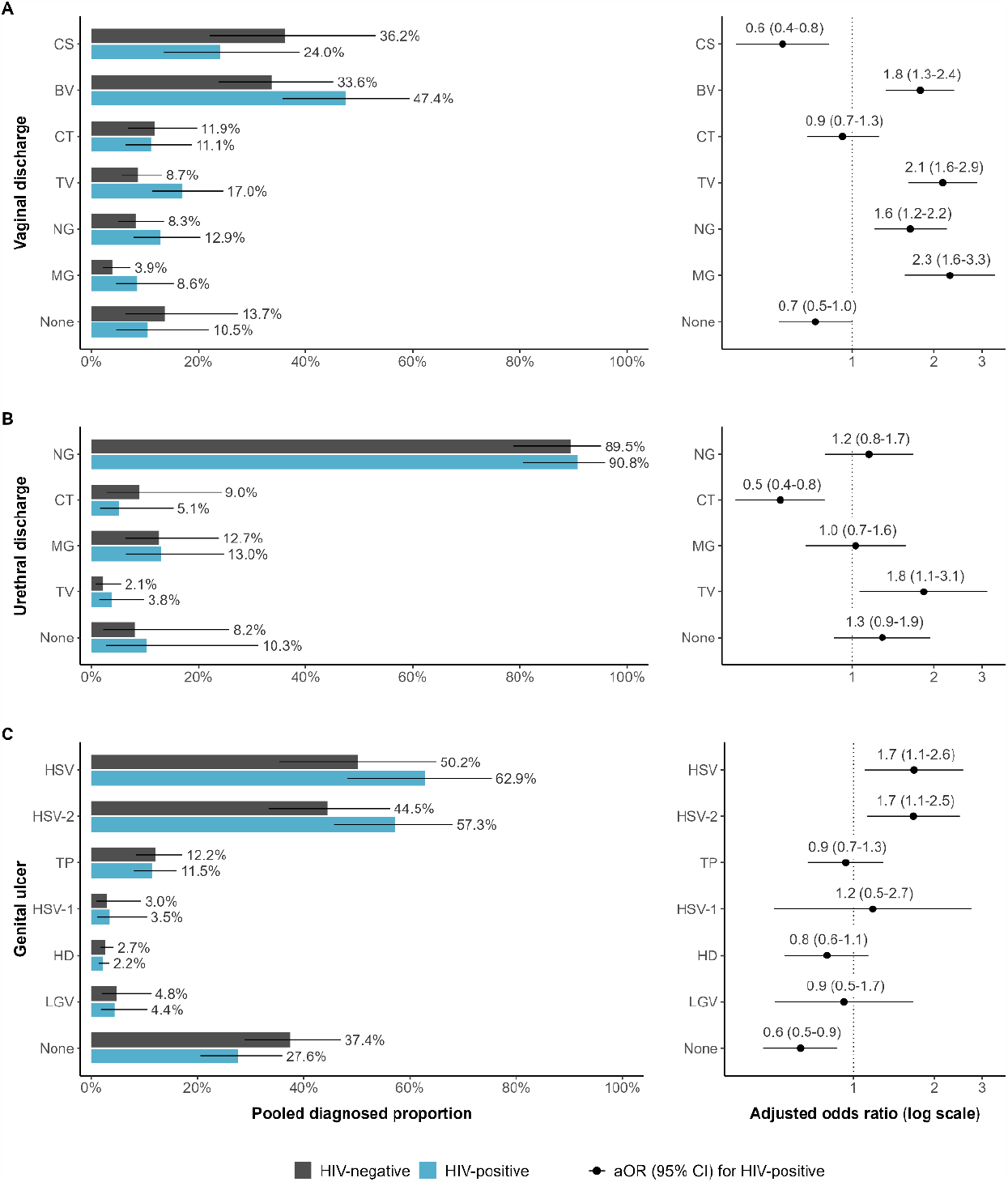
Comparison of diagnosed proportion per pathogen by HIV status in sub-Saharan Africa. Estimated diagnosed proportion per pathogen among HIV-positive and HIV-negative adults in 2015 (left) and adjusted odds of diagnosis per pathogen among HIV-positive compared to HIV-negative adults (right) symptomatic with (A) vaginal discharge, (B) urethral discharge, and (C) genital ulcer. Proportions and odds estimated using generalised linear mixed-effects models for each symptom. Bars represent sex-matched population-weighted mean proportions in 2015. Points represent the adjusted odds of diagnosis among HIV-positive adults. Solid lines represent 95% confidence intervals. BV: Bacterial vaginosis, CS: *Candida* species (any), CT: *Chlamydia trachomatis*, HD: *Haemophilus ducreyi*, HSV: Herpes simplex virus (unspecified), HSV-1: Herpes simplex virus type 1, HSV-2: Herpes simplex virus type 2, LGV: lymphogranuloma venereum, MG: *Mycoplasma genitalium*, NG: *Neisseria gonorrhoeae*, TP: *Treponema pallidum*, TV: *Trichomonas vaginalis*, None: unknown aetiology.

**Figure 2:**
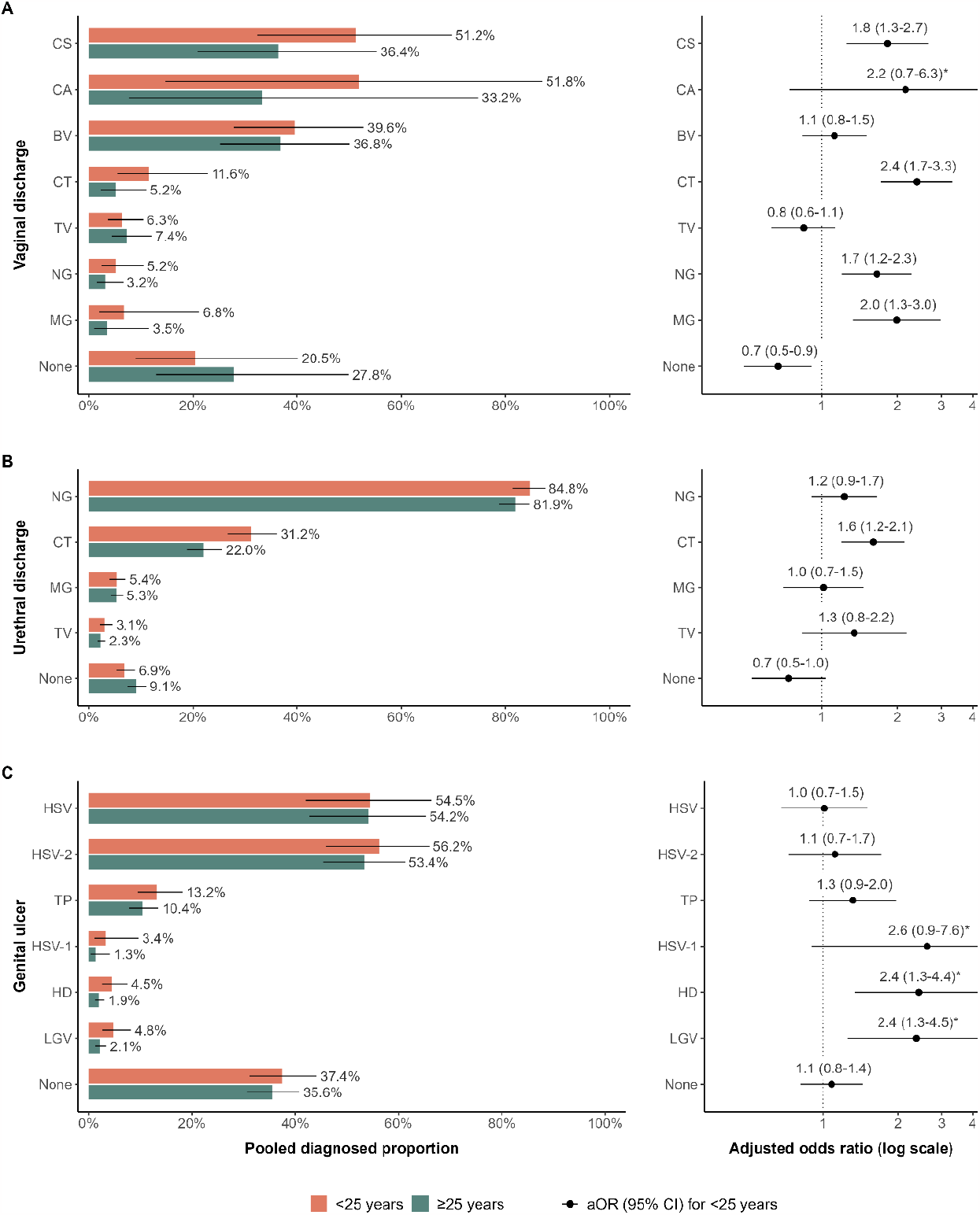
Comparison of diagnosed proportion per pathogen by age group in sub-Saharan Africa. Estimated diagnosed proportion per pathogen among youth < 25 years and adults ≥ 25 years in 2015 (left) and adjusted odds of diagnosis per pathogen among youth compared to adults (right) symptomatic with (A) vaginal discharge, (B) urethral discharge, and (C) genital ulcer. Proportions and odds estimated using generalised linear mixed-effects models for each symptom. Bars represent sex-matched population-weighted mean proportions in 2015. Points represent the adjusted odds of diagnosis among youth. Solid lines represent 95% confidence intervals. BV: Bacterial vaginosis, CA: *Candida albicans*, CS: *Candida* species (any), CT: *Chlamydia trachomatis*, HD: *Haemophilus ducreyi*, HSV: Herpes simplex virus (unspecified), HSV-1: Herpes simplex virus type 1, HSV-2: Herpes simplex virus type 2, LGV: lymphogranuloma venereum, MG: *Mycoplasma genitalium*, NG: *Neisseria gonorrhoeae*, TP: *Treponema pallidum*, TV: *Trichomonas vaginalis*, None: unknown aetiology.

## References

1 World Health Organization. Global health sector strategies on, respectively, HIV, viral hepatitis and sexually transmitted infections for the period 2022-2030. Geneva, 2022.

2 World Health Organization. Global health sector strategy on sexually transmitted infections 2016-2021. 2016 DOI:10.1055/s-2007-970201.

3 World Health Organization. Guidelines for the Management of Symptomatic Sexually Transmitted Infections. Geneva, 2021.

4 Johnson LF, Dorrington RE, Bradshaw D, et al. The effect of syndromic management interventions on the prevalence of sexually transmitted infections in South Africa. Sex Reprod Healthc 2011; 2: 13–20.

5 Wi TE, Ndowa FJ, Ferreyra C, et al. Diagnosing sexually transmitted infections in resource-constrained settings: challenges and ways forward. J. Int. AIDS Soc. 2019; 22: e25343.

6 World Health Organization. Accelerating the global Sexually Transmitted Infections response. Report on the first informal think-tank meeting. Geneva, 2020.

7 World Health Organization. Strategies and laboratory methods for strengthening surveillance of sexually transmitted infections. Geneva, 2012.

8 Taylor MM, Wi T, Gerbase A, et al. Assessment of country implementation of the WHO global health sector strategy on sexually transmitted infections (2016-2021). PLoS One 2022; 17: e0263550.

9 UN Statistics Division. Standard country or area codes for statistical use (M49). https://unstats.un.org/unsd/methodology/m49/ (accessed June 14, 2023).

10 World Health Organization. Laboratory tests for the detection of reproductive tract infections. 1999.

11 Peeling R, Sparling PF. Sexually transmitted diseases: Methods and Protocols, 1st ed. Totowa, NJ: Humana Press, 1999 DOI:10.1385/0896035352.

12 Holmes K, Sparling P, StammW, et al. Sexually Transmitted Diseases, 4th edn. New York: McGraw Hill Medical, 2008.

13 Unemo M, Ballard R, Ison C, et al. Laboratory diagnosis of sexually transmitted infections, including human immunodeficiency virus. Geneva, 2013.

14 World Health Organization. Prevalence and incidence of selected sexually transmitted infections: Methods and results used by WHO to generate 2005 estimates. Geneva, 2011.

15 Flor M, Weiß M, Selhorst T, et al. Comparison of Bayesian and frequentist methods for prevalence estimation under misclassification. BMC Public Health 2020; 20: 1135.

16 Speybroeck N, Devleesschauwer B, Joseph L, et al. Misclassification errors in prevalence estimation: Bayesian handling with care. Int J Public Health 2013; 58: 791–5.

17 Lin L, Chu H. Meta-analysis of proportions using generalized linear mixed models. Epidemiology 2020; 31: 713.

18 UN Population Division. Data Portal. https://population.un.org/dataportal/home (accessed Aug 21, 2023).

19 Munn Z, Moola S, Lisy K,et al.Chapter 5: Systematic reviews of prevalence and incidence. In: Aromataris E, Munn Z, eds. JBI Manual for Evidence Synthesis. JBI, 2020.

20 Stan Development Team. RSran: the R interface to Stan. R package version 2.21.2. 2020. https://mc-stan.org/ (accessed May 29, 2023).

21 Generalized Linear Mixed Models using Template Model Builder [R package glmmTMB version 1.1.7]. 2023; published online April 5. https://cran.r-project.org/package=glmmTMB (accessed Aug 14, 2023).

22 Michalow J, Wybrant M, Walters M, et al. Aetiology of genital ulcer, vaginal discharge, and urethral discharge syndromes in sub-Saharan Africa: a systematic review and meta-analysis. PROSPERO. 2022. https://www.crd.york.ac.uk/prospero/display_record.php?ID=CRD42022348045.

23 Page MJ, McKenzie JE, Bossuyt PM, et al. The PRISMA 2020 statement: an updated guideline for reporting systematic reviews. BMJ 2021; 372. DOI:10.1136/BMJ.N71.

24 World Health Organization. Guidelines for the management of symptomatic sexually transmitted infections. Web Annex A. Syndromic management or point of care tests for urethral discharge: systematic review and mathematical modelling. Geneva, 2021.

25 World Health Organization. Guidelines for the management of symptomatic sexually transmitted infections. Web Annex B. Updated systematic review of the performance of the vaginal discharge syndromic management in treating vaginal and cervical infection. Geneva, 2021.

26 World Health Organization. Guidelines for the management of symptomatic sexually transmitted infections. Web Annex C. Systematic review of risk factors for cervical infections in symptomatic or asymptomatic women. Geneva, 2021.

27 World Health Organization. Guidelines for the management of symptomatic sexually transmitted infections. Web Annex E. Systematic review for syndromic management of genital ulcer disease. Geneva, 2021.

28 World Health Organization. Guidelines for the management of symptomatic sexually transmitted infections. Web Annex G. Systematic review on the role of Mycoplasma genitalium in acute and persistent urethral discharge and pelvic inflammatory disease. Geneva, 2021.

29 Harfouche M, Abu-Hijleh FM, James C, et al. Epidemiology of herpes simplex virus type 2 in sub-Saharan Africa: Systematic review, meta-analyses, and meta-regressions. EClinicalMedicine 2021; 35: 100876.

30 Lewis DA. Epidemiology, clinical features, diagnosis and treatment of Haemophilus ducreyi – a disappearing pathogen? Expert Rev Anti Infect Ther 2014; 12: 687–96.

31 Steen R. Eradicating chancroid. Bull World Heal Organ 2001.

32 Smolak A, Rowley J, Nagelkerke N, et al. Trends and predictors of syphilis prevalence in the general population: Global pooled analyses of 1103 prevalence measures including 136 million syphilis tests. Clin Infect Dis 2018; 66: 1184–91.

33 Rowley J, Hoorn S Vander, Korenromp E, et al. Chlamydia, gonorrhoea, trichomoniasis and syphilis: Global prevalence and incidence estimates, 2016. Bull World Health Organ 2019; 97: 548–62.

34 Grabowski MK, Mpagazi J, Kiboneka S, et al. The HIV and sexually transmitted infection syndemic following mass scale-up of combination HIV interventions in two communities in southern Uganda: a population-based cross-sectional study. Lancet Glob Heal 2022; 10: E1825–34.

35 Kharsany ABM, McKinnon LR, Lewis L, et al. Population prevalence of sexually transmitted infections in a high HIV burden district in KwaZulu-Natal, South Africa: Implications for HIV epidemic control. Int J Infect Dis 2020; 98: 130–7.

36 Ebong USU, Makinde OA. Determinants of treatment seeking behaviour for sexually transmitted infections in Nigeria. Afr J Reprod Health 2021; 25: 105–12.

37 Ogale Id YP, Kennedy CE, Nalugoda F, et al. Nearly half of adults with symptoms of sexually transmitted infections (STIs) did not seek clinical care: A population-based study of treatment-seeking behavior among adults in Rakai, Uganda. PLOS Glob Public Heal 2023; 3: e0001626.

38 Seidu AA, Aboagye RG, Okyere J, et al. Towards the prevention of sexually transmitted infections (STIs): Healthcare-seeking behaviour of women with STIs or STI symptoms in sub-Saharan Africa. Sex Transm Infect 2023; 0: 1–7.

39 Mushi MF, Olum R, Bongomin F. Prevalence, antifungal susceptibility and etiology of vulvovaginal candidiasis in sub-Saharan Africa: a systematic review with meta-analysis and meta-regression. Med Mycol 2022; 9: 7.

40 Willems HME, Ahmed SS, Liu J, et al. Vulvovaginal candidiasis: A current understanding and burning questions. J Fungi 2020; 6.

