## Supplementary file 1 for "Aetiology of vaginal discharge, urethral discharge, and genital ulcer in sub-Saharan Africa: systematic review and meta-regression"

### Supplementary material

Julia Michalow[1*], Magdalene K Walters[1†], Olanrewaju Edun[1†], Max Wybrant[1], Bethan Davies[2], Tendesayi Kufa[3], Thabitha Mathega[3], Sungai T Chabata[4], Frances M Cowan[4,5], Anne Cori[1], Marie-Claude Boily[1], Jeffrey W Imai-Eaton[1,6]

[1] MRC Centre for Global Infectious Disease Analysis, School of Public Health, Imperial College London, London, United Kingdom

[2] MRC Centre for Environment and Health, Department of Epidemiology and Biostatistics, School of Public Health, Imperial College London, London, United Kingdom

[3] Centre for HIV & STI, National Institute for Communicable Diseases, Johannesburg, South Africa

[4] Centre for Sexual Health and HIV AIDS Research (CeSHHAR), Harare, Zimbabwe

[5] Department of International Public Health, Liverpool School of Tropical Medicine, Liverpool, United Kingdom

[6] Center for Communicable Disease Dynamics, Department of Epidemiology, Harvard T.H. Chan School of Public Health, Boston, MA, USA

[†] Contributed equally

**Table S1: Search terms for symptom, pathogen, and sub-Saharan Africa, by database and search domain.**

| |
|---|
| **a) MEDLINE search strategy**<br>Search conducted 25 July 2022 with 1717 articles retrieved. |
| **Symptoms domain:**<br>exp ulcer/ or exp vaginal discharge/ or exp urethritis/ or exp uterine cervicitis/ OR<br>(GUD or ((genital* or anogenital or penile or penis or vagin*) adj2 (ulcer* or vesicle* or papule* or chancr* or granulom* or sore* or lesion*)) or ((genital* or penile or penis or urethra* or vagin* or cervic* or cervix) adj2 discharge) or urethritis or cervicitis or vaginitis).ab,kw,ti |
| **AND RTI pathogen domain:**<br>exp Sexually Transmitted Diseases/ or exp Chancroid/ or exp Haemophilus ducreyi/ or exp Chlamydia infections/ or exp Chlamydia trachomatis/ or exp Lymphogranuloma Venereum/ or exp Gonorrhea/ or exp Neisseria gonorrhoeae/ or exp Syphilis/ or exp Treponema pallidum/ or exp Herpes Simplex/ or exp Herpesvirus 1, Human/ or exp Herpesvirus 2, Human/ or exp HIV infections/ or exp Trichomonas Infections/ or exp Trichomonas vaginalis/ or exp Mycoplasma genitalium/ or exp Vaginosis, Bacterial/ or exp Candidiasis, Vulvovaginal/ or exp Candida albicans/ OR<br>(sexually transmitted disease* or sexually transmitted infection* or STI* or STD* or chancroid* or ducreyi or chlamydia* or Lymphogranuloma Venereum or LGV or gonorrhoea* or gonorrhoeae* or gonorrhea* or syphili* or treponema palli* or herpe* or herpes simplex* or HSV* or HSV type* or HIV* or human immunodeficiency virus or trichomonas* or trichomoniasis* or vaginalis* or mycoplasma or genitalium* or vaginosis* or bacterial vaginosis* or candida* or candidiasis* or albicans).ab,kw,ti |
| **AND Sub-Saharan Africa domain:**<br>exp Africa south of the Sahara/ OR<br>(Africa* or Afriq* or Angola*or Benin* or Botswana* or Motswana* or Batswana* or Burkina* or Burundi* or Cabo Verde* or Cap-Vert* or Cape verde* or Camero* or Central African Republic* or Republique centrafricaine or Chad* or Comor* or Congo* or Democratic Republic of Congo or Republique democratique du Congo DRC or RDC or Cote d'Ivoire or Ivory Coast or Ivorian* or Guinea* or Eritrea* or Erythree*or Ethiop* or Gabon* or Gambi* or Ghana* or Guine* or Equatorial Guinea* or Guinee Equatoriale or Equatoguinean* or Guinea-Bissau* or Kenya* or Lesotho* or Basotho* or Liberia* or Madagas* or Malawi* or Mali* or Mauritani* or Mauri* or Mozambi* or Namibi* or Niger* or Nigeria* or Rwanda* or Rouanda* or Ruanda* or Sao Tome* or Senegal* or Seychel* or Sierra Leone* or Somali* or South Africa* or Afrique du Sud or South Sudan or Soudan de sud or Sudan* or Swazi* or eSwatini* or Tanzani* or Togo* or Uganda* or Ouganda* or Zambi* or Zimbabwe*).mp |
| **b) Embase search strategy**<br>Search conducted 25 July 2022 with 2639 articles retrieved. |
| **Symptoms domain:**<br>exp genital ulcer/ or exp vagina discharge/ or exp urethritis/ or exp urethral discharge/ or exp uterine cervicitis/ OR<br>(GUD or ((genital* or anogenital or penile or penis or vagin*) adj2 (ulcer* or vesicle* or papule* or chancr* or granulom* or sore* or lesion*)) or ((genital* or penile or penis or urethra* or vagin* or cervic* or cervix) adj2 discharge) or urethritis or cervicitis or vaginitis).ab,kw,ti |
| **AND RTI pathogen domain:**<br>exp sexually transmitted disease/ or exp ulcus molle/ or exp Haemophilus ducreyi/ or exp Chlamydia/ or exp Chlamydia trachomatis/ or exp lymphogranuloma venereum/ or exp gonorrhea/ or exp Neisseria gonorrhoeae/ or exp syphilis/ or exp Treponema pallidum/ or<br>exp Herpes simplex virus/ or exp Herpes simplex virus 1/ or exp Herpes simplex virus 2/ or exp Human immunodeficiency virus/ or exp Trichomonas/ or Trichomonas vaginalis/ or exp trichomoniasis/ or exp Mycoplasma genitalium/ or exp vaginitis/ or exp vagina candidiasis/ or Candida albicans/ OR<br>(sexually transmitted disease* or sexually transmitted infection* or STI* or STD* or chancroid* or ducreyi or chlamydia* or Lymphogranuloma Venereum or LGV or gonorrhoea* or gonorrhoeae* or gonorrhea* or chancre* or syphili* or treponema palli* or herpe* or herpes simplex* or HSV* or HSV type* or HIV* or human immunodeficiency virus or trichomonas* or trichomoniasis* or vaginalis* or mycoplasma or genitalium* or vaginosis* or bacterial vaginosis* or candida* or candidiasis* or albicans).ab,kw,ti |
| **AND Sub-Saharan Africa domain**<br>exp Africa south of the Sahara/ or exp africa, eastern/ or exp africa, western/ or exp africa, central/ or exp africa, southern/ OR<br>(Africa* or Afriq* or Angola*or Benin* or Botswana* or Motswana* or Batswana* or Burkina* or Burundi* or Cabo Verde* or Cap-Vert* or Cape verde* or Camero* or Central African Republic* or Republique centrafricaine or Chad* or Comor* or Congo* or Democratic Republic of Congo or Republique democratique du Congo DRC or RDC or Cote d'Ivoire or Ivory Coast or Ivorian* or Guinea* or Eritrea* or Erythree*or Ethiop* or Gabon* or Gambi* or Ghana* or Guine* or Equatorial Guinea* or Guinee Equatoriale or Equatoguinean* or Guinea-Bissau* or Kenya* or Lesotho* or Basotho* or Liberia* or Madagas* or Malawi* or Mali* or Mauritani* or Mauri* or Mozambi* or Namibi* or Niger* or Nigeria* or Rwanda* or Rouanda* or Ruanda* or Sao Tome* or Senegal* or Seychel* or Sierra Leone* or Somali* or South Africa* or Afrique du Sud or South Sudan or Soudan de sud or Sudan* or Swazi* or eSwatini* or Tanzani* or Togo* or Uganda* or Ouganda* or Zambi* or Zimbabwe*).mp |
| **c) Global Health search strategy**<br>Search conducted 25 July 2022 with 1491 articles retrieved. |
| **Symptoms domain:**<br>exp genital ulcers/ or urethritis/ or exp cervicitis/ OR<br>(GUD or ((genital* or anogenital or penile or penis or vagin*) adj2 (ulcer* or vesicle* or papule* or chancr* or granulom* or sore* or lesion*)) or ((genital* or penile or penis or urethra* or vagin* or cervic* or cervix) adj2 discharge) or urethritis or cervicitis or vaginitis).ab,ti,mp. |

| |
|---|
| **AND RTI pathogen domain:**<br>exp sexually transmitted disease/ or exp chancroid/ or exp Haemophilus ducreyi/ or exp Chlamydia/ or exp Chlamydia trachomatis/ or exp gonorrhoea/ or exp Neisseria gonorrhoeae/ or exp syphilis/ or exp Treponema pallidum/ or exp herpes simplex virus/ or exp Human herpesvirus 1/ or exp Human herpesvirus 2/ or exp human immunodeficiency virus/ or exp Trichomonas/ or Trichomonas vaginalis/ or exp trichomoniasis/ or exp Mycoplasma genitalium/ or exp bacterial vaginitis/ or exp Candida albicans/ OR<br>(sexually transmitted disease* or sexually transmitted infection* or STI* or STD* or chancroid* or ducreyi or chlamydia* or Lymphogranuloma Venereum or LGV or gonorrhoea* or gonorrhoeae* or gonorrhea* or chancre* or syphili* or treponema palli* or herpe* or herpes simplex* or HSV* or HSV type* or HIV* or human immunodeficiency virus or trichomonas* or trichomoniasis* or vaginalis* or mycoplasma or genitalium* or vaginosis* or bacterial vaginosis* or candida* or candidiasis* or albicans).ab,ti,mp. |
| **AND Sub-Saharan Africa domain**<br>exp Africa south of the Sahara/ or exp Central Africa/ or exp East Africa/ or exp Southern Africa/ or exp West Africa/ OR (Africa* or Afriq* or Angola*or Benin* or Botswana* or Motswana* or Batswana* or Burkina* or Burundi* or Cabo Verde* or Cap-Vert* or Cape verde* or Camero* or Central African Republic* or Republique centrafricaine or Chad* or Comor* or Congo* or Democratic Republic of Congo or Republique democratique du Congo DRC or RDC or Cote d'Ivoire or Ivory Coast or Ivorian* or Guinea* or Eritrea* or Erythree*or Ethiop* or Gabon* or Gambi* or Ghana* or Guine* or Equatorial Guinea* or Guinee Equatoriale or Equatoguinean* or Guinea-Bissau* or Kenya* or Lesotho* or Basotho* or Liberia* or Madagas* or Malawi* or Mali* or Mauritani* or Mauri* or Mozambi* or Namibi* or Niger* or Nigeria* or Rwanda* or Rouanda* or Ruanda* or Sao Tome* or Senegal* or Seychel* or Sierra Leone* or Somali* or South Africa* or Afrique du Sud or South Sudan or Soudan de sud or Sudan* or Swazi* or eSwatini* or Tanzani* or Togo* or Uganda* or Ouganda* or Zambi* or Zimbabwe*).mp |
| **e) Web of Science search strategy**<br>Search conducted 25 July 2022 with 1414 articles retrieved. |
| **Symptoms domain:**<br>TS=(((genital* or anogenital or penile or penis or vagin*) near/2 (ulcer* or vesicle* or papule* or chancr* or granulom* or sore* or lesion*)) or ((genital* or penile or penis or urethra* or vagin* or cervic* or cervix) near/2 discharge) or GUD or urethritis or cervicitis or vaginitis) |
| **AND RTI pathogen domain:**<br>TS=("sexually transmitted disease*" or "sexually transmitted infection*" or STI* or STD* or chancroid* or ducreyi or chlamydia* or "Lymphogranuloma Venereum" or LGV or gonorrhoea* or gonorrhoeae* or gonorrhea* or chancre* or syphili* or "treponema palli*" or herpe* or "herpes simplex*" or HSV* or HSV type* or HIV* or "human immunodeficiency virus" or trichomonas* or trichomoniasis* or vaginalis* or mycoplasma or genitalium* or vaginosis* or bacterial vaginosis* or candida* or candidiasis* or albicans) |
| **AND Sub-Saharan Africa domain**<br>TS=(Africa* or Afriq* or Angola*or Benin* or Botswana* or Motswana* or Batswana* or Burkina* or Burundi* or Cabo Verde* or Cap-Vert* or Cape verde* or Camero* or Central African Republic* or Republique centrafricaine or Chad* or Comor* or Congo* or Democratic Republic of Congo or Republique democratique du Congo DRC or RDC or Cote d'Ivoire or Ivory Coast or Ivorian* or Guinea* or Eritrea* or Erythree*or Ethiop* or Gabon* or Gambi* or Ghana* or Guine* or Equatorial Guinea* or Guinee Equatoriale or Equatoguinean* or Guinea-Bissau* or Kenya* or Lesotho* or Basotho* or Liberia* or Madagas* or Malawi* or Mali* or Mauritani* or Mauri* or Mozambi* or Namibi* or Niger* or Nigeria* or Rwanda* or Rouanda* or Ruanda* or Sao Tome* or Senegal* or Seychel* or Sierra Leone* or Somali* or South Africa* or Afrique du Sud or South Sudan or Soudan de sud or Sudan* or Swazi* or eSwatini* or Tanzani* or Togo* or Uganda* or Ouganda* or Zambi* or Zimbabwe*) |

#### Table S2: Sub-Saharan African countries included in the analysis, classified by region.

| Region | Countries |
|---|---|
| Eastern Africa | Burundi, Comoros, Djibouti, Eritrea, Ethiopia, Kenya, Madagascar, Malawi, Mauritius, Mozambique, Rwanda, Seychelles, Somalia, South Sudan, Uganda, United Republic of Tanzania, Zambia, Zimbabwe |
| Southern Africa | Botswana, Eswatini, Lesotho, Namibia, South Africa |
| Western Africa | Benin, Burkina Faso, Cabo Verde, Cote d'Ivoire, Gambia, Ghana, Guinea-Bissau, Guinea, Liberia, Mali, Mauritania, Niger, Nigeria, Senegal, Sierra Leone, Togo |
| Central Africa | Angola, Cameroon, Central African Republic, Chad, Congo, Democratic Republic of Congo, Equatorial Guinea, Gabon, Sao Tome and Principe |

Countries classified according to UN M49 Standard.[1]

#### Table S3: Variables extracted from included reports.

| Category | Extracted variables |
|---|---|
| Study characteristics | • Authors<br>• Publication title<br>• Publication year<br>• Years and duration of data collection<br>• Countries of study<br>• Subnational regions or cities of study<br>• Study site names<br>• Study site types<br>• Study design<br>• Study sampling method |
| Participant characteristics | • Study population<br>• Study population age (mean, median, range)<br>• Study population HIV status<br>• Study population HIV prevalence |
| Diagnostic methods | • Diagnostic test and specimen for each infection<br>• Number tested for each infection<br>• Number positive for each infection<br>• Proportion diagnosed positive with each infection |

### Text S1: Preparation and extraction of unpublished data

The literature search identified two databases. The first, from the South Africa National Institute for Communicable Diseases (NICD), covered surveillance among primary healthcare clinic attendees in South Africa from 2006 to 2022. These data were reported in several publications,[2,3,12,4–11] yet the database included unpublished surveillance rounds and enabled additional stratification of outcomes. The second was from the Centre for Sexual Health and HIV/AIDS Research (CeSHHAR) female sex worker population size estimation study in Zimbabwe in 2017. Conference abstracts reporting results from the study indicated relevant data, prompting request for special tabulation.

#### *South Africa NICD data extraction*

Data were available from 13 STI aetiologic surveillance rounds, which were cross-sectional aetiologic prevalence surveys among primary healthcare clinic attendees who were examined and diagnosed with vaginal discharge, urethral discharge, and genital ulcer syndromes (Table S4). Variables were standardised and combined into a single database. We extracted diagnosed proportions, stratified by year, sex, HIV-status, and age group. We calculated HIV-prevalence and mean age for each stratum. Prior full-text publications of these surveillance data were excluded from our review.

### Table S4: Surveys by National Institute for Communicable Diseases in South Africa

| Year(s) | Province | Facilities | Number participants |
|---|---|---|---|
| 2006-2007 | Gauteng | Alexandra Health Centre | 529 (4.0%) |
| | Northern Cape | Betty Gaetsewe Clinic, City Clinic, Masakhane Clinic | 416 (3.2%) |
| 2006-2007 | Western Cape | Spencer Road Clinic | 243 (1.9%) |
| 2007-2008 | Free State | Central Park Clinic, Pelonomi Poly Clinic | 360 (2.7%) |
| 2007-2009 | Gauteng | Alexandra Health Centre | 992 (7.5%) |
| 2010 | Gauteng | Alexandra Health Centre | 1 057 (8.0%) |
| 2010-2012 | Gauteng | Alexandra Health Centre | 720 (5.5%) |
| | Limpopo | Rethabile Heath Centre | 402 (3.0%) |
| 2011-2012 | Gauteng | Alexandra Health Centre | 722 (5.5%) |
| 2013-2014 | Gauteng | Alexandra Health Centre | 546 (4.0%) |
| 2013-2014 | KwaZulu-Natal | Central Durban Clinic | 406 (3.1%) |
| | Mpumalanga | Nelspruit Clinic | 243 (1.9%) |
| 2014-2016 | Eastern Cape | Gqebera Clinic | 92 (0.7%) |
| | Gauteng | Alexandra Health Clinic | 735 (5.6%) |
| | KwaZulu-Natal | East Boom Clinic, Phoenix Clinic | 438 (3.3%) |
| | Mpumalanga | Hluvukani Clinic, Kabokweni Clinic | 436 (3.3%) |
| | North West | Jouberton Clinic | 227 (1.7%) |
| 2017 | Eastern Cape | Zwide Clinic | 259 (1.9%) |
| | Free State | Heidedal Clinic | 129 (1.0%) |
| | Gauteng | Alexandra Clinic | 373 (2.8%) |
| | Western Cape | Khayelitsha Site B Clinic, Khayelitsha Site C Clinic | 223 (1.7%) |
| 2018-2019 | Gauteng | Alexandra Clinic | 356 (2.7%) |
| | Limpopo | Rethabile Health Centre | 139 (1.1%) |
| | Northern Cape | Kimberly Central Clinic | 209 (1.6%) |
| 2020-2022 | Gauteng | Alexandra Health Centre | 855 (6.5%) |
| | KwaZulu-Natal | Prince Cyril Zulu Communicable Disease Centre | 1 032 (7.8%) |
| | Western Cape | Spencer Road Clinic | 399 (3.8%) |
| **Total** | | | 13 159 (100%) |

#### *CeSHHAR female sex worker survey data extraction*

The female sex worker population size estimation study in Zimbabwe included 2507 women (1497 Harare, 808 Bulawayo, 202 Shamva) recruited using respondent-driven sampling (RDS).[13] A random sample of one third of participants were offered genital examination and STI testing, of whom 80 were identified as having abnormal vaginal discharge. Data were survey-weighted using RDS weights to determine diagnosed proportions and 95% confidence intervals, with stratification by HIV status and age-group. We calculated weighted HIV-prevalence and mean age for each stratum. We determined numerators and denominators for each proportion that accounted for the sampling design, using the method described by Stannah et al. (2023).[14] For each observation, we estimated a design effect using the ratio of variances of the RDS-adjusted proportion and simple random sample proportion, which was used to derive the effective sample size and number of individuals who tested positive.

### Text S2: Adjusting reported proportions to account for diagnostic test performance

#### *Diagnostic test performance*

We classified diagnostic tests from included studies into broad categories. Antibody tests included enzyme immunoassays or enzyme-linked immunosorbent assays. Culture included any culture test with a valid medium. Nucleic acid amplification tests (NAAT) included polymerase chain reaction, strand displacement assay, transcription mediated assay, ligase chain reaction, and GeneXpert. Treponemal tests for syphilis included Treponema pallidum Particle Agglutination Assay, Treponema pallidum Hemagglutination Assay, Fluorescent Treponemal Antibody Absorption, Microhemagglutination Assay for Treponema pallidum, and point of care rapid tests. Non-treponemal tests for syphilis included the Venereal Disease Research Laboratory test and Rapid Plasma Reagin test. Syphilis tests were categorised regardless of titre due to variations in reporting.

In studies aimed at detecting candidiasis, extractions initially labelled as *Candida albicans* according to study descriptions were reclassified as *Candida* species if the diagnostic tests used (i.e. culture, wet mount, and gram stain) were not suitable for identifying *Candida albicans*. Studies that described detecting *Gardnerella vaginalis* with diagnostic tests (i.e. Amsel Criteria or Nugent Score) appropriate for detecting bacterial vaginosis were reclassified as such.

Point value sensitivity and specificity estimates for each diagnostic test category were compiled from literature, with priority given to performance characteristics published by the WHO (Tables S5 – S7).[15,16] In cases where multiple sources were available with wide variation, we calculated mean sensitivity and specificity values. We made the following assumptions where data were lacking on performance characteristics. For Candida species, we assumed that the accuracy of gram stain was equivalent to wet mount, although the sensitivity of the former test is likely higher. For *Trichomonas vaginalis*, we assumed that the accuracy of culture using a urine sample was equivalent to that of culture with a genital fluid sample,[16,17] and that the accuracy of wet mount using a urine sample was equivalent to that of wet mount with a genital fluid sample.[16,18] In studies that reported using two diagnostic tests to identify a pathogen, we determined a combined sensitivity and specificity estimate: if either test in the study could be positive, we used the lower sensitivity and lower specificity of the two tests, and if both tests had to be positive, we used the lower sensitivity and higher specificity of the two tests.

#### *Bayesian estimation*

The Rogan-Gladen estimator (Eq. 1) is commonly used to estimate the true prevalence ($p^{true}$) of infection based on observed prevalence ($p^{observed}$) measured by an imperfect diagnostic, accounting for the sensitivity and specificity of a given diagnostic test.[19] However, direct application of the equation can result in negative values for $p^{true}$ when $p^{observed}$ is lower than the probability of a false positive result (1 - $specificity$). We therefore used a Bayesian approach to estimate $p^{true}$ in Eq. 1 by accounting for the observed sample size ($N$) and number of positive test results ($n$) (Eq 2.), and applying a uniform prior probability for $p_{true}$ between 0 and 1 (Eq. 3).[20,21]

$$p^{observed} = p^{true} * \ sensitivity + (1 - p^{true}) * \ (1 - specificity) \quad \text{(Eq. 1)}$$

$$n \sim binomial(N, p^{observed}) \quad \text{(Eq. 2)}$$

$$p^{true} \sim uniform(0,1) \quad \text{(Eq. 3)}$$

The model was fit using Markov Chain Monte Carlo, specifically the NUTS (No-U-Turn Sampler) algorithm, with four chains that each ran for 2000 iterations. Convergence was assessed qualitatively using trace plots of model parameters and quantitatively using the Gelman-Rubin statistic (R-hat). The posterior mean estimate ($\widehat{p^{true}}$ ) and standard error ($se^{true}$) were used to calculate adjusted numerators ($n^{adjusted}$) and denominators ($N^{adjusted}$) for each observation (Eq. 4 and 5), which were then used for pooling in generalized linear mixed-effects models.

$$N^{adjusted} = \frac{\widehat{p^{true}}\left(1 - \widehat{p^{true}}\right)}{(se^{true})^2} \quad \text{(Eq. 4)}$$

$$n^{adjusted} = p^{true} * \ N^{adjusted} \quad \text{(Eq. 5)}$$

### Table S5: Performance of diagnostic tests among women with vaginal discharge.

| Pathogen | Test | Specimen | Number studies | Sensitivity (%) | Specificity (%) | Source |
|---|---|---|---|---|---|---|
| BV | Amsel criteria | Genital fluid | 9 | 51.0 | 98.0 | Dadhwal 2010[22] |
| | Nugent score | Genital fluid | 15 | 87.0 | 95.0 | Thomason 1992[23] Schwebke 1996[24] |
| | Amsel criteria and Nugent score | Genital fluid | 2 | 51.0 | 98.0 | Combined test estimate* |
| | Amsel criteria or Nugent score | Genital fluid | 1 | 51.0 | 95.0 | Combined test estimate* |
| CA | Germ tube | Genital fluid | 7 | 98.9 | 98.5 | Hoppe 1999[25] |
| | NAAT | Genital fluid | 2 | 90.9 | 94.1 | Gaydos 2017[26] |
| CS | Culture | Genital fluid | 8 | 67.0 | 66.0 | WHO 1999[15] |
| | Gram stain | Genital fluid | 11 | 54.6 | 89.6 | Assumed equivalent to wet mount, but sensitivity likely higher. |
| | Wet mount | Genital fluid | 6 | 54.6 | 89.6 | Chatwani 2007[27] Schwebke 2018[28] |
| | Gram stain and culture | Genital fluid | 6 | 54.6 | 89.6 | Combined test estimate* |
| | Wet mount and culture | Genital fluid | 2 | 54.6 | 89.6 | Combined test estimate* |
| CT | Antibody test | Genital fluid | 7 | 65.0 | 100.0 | WHO 2011[16] |
| | Culture | Genital fluid | 1 | 63.4 | 100.0 | WHO 2011[16] |
| | DFA | Genital fluid | 5 | 82.0 | 98.5 | WHO 1999[15] |
| | NAAT | Genital fluid | 15 | 88.6 | 99.6 | WHO 2011[16] |
| | NAAT | Urine | 2 | 87.0 | 99.8 | WHO 2011[16] |
| | Rapid antigen test | Genital fluid | 1 | 48.0 | 98.0 | Grillo-Ardila 2020[29] |
| | Antibody test and NAAT | Genital fluid | 1 | 65.0 | 99.6 | Combined test estimate* |
| | Culture and DFA | Genital fluid | 1 | 97.0 | 99.8 | Combined test estimate* |
| MG | NAAT | Genital fluid | 6 | 93.3 | 97.6 | Gaydos 2019[30] |
| NG | Culture | Genital fluid | 25 | 75.7 | 100.0 | WHO 2011[16] |
| | NAAT | Genital fluid | 14 | 93.3 | 99.2 | WHO 2011[16] |
| | NAAT | Urine | 1 | 91.6 | 100.0 | WHO 2011[16] |
| TV | Culture | Genital fluid | 6 | 68.8 | 100.0 | WHO 2011[16] |
| | NAAT | Genital fluid | 16 | 95.0 | 98.0 | WHO 2011[16] |
| | Wet mount | Genital fluid | 33 | 52.0 | 100.0 | WHO 2011[16] |
| | Culture and NAAT | Genital fluid | 1 | 68.8 | 100.0 | Combined test estimate* |
| | Culture or NAAT | Genital fluid | 1 | 68.8 | 98.0 | Combined test estimate* |
| | Culture and wet mount | Genital fluid | 2 | 52.0 | 100.0 | Combined test estimate* |

Number of studies indicates the total count of studies included within the systematic review that used each diagnostic test and specimen combination for the detection of a specific pathogen.

* Combined sensitivity and specificity estimate determined for studies reporting two diagnostic tests to identify a pathogen. If either test could be positive, the lower sensitivity and lower specificity values were used from the two tests. If both tests had to be positive, the lower sensitivity and higher specificity values were used from the two tests.

### Table S6: Performance of diagnostic tests among men with urethral discharge.

| Pathogen | Test | Specimen | Number studies | Sensitivity (%) | Specificity (%) | Source |
|---|---|---|---|---|---|---|
| CT | Antibody test | Genital fluid | 10 | 84.0 | 98.0 | WHO 1999[15] |
| | Antibody test | Urine | 1 | 84.0 | 98.0 | WHO 1999[15] |
| | Culture | Genital fluid | 1 | 63.4 | 100.0 | WHO 2011[16] |
| | DFA | Genital fluid | 1 | 82.0 | 98.5 | WHO 2011[16] |
| | NAAT | Genital fluid | 8 | 87.5 | 99.2 | WHO 2011[16] |
| | NAAT | Urine | 7 | 87.8 | 99.3 | WHO 2011[16] |
| | Culture and DFA | Genital fluid | 2 | 63.4 | 100.0 | Combined test estimate* |
| MG | NAAT | Genital fluid | 4 | 98.1 | 99.9 | Gaydos 2019[30] |
| | NAAT | Urine | 4 | 89.4 | 99.1 | Gaydos 2019[30] |
| NG | Culture | Genital fluid | 29 | 87.6 | 100.0 | WHO 2011[16] |
| | Gram stain | Genital fluid | 3 | 87.6 | 100.0 | WHO 2011[16] |
| | NAAT | Genital fluid | 6 | 96.1 | 99.0 | WHO 2011[16] |
| | NAAT | Urine | 6 | 80.9 | 99.9 | WHO 2011[16] |
| | Gram stain and culture | Genital fluid | 2 | 87.6 | 100.0 | Combined test estimate* |
| | Gram stain or NAAT | Genital fluid and urine | 1 | 87.6 | 99.9 | Combined test estimate* |
| | Culture or NAAT | Genital fluid | 1 | 87.6 | 99.0 | Combined test estimate* |
| TV | Culture | Genital fluid | 4 | 74.5 | 100.0 | WHO 2011[16] |
| | Culture | Urine | 1 | 74.5 | 100.0 | Performance of urine culture and urethral swab culture assumed equivalent, based on WHO 2011[16] and Kaydos-Daniels 2004[17] |
| | NAAT | Genital fluid | 7 | 81.6 | 97.7 | WHO 2011[16] |
| | NAAT | Urine | 4 | 96.0 | 97.7 | WHO 2011[16] |
| | Wet mount | Genital fluid | 5 | 44.0 | 100.0 | WHO 2011[16] |
| | Wet mount | Urine | 1 | 44.0 | 100.0 | Performance of urine wet mount and urethral swab wet mount assumed equivalent due to insufficient performance data, although urine wet mount expected to be substantially less sensitive, based on WHO 2011[16] and Hobbs 2006[18] |
| | Culture or NAAT | Genital fluid and urine | 1 | 44.0 | 97.7 | Combined test estimate* |
| | Culture and wet mount | Genital fluid | 2 | 44.0 | 100.0 | Combined test estimate* |
| | Culture or wet mount | Urine | 1 | 44.0 | 100.0 | Combined test estimate* |

Number of studies indicates the total count of studies included within the systematic review that used each diagnostic test and specimen combination for the detection of a specific pathogen.

* Combined sensitivity and specificity estimate determined for studies reporting two diagnostic tests to identify a pathogen. If either test could be positive, the lower sensitivity and lower specificity values were used from the two tests. If both tests had to be positive, the lower sensitivity and higher specificity values were used from the two tests.

### Table S7: Performance of diagnostic tests among adults with genital ulcer.

| Pathogen | Test | Specimen | Number studies | Sensitivity (%) | Specificity (%) | Source |
|---|---|---|---|---|---|---|
| HD | Culture | Genital ulcer | 30 | 73.0 | 100.0 | WHO 1999[15] |
| | NAAT | Genital ulcer | 26 | 95.1 | 99.9 | Paz-Bailey 2005[31] |
| | Culture or NAAT | Genital ulcer | 1 | 73.0 | 99.9 | Combined test estimate* |
| | Culture and NAAT | Genital ulcer | 1 | 73.0 | 100.0 | Combined test estimate* |
| HSV | Culture | Genital ulcer | 16 | 75.0 | 100.0 | Legoff 2014[32] |
| | DFA | Genital ulcer | 1 | 80.0 | 95.0 | Legoff 2014[32] |
| | NAAT | Genital ulcer | 13 | 98.0 | 100.0 | Legoff 2014[32] |
| HSV-1 | DFA | Genital ulcer | 1 | 80.0 | 95.0 | Legoff 2014[32] |
| | NAAT | Genital ulcer | 13 | 99.0 | 99.0 | Arshad 2019[33] |
| HSV-2 | DFA | Genital ulcer | 2 | 80.0 | 95.0 | Legoff 2014[32] |
| | NAAT | Genital ulcer | 20 | 100.0 | 98.5 | Arshad 2019[33] |
| LGV | DFA | Genital ulcer | 2 | 95.5 | 99.4 | Diagnostic Hybrids 2008[34] |
| | NAAT | Genital ulcer | 8 | 95.5 | 98.5 | Touati 2021[35] |
| TP | Darkfield | Genital ulcer | 10 | 80.0 | 98.5 | WHO 1999[15] |
| | DFA | Genital ulcer | 1 | 86.5 | 94.5 | Peeling 2004[36] |
| | NAAT | Genital ulcer | 28 | 91.0 | 99.0 | WHO 1999[15] |
| | Non-treponemal | Blood | 6 | 91.9 | 97.9 | Mean sensitivity and specificity, from Hambie 1983[37], Perryman 1982[38], and Parham 1984[39] regardless of titre due to varied reporting |
| | Treponemal | Blood | 3 | 93.0 | 95.7 | Mean sensitivity and specificity from Cantor 2016[40], Sena 2010[41], and Park 2019[42] |
| | Non-treponemal and treponemal | Blood | 20 | 100.0 | 100.0 | Estimated performance; non-treponemal test performance regardless of titre due to varied reporting |
| | Darkfield or treponemal | Genital ulcer and blood | 1 | 80.0 | 95.7 | Combined test estimate* |
| | Darkfield, or non-treponemal and treponemal | Genital ulcer and blood | 1 | 80.0 | 98.5 | Combined test estimate* |
| | NAAT, or non-treponemal and treponemal | Genital ulcer and blood | 1 | 91.0 | 99.0 | Combined test estimate* |

Number of studies indicates the total count of studies included within the systematic review that used each diagnostic test and specimen combination for the detection of a specific pathogen.

* Combined sensitivity and specificity estimate determined for studies reporting two diagnostic tests to identify a pathogen. If either test could be positive, the lower sensitivity and lower specificity values were used from the two tests. If both tests had to be positive, the lower sensitivity and higher specificity values were used from the two tests.

**Table S8: Risk of bias assessment tool.**

| Indicator | Definition of yes | Yes | No | Unclear |
|---|---|---|---|---|
| 1. Was the study specifically designed to estimate the outcome/s of interest? | The primary objective of the study was to determine sexually transmitted / reproductive tract infection aetiology among individuals symptomatic with genital ulcer, vaginal discharge and/or urethral discharge. | □ | □ | □ |
| 2. Were the criteria for inclusion in the sample clearly defined? | The inclusion/exclusion criteria clearly define the population being recruited into the study. This includes their sex, age group (age range, or classified as youth or adult), and population type (e.g. general population, key population, symptomatic clinic attendee). | □ | □ | □ |
| 3. Were study participants and their setting described in sufficient detail to determine representativeness of the target population? | There is a clear description of study participant characteristics, including their sex, age group and HIV status. The study site locations are described. The name of the health facility is reported, or details of the facility are described in terms of urban/rural and sector (private, government, non-profit) classification. | □ | □ | □ |
| 4. Were study participants recruited in an appropriate way? | The study used either consecutive or random sampling methods for participant recruitment. If it is claimed that a random sample was chosen, the method of randomization should be stated. It is acceptable if studies do not say 'consecutive' but instead describe consecutive enrolment, i.e., 'all patients from …. till …. were included'. | □ | □ | □ |
| 5. Was the participation rate adequate to minimise selection bias? | The participation rate in the study was at least 80%. Participation is defined as providing specimens for STI testing, among those identified as having symptoms. | □ | □ | □ |
| 6. Were objective, standard criteria used to define participant symptoms? | There is a clear, consistently applied definition to identify participants with the symptom(s) of focus in the study. Specifically, all participants were examined by a healthcare provider to identify their symptoms. The classification of symptoms is not based solely on participant self-reporting. | □ | □ | □ |
| 7. Was the sample size sufficient to measure the outcome? | The sample size is justified through a calculation, or the sample includes at least 100 participants. | □ | □ | □ |
| 8. Were the outcomes measured in a reliable and consistent way? | All data were collected directly from the participants. Proxy data (i.e. medical records) were not used in any instances. The same mode of sample collection and diagnosis was used for all participants. | □ | □ | □ |
| 9. Were multiple pathogens assessed to prevent potential misdiagnoses? | Symptomatic participants were tested for at least 3 different sexually transmitted / reproductive tract infection pathogens. | □ | □ | □ |
| 10. Was the reporting of the outcome(s) unambiguous? | The numerators, denominator(s) and proportion are clearly reported for each infection. The results are internally consistent. The reported proportion was not different when calculated from reported numerators and denominators. | □ | □ | □ |

Tool adapted from the Joanna Briggs Institute appraisal tool for prevalence studies.[43]

### Table S9: PRISMA checklist

| Section and Topic | Item | Checklist item | Location reported |
|---|---|---|---|
| **TITLE** | | | |
| Title | 1 | Identify the report as a systematic review. | Title |
| **ABSTRACT** | | | |
| Abstract | 2 | See the PRISMA 2020 for Abstracts checklist. | Abstract |
| **INTRODUCTION** | | | |
| Rationale | 3 | Describe the rationale for the review in the context of existing knowledge. | Introduction |
| Objectives | 4 | Provide an explicit statement of the objective(s) or question(s) the review addresses. | Introduction |
| **METHODS** | | | |
| Eligibility criteria | 5 | Specify the inclusion and exclusion criteria for the review and how studies were grouped for the syntheses. | Methods |
| Information sources | 6 | Specify all databases, registers, websites, organisations, reference lists and other sources searched or consulted to identify studies. Specify the date when each source was last searched or consulted. | Methods, Figure 1 |
| Search strategy | 7 | Present the full search strategies for all databases, registers and websites, including any filters and limits used. | Methods, Table S1-S2 |
| Selection process | 8 | Specify the methods used to decide whether a study met the inclusion criteria of the review, including how many reviewers screened each record and each report retrieved, whether they worked independently, and if applicable, details of automation tools used in the process. | Methods |
| Data collection process | 9 | Specify the methods used to collect data from reports, including how many reviewers collected data from each report, whether they worked independently, any processes for obtaining or confirming data from study investigators, and if applicable, details of automation tools used in the process. | Methods |
| Data items | 10a | List and define all outcomes for which data were sought. Specify whether all results that were compatible with each outcome domain in each study were sought (e.g. for all measures, time points, analyses), and if not, the methods used to decide which results to collect. | Methods |
| | 10b | List and define all other variables for which data were sought (e.g. participant and intervention characteristics, funding sources). Describe any assumptions made about any missing or unclear information. | Methods, Table S3 |
| Study risk of bias assessment | 11 | Specify the methods used to assess risk of bias in the included studies, including details of the tool(s) used, how many reviewers assessed each study and whether they worked independently, and if applicable, details of automation tools used in the process. | Methods, Table S8 |
| Effect measures | 12 | Specify for each outcome the effect measure(s) (e.g. risk ratio, mean difference) used in the synthesis or presentation of results. | Methods |
| Synthesis methods | 13a | Describe the processes used to decide which studies were eligible for each synthesis (e.g. tabulating the study intervention characteristics and comparing against the planned groups for each synthesis (item #5)). | Methods |
| | 13b | Describe any methods required to prepare the data for presentation or synthesis, such as handling of missing summary statistics, or data conversions. | Methods, Text S1-S2, Table S5-S7 |
| | 13c | Describe any methods used to tabulate or visually display results of individual studies and syntheses. | Methods |
| | 13d | Describe any methods used to synthesize results and provide a rationale for the choice(s). If meta-analysis was performed, describe the model(s), method(s) to identify the presence and extent of statistical heterogeneity, and software package(s) used. | Methods |
| | 13e | Describe any methods used to explore possible causes of heterogeneity among study results (e.g. subgroup analysis, meta-regression). | Methods |
| | 13f | Describe any sensitivity analyses conducted to assess robustness of the synthesized results. | Methods |
| Reporting bias assessment | 14 | Describe any methods used to assess risk of bias due to missing results in a synthesis (arising from reporting biases). | Methods |

| Section and Topic | Item | Checklist item | Location reported |
|---|---|---|---|
| Certainty assessment | 15 | Describe any methods used to assess certainty (or confidence) in the body of evidence for an outcome. | Not applicable |
| | | **RESULTS** | |
| Study selection | 16a | Describe the results of the search and selection process, from the number of records identified in the search to the number of studies included in the review, ideally using a flow diagram. | Results, Figure 1 |
| | 16b | Cite studies that might appear to meet the inclusion criteria, but which were excluded, and explain why they were excluded. | Figure 1 |
| Study characteristics | 17 | Cite each included study and present its characteristics. | Data appendix |
| Risk of bias in studies | 18 | Present assessments of risk of bias for each included study. | Table S17 |
| Results of individual studies | 19 | For all outcomes, present, for each study: (a) summary statistics for each group (where appropriate) and (b) an effect estimate and its precision (e.g. confidence/credible interval), ideally using structured tables or plots. | Data appendix |
| Results of syntheses | 20a | For each synthesis, briefly summarise the characteristics and risk of bias among contributing studies. | Results, Table 1, Table S10-S12 |
| | 20b | Present results of all statistical syntheses conducted. If meta-analysis was done, present for each the summary estimate and its precision (e.g. confidence/credible interval) and measures of statistical heterogeneity. If comparing groups, describe the direction of the effect. | Results, Figure 2-4, Table S16, Figure S2 |
| | 20c | Present results of all investigations of possible causes of heterogeneity among study results. | Table S14-S16 |
| | 20d | Present results of all sensitivity analyses conducted to assess the robustness of the synthesized results. | Figure S3-S4 |
| Reporting biases | 21 | Present assessments of risk of bias due to missing results (arising from reporting biases) for each synthesis assessed. | Not applicable |
| Certainty of evidence | 22 | Present assessments of certainty (or confidence) in the body of evidence for each outcome assessed. | Not applicable |
| | | **DISCUSSION** | |
| Discussion | 23a | Provide a general interpretation of the results in the context of other evidence. | Discussion |
| | 23b | Discuss any limitations of the evidence included in the review. | Discussion |
| | 23c | Discuss any limitations of the review processes used. | Discussion |
| | 23d | Discuss implications of the results for practice, policy, and future research. | Discussion |
| | | **OTHER INFORMATION** | |
| Registration and protocol | 24a | Provide registration information for the review, including register name and registration number, or state that the review was not registered. | Methods |
| | 24b | Indicate where the review protocol can be accessed, or state that a protocol was not prepared. | Methods |
| | 24c | Describe and explain any amendments to information provided at registration or in the protocol. | Not applicable |
| Support | 25 | Describe sources of financial or non-financial support for the review, and the role of the funders or sponsors in the review. | Funding |
| Competing interests | 26 | Declare any competing interests of review authors. | Declaration of interests |
| Availability of data, code, other materials | 27 | Report which of the following are publicly available and where they can be found: template data collection forms; data extracted from included studies; data used for all analyses; analytic code; any other materials used in the review. | Data availability |

Source: Preferred Reporting Items for Systematic reviews and Meta-Analyses (PRISMA) 2020[44]

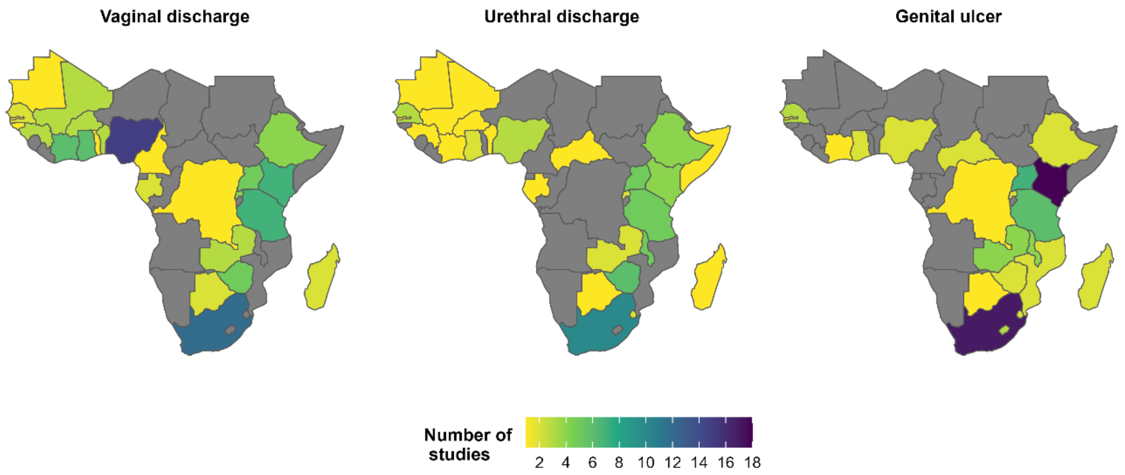

**Figure S1: Number of studies included per symptom by country in sub-Saharan Africa.**

Number of studies among individuals with vaginal discharge, urethral discharge, and genital ulcer that were included in the systematic review. Total studies were 83 for vaginal discharge, 53 for urethral discharge, and 78 for genital ulcer.

**Table S10: Summary of participant and study characteristics for studies included in the analysis of regional trends in aetiology, among adults of mixed or unmeasured HIV status.**

| | | Vaginal discharge (N=76) | Urethral discharge (N=49) | Genital ulcer (N=72) | All symptoms (N=172) |
|---|---|---|---|---|---|
| **Population characteristics** | | | | | |
| **Population group**[1] | Symptomatic clinic attendee | 53 (69.7%) | 44 (89.8%) | 57 (79.2%) | 133 (77.3%) |
| | General | 23 (30.3%) | 2 (4.1%) | 5 (6.9%) | 29 (16.9%) |
| | Higher-risk general | 2 (2.6%) | 3 (6.1%) | 7 (9.7%) | 10 (5.8%) |
| | Key populations | 6 (7.9%) | 1 (2%) | 7 (9.7%) | 13 (7.6%) |
| | Youth | - | - | - | - |
| **Sex** | Male | - | 49 (100.0%) | 19 (26.4%) | 53 (30.8%) |
| | Female | 76 (100.0%) | - | 19 (26.4%) | 78 (45.3%) |
| | Mixed | - | - | 34 (47.2%) | 41 (23.8%) |
| **Mean or median age** | < 25 years | 7 (9.2%) | 2 (4.1%) | 1 (1.4%) | 7 (4.1%) |
| | ≥ 25 years | 17 (22.4%) | 12 (24.5%) | 19 (26.4%) | 43 (25%) |
| | NR | 52 (68.4%) | 35 (71.4%) | 52 (72.2%) | 122 (70.9%) |
| **HIV prevalence** | < 25% | 6 (7.9%) | 1 (2%) | 4 (5.6%) | 11 (6.4%) |
| | ≥ 25% | 8 (10.5%) | 6 (12.2%) | 32 (44.4%) | 38 (22.1%) |
| | NR | 62 (81.6%) | 42 (85.7%) | 36 (50%) | 123 (71.5%) |
| **Study characteristics** | | | | | |
| **Region**[1] | Central Africa | 4 (5.3%) | 2 (4.1%) | 1 (1.4%) | 6 (3.5%) |
| | Eastern Africa | 28 (36.8%) | 26 (53.1%) | 43 (59.7%) | 80 (46.5%) |
| | Southern Africa | 10 (13.2%) | 11 (22.4%) | 21 (29.2%) | 39 (22.7%) |
| | Western Africa | 33 (43.4%) | 9 (18.4%) | 6 (8.3%) | 45 (26.2%) |
| | Multiple (not stratified) | 1 (1.3%) | 1 (2.0%) | 1 (1.4%) | 2 (1.2%) |
| **Study midpoint year**[1,2] | <1990 | 8 (10.5%) | 9 (18.4%) | 18 (25.0%) | 32 (18.6%) |
| | 1990 – 1999 | 19 (25.0%) | 16 (32.7%) | 34 (47.2%) | 60 (34.9%) |
| | 2000 – 2009 | 27 (35.5%) | 14 (28.6%) | 17 (23.6%) | 50 (29.1%) |
| | 2010 – 2022 | 25 (32.9%) | 11 (22.4%) | 7 (9.7%) | 36 (20.9%) |
| **Study design** | Cross sectional | 65 (85.5%) | 41 (83.7%) | 56 (77.8%) | 144 (83.7%) |
| | Cohort baseline | 9 (11.8%) | 7 (14.3%) | 12 (16.7%) | 22 (12.8%) |
| | RCT baseline | 2 (2.6%) | 1 (2.0%) | 4 (5.6%) | 6 (3.5%) |
| **Sampling method** | Non-probability based | 65 (85.5%) | 47 (95.9%) | 68 (94.4%) | 156 (90.7%) |
| | Probability based | 5 (6.6%) | 1 (2.0%) | 3 (4.2%) | 8 (4.7%) |
| | NR | 6 (7.9%) | 1 (2.0%) | 1 (1.4%) | 8 (4.7%) |
| **Sample size**[3] | < 100 | 22 (28.9%) | 16 (32.7%) | 42 (58.3%) | 67 (39.0%) |
| | ≥ 100 | 54 (71.1%) | 33 (67.3%) | 30 (41.7%) | 105 (61.0%) |
| **Number pathogens extracted per study** | 1 | 23 (30.3%) | 17 (34.7%) | 20 (27.8%) | 50 (29.1%) |
| | 2 - 3 | 29 (38.2%) | 24 (49.0%) | 36 (50.0%) | 79 (45.9%) |
| | 4 - 6 | 24 (31.6%) | 8 (16.3%) | 16 (22.2%) | 43 (25.0%) |
| **Risk of bias** | Lower | 16 (21.1%) | 12 (24.5%) | 32 (44.4%) | 47 (27.3%) |
| | Moderate | 44 (57.9%) | 27 (55.1%) | 29 (40.3%) | 91 (52.9%) |
| | Higher | 16 (21.1%) | 10 (20.4%) | 11 (15.3%) | 34 (19.8%) |

[1]The same study is included in more than one subcategory when it reports across different variable levels or multiple variable levels are relevant.

[2] Midpoint year between start and end dates of data collection. If dates were not reported, year of publication was used as a proxy.

[3] Sample size defined as the number of symptomatic individuals tested in the study. If a study used different samples across tests, the largest sample size is reported.

### Table S11: Summary of participant and study characteristics for studies included in the analysis to assess the effect of HIV status, among adults of known HIV status.

| | | Vaginal discharge (N=10) | Urethral discharge (N=5) | Genital ulcer (N=26) | All symptoms (N=37) |
|---|---|---|---|---|---|
| **Population characteristics** | | | | | |
| **Population group[1]** | Symptomatic clinic attendee | 9 (90.0%) | 5 (100%) | 22 (84.6%) | 32 (86.5%) |
| | General | - | - | 1 (3.8%) | 1 (2.7%) |
| | Higher-risk general | - | - | 2 (7.7%) | 2 (5.4%) |
| | Key populations | 1 (10.0%) | - | 3 (11.5%) | 4 (10.8%) |
| | Youth | - | - | - | - |
| **Sex[1]** | Male | - | 5 (100.0%) | 11 (42.3%) | 15 (40.5%) |
| | Female | 10 (100.0%) | - | 9 (34.6%) | 16 (43.2%) |
| | Mixed | - | - | 10 (38.5%) | 10 (27.0%) |
| **HIV status[1]** | Positive | 8 (80.0%) | 5 (100.0%) | 25 (96.2%) | 33 (89.2%) |
| | Negative | 9 (90.0%) | 4 (80.0%) | 23 (88.5%) | 34 (91.9%) |
| **Study characteristics** | | | | | |
| **Region[1]** | Central Africa | - | - | - | - |
| | Eastern Africa | 3 (30.0%) | 2 (40.0%) | 14 (53.8%) | 18 (48.6%) |
| | Southern Africa | 4 (40.0%) | 3 (60.0%) | 9 (34.6%) | 13 (35.1%) |
| | Western Africa | 3 (30.0%) | - | 2 (7.7%) | 5 (13.5%) |
| | Multiple (not stratified) | - | - | 1 (3.8%) | 1 (2.7%) |
| **Study midpoint year[1,2]** | <1990 | - | 1 (20.0%) | 2 (7.7%) | 3 (8.1%) |
| | 1990 – 1999 | - | 1 (20.0%) | 13 (50.0%) | 9 (24.3%) |
| | 2000 – 2009 | 5 (50.0%) | 2 (40.0%) | 8 (30.8%) | 13 (35.1%) |
| | 2010 – 2022 | 6 (60.0%) | 2 (40.0%) | 5 (19.2%) | 14 (37.8%) |
| **Study design** | Cross sectional | 7 (70.0%) | 4 (80.0%) | 18 (69.2%) | 25 (67.6%) |
| | Cohort baseline | 1 (10.0%) | 1 (20.0%) | 5 (19.2%) | 7 (18.9%) |
| | RCT baseline | 2 (20.0%) | - | 3 (11.5%) | 5 (13.5%) |
| **Sampling method** | Non-probability based | 7 (70.0%) | 5 (100.0%) | 23 (88.5%) | 32 (86.5%) |
| | Probability based | 3 (30.0%) | - | 3 (11.5%) | 5 (13.5%) |
| **Sample size[3]** | < 100 | 3 (30.0%) | 1 (20.0%) | 13 (50.0%) | 16 (43.2%) |
| | ≥ 100 | 7 (70.0%) | 4 (80.0%) | 13 (50.0%) | 21 (56.8%) |
| **Number pathogens extracted per study** | 1 | 1 (10.0%) | 0 (0.0%) | 4 (15.4%) | 4 (10.8%) |
| | 2 - 3 | 3 (30.0%) | 3 (60.0%) | 15 (57.7%) | 21 (56.8%) |
| | 4 - 6 | 6 (60.0%) | 2 (40.0%) | 7 (26.9%) | 12 (32.4%) |
| **Risk of bias** | Lower | 4 (40.0%) | 1 (20.0%) | 18 (69.2%) | 19 (51.4%) |
| | Moderate | 5 (50.0%) | 4 (80.0%) | 6 (23.1%) | 15 (40.5%) |
| | Higher | 1 (10.0%) | 0 (0.0%) | 2 (7.7%) | 3 (8.1%) |

[1]The same study is included in more than one subcategory when it reports across different variable levels or multiple variable levels are relevant.

[2] Midpoint year between start and end dates of data collection. If dates were not reported, year of publication was used as a proxy.

[3] Sample size defined as the number of symptomatic individuals tested in the study. If a study used different samples across tests, the largest sample size is reported.

**Table S12: Summary of participant and study characteristics for studies included in the analysis to assess the effect of age, among youth < 25 years and adults ≥ 25 years.**

| | | Vaginal discharge (N=17) | Urethral discharge (N=7) | Genital ulcer (N=7) | All symptoms (N=24) |
|---|---|---|---|---|---|
| **Population characteristics** | | | | | |
| **Population group**[1] | Symptomatic clinic attendee | 14 (82.4%) | 6 (85.7%) | 6 (85.7%) | 20 (83.3%) |
| | General | 1 (5.9%) | - | 1 (14.3%) | 2 (8.3%) |
| | Higher-risk general | - | - | - | - |
| | Key populations | 1 (5.9%) | - | - | 1 (4.2%) |
| | Youth | 15 (88.2%) | 7 (100.0%) | 6 (85.7%) | 20 (83.3%) |
| **Sex** | Male | - | 7 (100.0%) | 1 (14.3%) | 4 (16.7%) |
| | Female | 17 (100.0%) | - | 1 (14.3%) | 14 (58.3%) |
| | Mixed | - | - | 5 (71.4%) | 6 (25%) |
| **Age group**[1] | < 25 years | 15 (88.2%) | 7 (100.0%) | 6 (85.7%) | 21 (87.5%) |
| | ≥ 25 years | 13 (76.5%) | 3 (42.9%) | 4 (57.1%) | 18 (75%) |
| **Study characteristics** | | | | | |
| **Region**[1] | Central Africa | - | - | - | - |
| | Eastern Africa | 6 (35.3%) | 5 (71.4%) | 5 (71.4%) | 11 (45.8%) |
| | Southern Africa | 1 (5.9%) | 1 (14.3%) | 1 (14.3%) | 1 (4.2%) |
| | Western Africa | 10 (58.8%) | 1 (14.3%) | - | 11 (45.8%) |
| | Multiple (not stratified) | - | - | 1 (14.3%) | 1 (4.2%) |
| **Study midpoint year**[1,2] | <1990 | 1 (5.9%) | 1 (14.3%) | - | 2 (8.3%) |
| | 1990 – 1999 | 2 (11.8%) | 3 (42.9%) | 1 (14.3%) | 3 (12.5%) |
| | 2000 – 2009 | 10 (58.8%) | 3 (42.9%) | 5 (71.4%) | 14 (58.3%) |
| | 2010 – 2022 | 5 (29.4%) | 1 (14.3%) | 2 (28.6%) | 6 (25%) |
| **Study design** | Cross sectional | 14 (82.4%) | 7 (100%) | 4 (57.1%) | 18 (75%) |
| | Cohort baseline | 2 (11.8%) | - | 1 (14.3%) | 3 (12.5%) |
| | RCT baseline | 1 (5.9%) | - | 2 (28.6%) | 3 (12.5%) |
| **Sampling method** | Non-probability based | 13 (76.5%) | 5 (71.4%) | 6 (85.7%) | 20 (83.3%) |
| | Probability based | 2 (11.8%) | 2 (28.6%) | 1 (14.3%) | 2 (8.3%) |
| | NR | 2 (11.8%) | - | - | 2 (8.3%) |
| **Sample size**[3] | < 100 | 4 (23.5%) | 5 (71.4%) | 3 (42.9%) | 7 (29.2%) |
| | ≥ 100 | 13 (76.5%) | 2 (28.6%) | 4 (57.1%) | 17 (70.8%) |
| **Number pathogens extracted per study** | 1 | 5 (29.4%) | 3 (42.9%) | 2 (28.6%) | 6 (25%) |
| | 2 - 3 | 5 (29.4%) | 3 (42.9%) | 1 (14.3%) | 8 (33.3%) |
| | 4 - 6 | 7 (41.2%) | 1 (14.3%) | 4 (57.1%) | 10 (41.7%) |
| **Risk of bias** | Lower | 4 (23.5%) | 3 (42.9%) | 4 (57.1%) | 9 (37.5%) |
| | Moderate | 11 (64.7%) | 3 (42.9%) | 2 (28.6%) | 13 (54.2%) |
| | Higher | 2 (11.8%) | 1 (14.3%) | 1 (14.3%) | 2 (8.3%) |

[1]The same study is included in more than one subcategory when it reports across different variable levels or multiple variable levels are relevant.

[2] Mid-point year between start and end dates of data collection. If dates were not reported, year of publication was used as a proxy.

[3] Sample size defined as the number of symptomatic individuals tested in the study. If a study used different samples across tests, the largest sample size is reported.

**Table S13: Generalized linear mixed-effects models on the odds of vaginal discharge, urethral discharge, and genital ulcer among adults of mixed or unmeasured HIV status in sub-Saharan Africa.**

**A. Vaginal discharge**

| Variable | aOR (95% CI) |
|---|---|
| BV | 1.08 (0.51-2.29) |
| CA | 0.45 (0.13-1.54) |
| CS | 1.56 (0.65-3.74) |
| CT | 0.21 (0.10-0.43) *** |
| MG | 0.06 (0.02-0.15) *** |
| NG | 0.09 (0.04-0.18) *** |
| TV | 0.17 (0.08-0.33) *** |
| None | 0.34 (0.14-0.81) * |
| BV:year | 1.01 (0.96-1.06) |
| CA:year | 1.01 (0.95-1.06) |
| CS:year | 1.10 (1.05-1.16) *** |
| CT:year | 1.04 (1.00-1.08) |
| MG:year | 0.99 (0.90-1.10) |
| NG:year | 0.99 (0.96-1.03) |
| TV:year | 0.94 (0.91-0.97) *** |
| None:year | 1.00 (0.94-1.05) |
| **Random effects** | |
| $\sigma_{00}$ region:sti | 0.51 |
| $\sigma_{00}$ study | 0.96 |
| $\sigma_{11}$ region:sti.year | 0.02 |
| $\rho_{01}$ region:sti | 0.53 |
| N region | 3 |
| N observation | 337 |

**B. Urethral discharge**

| Variable | aOR (95% CI) |
|---|---|
| CT | 0.29 (0.20-0.42) *** |
| MG | 0.06 (0.04-0.09) *** |
| NG | 3.80 (2.70-5.35) *** |
| TV | 0.03 (0.02-0.04) *** |
| None | 0.11 (0.07-0.17) *** |
| CT:year | 1.02 (0.99-1.06) |
| MG:year | 0.95 (0.91-1.00) * |
| NG:year | 1.02 (1.00-1.04) |
| TV:year | 0.93 (0.90-0.95) *** |
| None:year | 0.95 (0.91-0.99) * |
| **Random effects** | |
| $\sigma_{00}$ region:sti | 0.04 |
| $\sigma_{00}$ study | 0.87 |
| $\sigma_{11}$ region:sti.year | 0.01 |
| $\rho_{01}$ region:sti | -0.99 |
| N region | 3 |
| N observation | 216 |

**C. Genital ulcer**

| Variable | aOR (95% CI) |
|---|---|
| HD | 0.01 (0.01-0.01) *** |
| HSV | 1.98 (1.20-3.26) ** |
| HSV-1 | 0.01 (0.01-0.03) *** |
| HSV-2 | 1.33 (0.90-1.98) |
| LGV | 0.02 (0.01-0.03) *** |
| TP | 0.10 (0.07-0.13) *** |
| None | 0.60 (0.43-0.84) ** |
| HD:year | 0.83 (0.82-0.85) *** |
| HSV:year | 1.09 (1.07-1.12) *** |
| HSV-1:year | 0.98 (0.92-1.04) |
| HSV-2:year | 1.02 (0.99-1.05) |
| LGV:year | 0.92 (0.88-0.95) *** |
| TP:year | 0.97 (0.96-0.99) *** |
| None:year | 1.02 (1.00-1.04) * |
| HD:male | 1.89 (1.24-2.89) ** |
| HSV:male | 0.71 (0.42-1.19) |
| HSV-1:male | 1.44 (0.55-3.80) |
| HSV-2:male | 0.67 (0.40-1.12) |
| LGV:male | 0.92 (0.45-1.87) |
| TP:male | 1.19 (0.82-1.72) |
| None:male | 0.79 (0.52-1.21) |
| HD:male and female | 1.54 (0.85-2.77) |
| HSV:male and female | 0.99 (0.46-2.14) |
| HSV-1:male and female | 1.04 (0.27-4.04) |
| HSV-2:male and female | 1.12 (0.43-2.89) |
| LGV:male and female | 0.50 (0.11-2.36) |
| TP:male and female | 0.72 (0.43-1.20) |
| None:male and female | 0.88 (0.50-1.54) |
| **Random effects** | |
| $\sigma_{00}$ region:sti | 0.00 |
| $\sigma_{00}$ study | 0.80 |
| $\sigma_{11}$ region:sti.year | 0.00 |
| $\rho_{01}$ region:sti | 0.68 |
| N region | 3 |
| N observation | 473 |

Midpoint year of study is centred at 2015. aOR: adjusted odds ratio, 95% CI: 95% confidence interval, $\sigma_{00}$: random intercept standard deviation, $\sigma_{11}$: random slope standard deviation, $\rho_{01}$: correlation between intercept and slope, N: number, * $p < 0.05$, ** $p < 0.01$, *** $p < 0.001$.

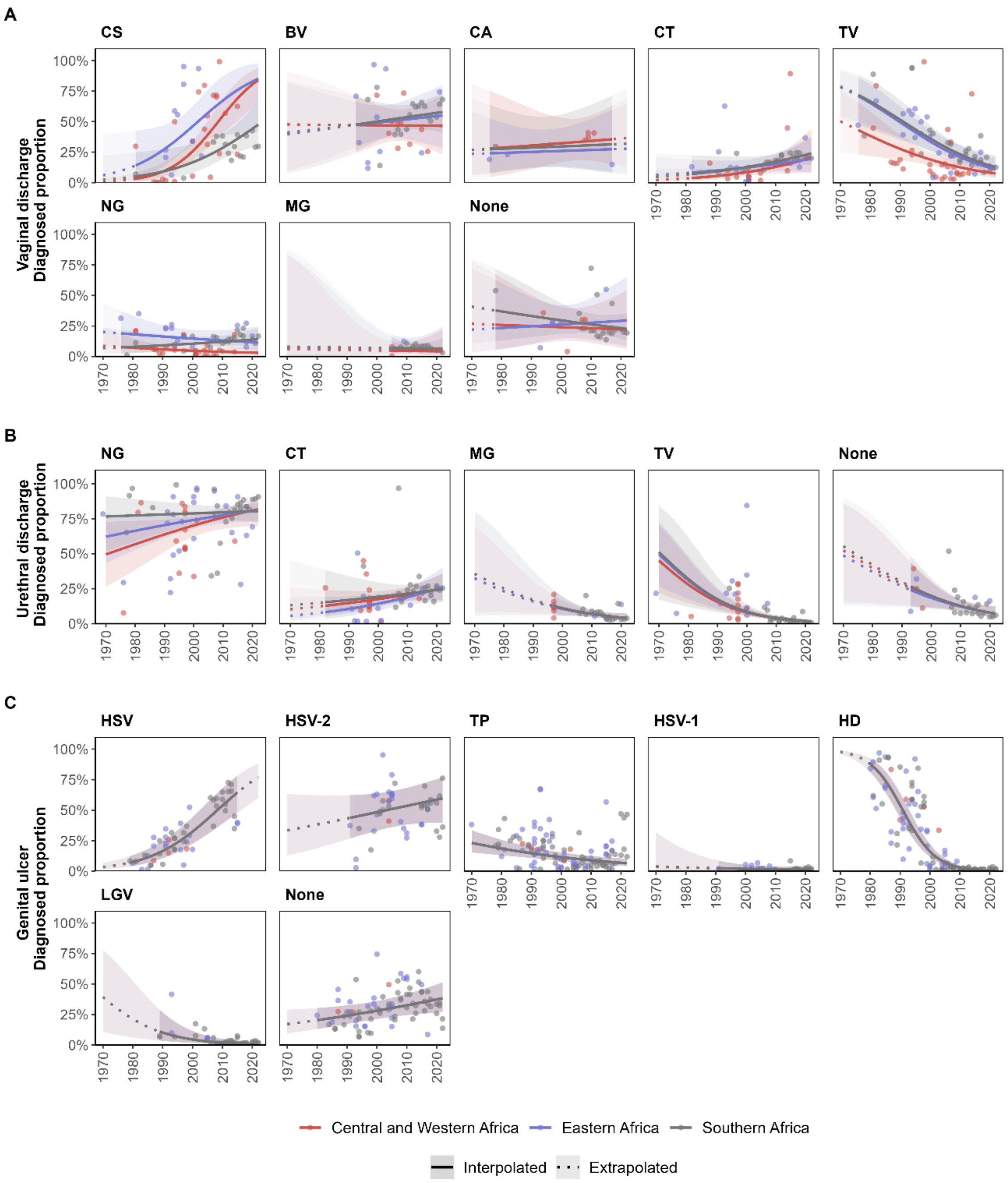

**Figure S2: Estimated diagnosed proportion per pathogen among symptomatic adults of mixed HIV-status in sub-Saharan Africa, by region over time.**

Diagnosed proportion per pathogen among individuals symptomatic with (A) vaginal discharge, (B) urethral discharge, and (C) genital ulcer. Proportions estimated using generalised linear mixed-effects models for each symptom. Lines and shaded areas represent sex-matched population-weighted mean proportions and 95% confidence intervals Solid lines and darker shading denote estimates and confidence intervals within the observed data range (interpolated), while dotted lines and lighter shading indicate estimates and confidence intervals beyond that time frame (extrapolated). Points represent study observations adjusted for diagnostic test performance.

**Table S14: Generalized linear mixed-effects models on the odds of vaginal discharge, urethral discharge, and genital ulcer among adults of known HIV status in sub-Saharan Africa.**

**A. Vaginal discharge**

| Variable | aOR (95% CI) |
|---|---|
| BV | 0.61 (0.35-1.04) |
| CS | 0.55 (0.29-1.02) |
| CT | 0.16 (0.09-0.28) *** |
| MG | 0.04 (0.02-0.08) *** |
| NG | 0.10 (0.06-0.18) *** |
| TV | 0.11 (0.06-0.19) *** |
| None | 0.19 (0.10-0.39) *** |
| BV:year | 1.02 (0.99-1.05) |
| CS:year | 0.97 (0.93-1.01) |
| CT:year | 1.04 (1.01-1.07) ** |
| MG:year | 1.00 (0.96-1.04) |
| NG:year | 1.02 (0.99-1.05) |
| TV:year | 0.94 (0.92-0.97) *** |
| None:year | 1.00 (0.97-1.03) |
| BV:HIV-positive | 1.78 (1.33-2.38) *** |
| CS:HIV-positive | 0.56 (0.38-0.82) ** |
| CT:HIV-positive | 0.92 (0.68-1.25) |
| MG:HIV-positive | 2.28 (1.56-3.33) *** |
| NG:HIV-positive | 1.64 (1.21-2.22) ** |
| TV:HIV-positive | 2.15 (1.61-2.87) *** |
| None:HIV-positive | 0.74 (0.54-1.00) |
| **Random effects** | |
| $\sigma_{00}$ region:sti | 0.39 |
| $\sigma_{00}$ study | 0.41 |
| N region | 3 |
| N observation | 267 |

**B. Urethral discharge**

| Variable | aOR (95% CI) |
|---|---|
| CT | 0.17 (0.06-0.49) ** |
| MG | 0.10 (0.04-0.24) *** |
| NG | 6.86 (2.69-17.52) *** |
| TV | 0.02 (0.01-0.06) *** |
| None | 0.09 (0.03-0.28) *** |
| CT:year | 0.99 (0.95-1.03) |
| MG:year | 0.94 (0.89-0.98) ** |
| NG:year | 1.08 (1.04-1.11) *** |
| TV:year | 0.89 (0.84-0.94) *** |
| None:year | 0.91 (0.88-0.95) *** |
| CT:HIV-positive | 0.55 (0.38-0.79) ** |
| MG:HIV-positive | 1.03 (0.67-1.58) |
| NG:HIV-positive | 1.15 (0.80-1.68) |
| TV:HIV-positive | 1.83 (1.07-3.14) * |
| None:HIV-positive | 1.29 (0.86-1.94) |
| **Random effects** | |
| $\sigma_{00}$ region:sti | 0.56 |
| $\sigma_{00}$ study | 0.49 |
| N region | 2 |
| N observation | 169 |

**C. Genital ulcer**

| Variable | aOR (95% CI) |
|---|---|
| HD | 0.02 (0.02-0.04) *** |
| HSV | 1.08 (0.66-1.75) |
| HSV-1 | 0.03 (0.01-0.07) *** |
| HSV-2 | 1.19 (0.80-1.77) |
| LGV | 0.04 (0.02-0.07) *** |
| TP | 0.11 (0.08-0.16) *** |
| None | 0.73 (0.53-1.01) |
| HD:year | 0.86 (0.84-0.88) *** |
| HSV:year | 1.06 (1.04-1.09) *** |
| HSV-1:year | 1.02 (0.91-1.14) |
| HSV-2:year | 1.00 (0.98-1.03) |
| LGV:year | 0.98 (0.93-1.04) |
| TP:year | 0.99 (0.98-1.01) |
| None:year | 1.01 (0.99-1.03) |
| HD:male | 1.63 (1.06-2.49) * |
| HSV:male | 1.04 (0.66-1.63) |
| HSV-1:male | 0.78 (0.30-2.02) |
| HSV-2:male | 0.46 (0.29-0.74) ** |
| LGV:male | 0.71 (0.38-1.32) |
| TP:male | 1.38 (0.96-2.00) |
| None:male | 0.85 (0.60-1.21) |
| HD:male and female | 0.98 (0.58-1.65) |
| HSV:male and female | 0.86 (0.43-1.74) |
| HSV-1:male and female | 1.27 (0.22-7.45) |
| HSV-2:male and female | 0.67 (0.36-1.24) |
| LGV:male and female | 1.91 (0.50-7.31) |
| TP:male and female | 1.25 (0.76-2.03) |
| None:male and female | 0.72 (0.44-1.17) |
| HD:HIV-positive | 0.80 (0.56-1.14) |
| HSV:HIV-positive | 1.68 (1.10-2.55) * |
| HSV-1:HIV-positive | 1.18 (0.51-2.73) |
| HSV-2:HIV-positive | 1.67 (1.12-2.48) * |
| LGV:HIV-positive | 0.92 (0.51-1.67) |
| TP:HIV-positive | 0.94 (0.68-1.29) |
| None:HIV-positive | 0.64 (0.47-0.88) ** |
| **Random effects** | |
| $\sigma_{00}$ region:sti | <0.001 |
| $\sigma_{00}$ study | 0.67 |
| N region | 3 |
| N observation | 492 |

Midpoint year of study is centred at 2015. aOR: adjusted odds ratio, 95% CI: 95% confidence interval, $\sigma_{00}$: random intercept standard deviation, N: number, * $p < 0.05$, ** $p < 0.01$, *** $p < 0.001$.

**Table S15: Generalized linear mixed-effects models on the odds of vaginal discharge, urethral discharge, and genital ulcer among youth under 25 years and adults 25 years and older in sub-Saharan Africa.**

**A. Vaginal discharge**

| Variable | aOR (95% CI) |
|---|---|
| BV | 0.69 (0.29-1.60) |
| CA | 0.49 (0.07-3.51) |
| CS | 0.49 (0.19-1.24) |
| CT | 0.08 (0.03-0.19) *** |
| MG | 0.04 (0.01-0.13) *** |
| NG | 0.05 (0.02-0.13) *** |
| TV | 0.11 (0.04-0.25) *** |
| None | 0.36 (0.13-0.97) * |
| BV:year | 1.05 (1.02-1.09) ** |
| CA:year | 1.05 (0.96-1.14) |
| CS:year | 1.01 (0.96-1.05) |
| CT:year | 1.04 (1.01-1.08) * |
| MG:year | 0.99 (0.95-1.03) |
| NG:year | 1.00 (0.97-1.03) |
| TV:year | 0.92 (0.89-0.95) *** |
| None:year | 0.98 (0.95-1.02) |
| BV:<25 years | 1.13 (0.84-1.52) |
| CA:<25 years | 2.16 (0.75-6.26) |
| CS:<25 years | 1.83 (1.26-2.67) ** |
| CT:<25 years | 2.40 (1.73-3.32) *** |
| MG:<25 years | 1.99 (1.33-2.98) *** |
| NG:<25 years | 1.66 (1.20-2.28) ** |
| TV:<25 years | 0.85 (0.63-1.13) |
| None:<25 years | 0.67 (0.49-0.91) * |
| **Random effects** | |
| $\sigma_{00}$ region:sti | 0.68 |
| $\sigma_{00}$ study | 0.42 |
| N region | 3 |
| N observation | 281 |

**B. Urethral discharge**

| Variable | aOR (95% CI) |
|---|---|
| CT | 0.28 (0.23-0.34) *** |
| MG | 0.06 (0.04-0.07) *** |
| NG | 4.51 (3.70-5.51) *** |
| TV | 0.02 (0.02-0.03) *** |
| None | 0.10 (0.08-0.12) *** |
| CT:year | 1.04 (1.01-1.07) ** |
| MG:year | 0.92 (0.89-0.96) *** |
| NG:year | 1.12 (1.10-1.15) *** |
| TV:year | 0.89 (0.85-0.93) *** |
| None:year | 0.90 (0.87-0.94) *** |
| CT:<25 years | 1.60 (1.20-2.15) ** |
| MG:<25 years | 1.02 (0.70-1.47) |
| NG:<25 years | 1.23 (0.91-1.66) |
| TV:<25 years | 1.35 (0.83-2.18) |
| None:<25 years | 0.74 (0.53-1.04) |
| **Random effects** | |
| $\sigma_{00}$ region:sti | <0.001 |
| $\sigma_{00}$ study | 0.35 |
| N region | 3 |
| N observation | 161 |

**C. Genital ulcer**

| Variable | aOR (95% CI) |
|---|---|
| HD | 0.02 (0.01-0.04) *** |
| HSV | 1.26 (0.79-2.02) |
| HSV-1 | 0.01 (0.00-0.05) *** |
| HSV-2 | 1.44 (1.05-1.96) * |
| LGV | 0.02 (0.01-0.04) *** |
| TP | 0.08 (0.06-0.12) *** |
| None | 0.57 (0.46-0.72) *** |
| HD:year | 1.02 (0.96-1.08) |
| HSV:year | 0.99 (0.91-1.07) |
| HSV-1:year | 1.09 (0.88-1.37) |
| HSV-2:year | 0.99 (0.94-1.03) |
| LGV:year | 1.02 (0.95-1.10) |
| TP:year | 1.10 (1.06-1.14) *** |
| None:year | 0.97 (0.94-1.00) * |
| HD:Male | 0.65 (0.36-1.18) |
| HSV:Male | 0.88 (0.60-1.30) |
| HSV-1:Male | 0.79 (0.29-2.14) |
| HSV-2:Male | 0.63 (0.42-0.95) * |
| LGV:Male | 0.76 (0.41-1.43) |
| TP:Male | 1.99 (1.36-2.90) *** |
| None:Male | 0.93 (0.70-1.22) |
| HD:<25 years | 2.43 (1.34-4.39) ** |
| HSV:<25 years | 1.01 (0.68-1.51) |
| HSV-1:<25 years | 2.62 (0.90-7.61) |
| HSV-2:<25 years | 1.12 (0.73-1.72) |
| LGV:<25 years | 2.37 (1.25-4.49) ** |
| TP:<25 years | 1.32 (0.88-1.97) |
| None:<25 years | 1.08 (0.81-1.45) |
| **Random effects** | |
| $\sigma_{00}$ region:sti | <0.001 |
| $\sigma_{00}$ study | 0.36 |
| N region | 3 |
| N observation | 274 |

Midpoint year of study is centred at 2015. aOR: adjusted odds ratio, 95% CI: 95% confidence interval, $\sigma_{00}$: random intercept standard deviation, N: number, * p < 0.05, ** p < 0.01, *** p < 0.001.

**Table S16: Estimated population-weighted aetiologic proportions for 2015 in sub-Saharan Africa**

| Pathogen | Overall | | | HIV positive | | | HIV negative | | | < 25 years | | | ≥ 25 years | | |
|---|---|---|---|---|---|---|---|---|---|---|---|---|---|---|---|
| | No. study | No. obs | Proportion % (95% CI) | No. study | No. obs | Proportion % (95% CI) | No. study | No. obs | Proportion % (95% CI) | No. study | No. obs | Proportion % (95% CI) | No. study | No. obs | Proportion % (95% CI) |
| **A. Vaginal discharge** | | | | | | | | | | | | | | | |
| CS | 31 | 50 | 69.4 (44.1-86.6) | 4 | 17 | 24.0 (13.5-39.0) | 3 | 16 | 36.2 (22.1-53.1) | 5 | 18 | 51.2 (32.3-69.7) | 6 | 19 | 36.4 (21.0-55.2) |
| BV | 23 | 39 | 50.0 (32.3-67.8) | 6 | 20 | 47.4 (35.7-59.4) | 7 | 21 | 33.6 (23.7-45.2) | 6 | 21 | 39.6 (27.8-52.8) | 7 | 21 | 36.8 (25.3-50.1) |
| CA | 9 | 9 | 31.5 (12.4-59.9) | 0 | 0 | - | 0 | 0 | - | 2 | 2 | 51.8 (14.7-87.0) | 2 | 2 | 33.2 (7.7-74.7) |
| CT | 29 | 49 | 16.5 (8.7-29.0) | 5 | 19 | 11.1 (6.3-18.7) | 6 | 20 | 11.9 (6.9-19.7) | 6 | 20 | 11.6 (5.5-22.8) | 4 | 18 | 5.2 (2.4-11.1) |
| TV | 55 | 80 | 12.9 (7.7-20.7) | 8 | 22 | 17.0 (11.5-24.6) | 8 | 22 | 8.7 (5.7-13.2) | 11 | 25 | 6.3 (3.7-10.5) | 11 | 25 | 7.4 (4.4-12.1) |
| NG | 37 | 62 | 6.6 (3.2-13.0) | 6 | 20 | 12.9 (7.9-20.4) | 6 | 20 | 8.3 (5.0-13.6) | 7 | 21 | 5.2 (2.5-10.5) | 8 | 22 | 3.2 (1.5-6.6) |
| MG | 5 | 20 | 5.4 (1.8-14.8) | 3 | 17 | 8.6 (4.6-15.4) | 4 | 18 | 3.9 (2.1-7.4) | 1 | 15 | 6.8 (1.9-21.2) | 2 | 16 | 3.5 (1.0-11.6) |
| None | 13 | 28 | 25.1 (10.6-48.5) | 3 | 17 | 10.5 (4.6-22.0) | 4 | 18 | 13.7 (6.3-27.4) | 4 | 18 | 20.5 (9.0-40.2) | 4 | 18 | 27.8 (13.0-49.9) |
| **B. Urethral discharge** | | | | | | | | | | | | | | | |
| NG | 46 | 67 | 78.8 (70.9-85.1) | 3 | 17 | 90.8 (80.6-95.9) | 4 | 19 | 89.5 (78.7-95.1) | 4 | 18 | 84.8 (81.4-87.6) | 3 | 17 | 81.9 (78.7-84.6) |
| CT | 27 | 48 | 22.2 (16.0-30.1) | 3 | 17 | 5.1 (1.6-15.3) | 4 | 19 | 9.0 (2.9-24.3) | 3 | 17 | 31.2 (26.7-36.1) | 1 | 15 | 22.0 (18.8-25.6) |
| MG | 8 | 29 | 5.6 (3.7-8.2) | 2 | 16 | 13.0 (6.4-24.7) | 2 | 17 | 12.7 (6.3-23.8) | 1 | 15 | 5.4 (4.0-7.1) | 1 | 15 | 5.3 (4.2-6.6) |
| TV | 25 | 46 | 2.8 (1.8-4.2) | 2 | 16 | 3.8 (1.4-9.8) | 2 | 17 | 2.1 (0.8-5.5) | 4 | 18 | 3.1 (2.1-4.5) | 2 | 16 | 2.3 (1.6-3.2) |
| None | 11 | 26 | 10.0 (6.9-14.2) | 1 | 15 | 10.3 (2.8-31.2) | 1 | 16 | 8.2 (2.2-25.8) | 1 | 15 | 6.9 (5.3-8.9) | 1 | 15 | 9.1 (7.5-11.0) |
| **C. Genital ulcer** | | | | | | | | | | | | | | | |
| HSV | 27 | 54 | 64.4 (47.8-78.1) | 10 | 28 | 62.9 (48.3-75.4) | 7 | 23 | 50.2 (35.4-65.0) | 1 | 12 | 54.5 (42.0-66.4) | 1 | 15 | 54.2 (42.7-65.3) |
| HSV-2 | 21 | 46 | 56.1 (39.2-71.6) | 13 | 28 | 57.3 (45.7-68.1) | 12 | 27 | 44.5 (33.4-56.3) | 2 | 10 | 56.2 (46.0-66.0) | 2 | 15 | 53.4 (45.3-61.2) |
| TP | 62 | 115 | 7.8 (5.3-11.4) | 20 | 55 | 11.5 (8.1-16.2) | 19 | 51 | 12.2 (8.5-17.2) | 4 | 27 | 13.2 (9.5-18.2) | 1 | 29 | 10.4 (7.9-13.5) |
| HSV-1 | 13 | 30 | 1.4 (0.5-3.7) | 5 | 16 | 3.5 (1.1-10.5) | 4 | 15 | 3.0 (0.9-9.4) | 1 | 7 | 3.4 (1.1-9.7) | 1 | 12 | 1.3 (0.4-4.1) |
| LGV | 10 | 42 | 1.3 (0.4-3.5) | 3 | 28 | 4.4 (1.8-10.6) | 3 | 27 | 4.8 (1.9-11.3) | 1 | 18 | 4.8 (2.7-8.2) | 1 | 25 | 2.1 (1.2-3.4) |
| HD | 54 | 105 | 1.3 (0.7-2.1) | 21 | 54 | 2.2 (1.4-3.4) | 21 | 52 | 2.7 (1.7-4.2) | 2 | 23 | 4.5 (2.7-7.5) | 1 | 29 | 1.9 (1.2-3.1) |
| None | 36 | 81 | 34.9 (25.6-45.5) | 13 | 46 | 27.6 (20.6-36.0) | 12 | 42 | 37.4 (28.8-47.0) | 2 | 23 | 37.4 (31.2-44.1) | 1 | 29 | 35.6 (30.7-40.8) |

No. study = number of unique studies; No. obs = number of observations. Observations refers to the estimates extracted from a study; multiple observations are possible when study outcomes are stratified by population type, country, year or time-period, sex, HIV status, or age (as available).

**Table S17: Risk of bias assessment for reports included in the review.**

**Assessment:** ● Yes = 1 point ● Unclear = 0 point ● No = 0 point

| Reference | Year[1] | Country | Q1 | Q2 | Q3 | Q4 | Q5 | Q6 | Q7 | Q8 | Q9 | Q10 | Score | Rating |
|---|---|---|---|---|---|---|---|---|---|---|---|---|---|---|
| Arya 1972[45] | 1969 | Uganda | Yes | Yes | No | Yes | Yes | Yes | Yes | Yes | No | Yes | 8 | Lower |
| Ongom 1971[46] | 1970 | Uganda | No | No | No | Yes | Unclear | No | No | Yes | No | No | 2 | Higher |
| Van Binsbergen 1980[47] | 1976 | Kenya | Yes | Yes | No | Yes | Unclear | No | Yes | Yes | Yes | Yes | 7 | Moderate |
| Fakunle 1976[48] | 1976* | Nigeria | No | Yes | No | Unclear | Unclear | Yes | Yes | Yes | No | Yes | 5 | Moderate |
| Ballard 1977[49] | 1977* | South Africa | Yes | Yes | No | Unclear | Unclear | Unclear | Yes | Yes | No | No | 4 | Higher |
| Rubin 1980[50] | 1978 | South Africa | No | No | No | Unclear | Unclear | Yes | Yes | Yes | Yes | No | 4 | Higher |
| Meheus 1982[51] | 1978 | Eswatini | Yes | Yes | No | Yes | Unclear | Yes | No | Yes | No | Yes | 6 | Moderate |
| Meheus 1983[52] | 1979 | Eswatini | Yes | Yes | No | Yes | Yes | Yes | Yes | Yes | No | Yes | 8 | Lower |
| Ursi 1981[53] | 1979 | Eswatini | No | Yes | No | Yes | Unclear | Yes | Yes | Yes | No | Yes | 6 | Moderate |
| Meheus 1980[54] | 1979 | Eswatini | Yes | Yes | No | Yes | Yes | Yes | No | Yes | No | Yes | 7 | Moderate |
| Nsanze 1981[55] | 1980 | Kenya | Yes | Yes | No | No | Yes | Yes | No | Yes | Yes | Yes | 7 | Moderate |
| Fast 1983[56] | 1981 | Kenya | No | No | No | Unclear | Unclear | Yes | Yes | No | No | No | 2 | Higher |
| Yvert 1984[57] | 1981 | Gabon | Yes | Yes | No | Unclear | Unclear | No | Yes | Yes | Yes | Yes | 6 | Moderate |
| Hoosen 1981[58] | 1981* | South Africa | No | Yes | No | No | Yes | No | Yes | Yes | Yes | No | 5 | Moderate |
| Crewe-Brown 1982[59] | 1981 | South Africa | Yes | Yes | No | Yes | Yes | Yes | Yes | No | Yes | No | 7 | Moderate |
| D'Costa 1985[60] | 1982 | Kenya | No | Yes | No | Unclear | Unclear | Yes | No | Yes | No | No | 3 | Higher |
| Mirza 1983[61] | 1982 | Kenya | Yes | Yes | No | Yes | Yes | No | Yes | Yes | Yes | Yes | 8 | Lower |
| Mabey 1982[62] | 1982* | The Gambia | No | Yes | No | Yes | Yes | Yes | No | Yes | No | Yes | 6 | Moderate |
| Mabey 1984[63] | 1982 | The Gambia | No | Yes | No | Yes | Yes | Yes | No | Yes | No | No | 5 | Moderate |
| Plummer 1983[64] | 1983* | Kenya | No | Yes | No | Unclear | Yes | Yes | No | Yes | No | No | 4 | Higher |
| Mauff 1983[65] | 1983* | South Africa | Yes | Yes | No | No | Yes | Yes | No | Yes | No | Yes | 6 | Moderate |
| Coovadia 1985[66] | 1984 | South Africa | Yes | Yes | No | Unclear | Yes | Yes | Yes | Yes | Yes | No | 7 | Moderate |
| Greenblatt 1988[67] | 1985 | Kenya | No | Yes | Yes | Yes | Yes | Yes | Yes | Yes | Yes | Yes | 9 | Lower |
| Plummer 1985[68] | 1985* | Kenya | Yes | Yes | No | No | Yes | Yes | No | Yes | Yes | No | 6 | Moderate |
| Kreiss 1986[69] | 1985 | Kenya | No | Yes | Yes | Unclear | Unclear | Yes | No | Yes | No | Yes | 5 | Moderate |
| Dangor 1989[70] | 1986 | South Africa | Yes | Yes | No | Yes | Unclear | Yes | Yes | No | Yes | Yes | 7 | Moderate |
| Ismail 1990[71] | 1986 | Somalia | Yes | Yes | Yes | Yes | No | No | No | Yes | No | No | 5 | Moderate |
| Meiring 1989[72] | 1986 | South Africa | Yes | No | No | Unclear | Unclear | No | Yes | Yes | No | No | 3 | Higher |
| Mabey 1987[73] | 1987* | The Gambia | Yes | Yes | No | Unclear | Yes | Yes | Yes | Yes | Yes | Yes | 8 | Lower |
| Leclerc 1988[74] | 1988* | Gabon | Yes | Yes | No | Unclear | Yes | No | Yes | Yes | No | Yes | 6 | Moderate |
| Bogaerts 1998[75] | 1989 | Rwanda | Yes | Yes | Yes | Yes | Yes | Yes | Yes | Yes | Yes | No | 9 | Lower |
| Faye-Kette 1993[76] | 1989 | Côte d'Ivoire | Yes | Yes | No | Unclear | Unclear | No | Yes | Yes | Yes | No | 5 | Moderate |
| O'Farrell 1994[77] | 1989 | South Africa | No | Yes | No | Unclear | Unclear | Yes | Yes | Yes | Yes | No | 5 | Moderate |
| Daly 1994[78] | 1990 | Kenya | No | Yes | Yes | Yes | Yes | Yes | No | Yes | No | No | 6 | Moderate |

| Reference | Year[1] | Country | Q1 | Q2 | Q3 | Q4 | Q5 | Q6 | Q7 | Q8 | Q9 | Q10 | Score | Rating |
|---|---|---|---|---|---|---|---|---|---|---|---|---|---|---|
| Thior 1997[79] | 1990 | Senegal | 🔴 | 🟢 | 🟢 | 🟢 | 🟡 | 🟡 | 🟢 | 🟢 | 🔴 | 🟢 | 6 | Moderate |
| Ndinya-Achola 1996[80] | 1991 | Kenya | 🔴 | 🟢 | 🔴 | 🟡 | 🟡 | 🟢 | 🟢 | 🟢 | 🔴 | 🔴 | 4 | Higher |
| Kamya 1995[81] | 1991 | Uganda | 🟢 | 🟢 | 🟢 | 🟢 | 🟢 | 🟢 | 🟢 | 🟢 | 🟢 | 🔴 | 9 | Lower |
| Hanson 1996[82] | 1991 | Zambia | 🔴 | 🟢 | 🟢 | 🟢 | 🟡 | 🟢 | 🟢 | 🟢 | 🟢 | 🟢 | 8 | Lower |
| LeBacq 1993[83] | 1991 | Zimbabwe | 🟢 | 🟢 | 🟢 | 🟢 | 🔴 | 🟢 | 🔴 | 🟢 | 🟢 | 🟢 | 8 | Lower |
| Kimani 1995[84] | 1992 | Kenya | 🔴 | 🟢 | 🟢 | 🟢 | 🟢 | 🟢 | 🟢 | 🟢 | 🔴 | 🔴 | 7 | Moderate |
| Oni 1997[85] | 1992 | Nigeria | 🟢 | 🟢 | 🔴 | 🟢 | 🟡 | 🟢 | 🔴 | 🟢 | 🔴 | 🟢 | 6 | Moderate |
| Totten 2000[86] | 1992 | Senegal | 🟢 | 🔴 | 🔴 | 🟢 | 🟡 | 🟢 | 🔴 | 🟢 | 🟢 | 🟢 | 6 | Moderate |
| Htun 2007[87] | 1992 | South Africa | 🟢 | 🟢 | 🟢 | 🟢 | 🟢 | 🟢 | 🟢 | 🟢 | 🟢 | 🟢 | 10 | Lower |
| Grosskurth 1996[88] | 1992 | Tanzania | 🟢 | 🟢 | 🔴 | 🟢 | 🟢 | 🟢 | 🟢 | 🟢 | 🔴 | 🟢 | 8 | Lower |
| Tswana 1995[89] | 1992 | Zimbabwe | 🟢 | 🟢 | 🟢 | 🟢 | 🟢 | 🟢 | 🟢 | 🟢 | 🔴 | 🔴 | 8 | Lower |
| Ghys 1995[90] | 1993 | Côte d'Ivoire | 🔴 | 🟢 | 🟢 | 🔴 | 🟡 | 🟢 | 🟢 | 🟢 | 🔴 | 🔴 | 5 | Moderate |
| Meless 1997[91] | 1993 | Ethiopia | 🟢 | 🟢 | 🔴 | 🟡 | 🟡 | 🟡 | 🟢 | 🟢 | 🔴 | 🟢 | 5 | Moderate |
| Harms 1994[92] | 1993 | Madagascar | 🟢 | 🟢 | 🟢 | 🔴 | 🟢 | 🟢 | 🟢 | 🔴 | 🟢 | 🟢 | 8 | Lower |
| Dallabetta 1998[93] | 1993 | Malawi | 🟢 | 🟢 | 🔴 | 🟢 | 🔴 | 🟢 | 🟢 | 🟢 | 🔴 | 🔴 | 6 | Moderate |
| Behets 1995[94] | 1993 | Malawi | 🟢 | 🟢 | 🟢 | 🟢 | 🟢 | 🟢 | 🟢 | 🟢 | 🔴 | 🟢 | 9 | Lower |
| Cossa 1994[95] | 1993 | Mozambique | 🔴 | 🟢 | 🟢 | 🟢 | 🟡 | 🟢 | 🔴 | 🟢 | 🔴 | 🔴 | 5 | Moderate |
| Alary 1998[96] | 1994 | Benin | 🔴 | 🟢 | 🔴 | 🟢 | 🟢 | 🔴 | 🟢 | 🟢 | 🟢 | 🔴 | 6 | Moderate |
| Thomas 1996[97] | 1994 | Kenya | 🔴 | 🟢 | 🟢 | 🟢 | 🟢 | 🔴 | 🔴 | 🟢 | 🟢 | 🔴 | 6 | Moderate |
| King 1998[98] | 1994 | Kenya | 🔴 | 🟢 | 🟢 | 🟢 | 🟢 | 🟢 | 🟢 | 🟢 | 🔴 | 🟢 | 8 | Lower |
| Htun 1998[99] | 1994 | Lesotho | 🔴 | 🟢 | 🔴 | 🟢 | 🟢 | 🟢 | 🟢 | 🟢 | 🟢 | 🟢 | 8 | Lower |
| Morse 1997[100] | 1994 | Lesotho | 🔴 | 🟢 | 🟢 | 🟢 | 🟢 | 🟢 | 🟢 | 🟢 | 🟢 | 🟢 | 9 | Lower |
| Lo 1997[101] | 1994 | Mauritania | 🟢 | 🔴 | 🔴 | 🟡 | 🟡 | 🔴 | 🟢 | 🟢 | 🟢 | 🔴 | 4 | Higher |
| Chen 2000[102] | 1994 | South Africa | 🟢 | 🟢 | 🟢 | 🟢 | 🟢 | 🟢 | 🟢 | 🟢 | 🟢 | 🟢 | 10 | Lower |
| Schneider 1998[103] | 1994 | South Africa | 🔴 | 🟢 | 🔴 | 🟢 | 🟡 | 🟢 | 🟢 | 🟢 | 🔴 | 🔴 | 5 | Moderate |
| Pillay 1994[104] | 1994* | South Africa | 🟢 | 🟢 | 🔴 | 🟢 | 🟡 | 🟡 | 🟢 | 🔴 | 🔴 | 🟢 | 5 | Moderate |
| Mayaud 1997[105] | 1994 | Tanzania | 🔴 | 🟢 | 🔴 | 🟢 | 🟡 | 🟢 | 🔴 | 🟢 | 🔴 | 🔴 | 4 | Higher |
| Aseffa 1998[106] | 1995 | Ethiopia | 🔴 | 🟢 | 🟢 | 🟢 | 🟡 | 🟢 | 🔴 | 🟢 | 🔴 | 🟢 | 6 | Moderate |
| MacDonald 2001[107] | 1995 | Kenya | 🔴 | 🟢 | 🟢 | 🟡 | 🟡 | 🟢 | 🟢 | 🟢 | 🔴 | 🟢 | 6 | Moderate |
| Jackson 1997[108] | 1995 | Kenya | 🔴 | 🟢 | 🟢 | 🟢 | 🟡 | 🟢 | 🔴 | 🟢 | 🟢 | 🟢 | 7 | Moderate |
| Gateau 1996[109] | 1995 | Madagascar | 🔴 | 🔴 | 🔴 | 🟢 | 🟢 | 🟢 | 🔴 | 🟢 | 🔴 | 🔴 | 4 | Higher |
| Bakare 2002[110] | 1995 | Nigeria | 🟢 | 🟢 | 🔴 | 🟢 | 🟡 | 🟢 | 🟢 | 🟢 | 🟢 | 🔴 | 7 | Moderate |
| Ramuthaga 1995[111] | 1995* | South Africa | 🔴 | 🔴 | 🔴 | 🟡 | 🟡 | 🟢 | 🟢 | 🔴 | 🔴 | 🟢 | 3 | Higher |
| Ahmed 1995[112] | 1995* | Zambia | 🔴 | 🔴 | 🔴 | 🟡 | 🟡 | 🟢 | 🔴 | 🟢 | 🔴 | 🟢 | 3 | Higher |
| Latif 1999[113] | 1995 | Zimbabwe | 🔴 | 🟢 | 🟢 | 🟢 | 🟢 | 🟢 | 🟢 | 🟢 | 🟢 | 🟢 | 9 | Lower |
| Farina 2000[114] | 1996 | Comoros Islands | 🔴 | 🟢 | 🔴 | 🟡 | 🟢 | 🟢 | 🔴 | 🟢 | 🔴 | 🟢 | 5 | Moderate |
| Malonza 1999[115] | 1996 | Kenya | 🔴 | 🟢 | 🟢 | 🟡 | 🟢 | 🟢 | 🟢 | 🔴 | 🟢 | 🟢 | 7 | Moderate |

| Reference | Year[1] | Country | Q1 | Q2 | Q3 | Q4 | Q5 | Q6 | Q7 | Q8 | Q9 | Q10 | Score | Rating |
|---|---|---|---|---|---|---|---|---|---|---|---|---|---|---|
| Cohen 1997[116] | 1996 | Malawi | Red | Green | Green | Yellow | Yellow | Green | Green | Green | Green | Green | 7 | Moderate |
| Dieye 2003[117] | 1996 | Senegal | Red | Green | Red | Yellow | Red | Red | Red | Green | Red | Red | 2 | Higher |
| Lai 2003[118] | 1996 | South Africa | Green | Green | Green | Green | Green | Green | Green | Green | Green | Green | 10 | Lower |
| Ballard 1996[119] | 1996* | South Africa | Red | Red | Green | Yellow | Yellow | Green | Red | Green | Red | Red | 3 | Higher |
| Pepin 2001[120] | 1997 | Benin, Burkina Faso, Côte d'Ivoire, Ghana, Guinea, Mali, Senegal | Green | Green | Red | Green | Yellow | Green | Green | Green | Green | Green | 8 | Lower |
| Morency 2001[121] | 1997 | Central African Republic | Green | Green | Red | Green | Green | Green | Green | Red | Green | Green | 8 | Lower |
| Gomes 2001[122] | 1997 | Guinea-Bissau | Green | Green | Red | Yellow | Green | Green | Green | Green | Red | Green | 7 | Moderate |
| Fonck 2000[123] | 1997 | Kenya | Red | Green | Green | Green | Yellow | Green | Green | Green | Green | Green | 8 | Lower |
| Ballard 2007[124] | 1997 | Lesotho, South Africa | Green | Green | Green | Yellow | Yellow | Green | Green | Green | Green | Green | 8 | Lower |
| Behets 1999[125] | 1997 | Madagascar | Green | Green | Red | Green | Yellow | Green | Green | Green | Green | Green | 8 | Lower |
| Rajagopal 1999[126] | 1997 | South Africa | Red | Green | Green | Yellow | Yellow | Green | Red | Green | Red | Green | 5 | Moderate |
| Hoosen 1997[127] | 1997* | South Africa | Green | Green | Green | Yellow | Yellow | Red | Red | Green | Green | Red | 5 | Moderate |
| Ramjee 1998[128] | 1997 | South Africa | Red | Green | Green | Red | Green | Green | Red | Green | Red | Red | 5 | Moderate |
| Paz-Bailey 2005[31] | 1998 | Botswana | Green | Green | Green | Green | Yellow | Green | Green | Green | Green | Green | 9 | Lower |
| Nzila 1991[129] | 1998 | Democratic Republic of Congo | Red | Green | Green | Red | Yellow | Green | Red | Green | Red | Red | 4 | Higher |
| Anorlu 2001[130] | 1998 | Nigeria | Green | Green | Red | Yellow | Green | Green | Green | Green | Red | Green | 7 | Moderate |
| Ballard 2002[131] | 1998 | South Africa | Green | Green | Green | Green | Green | Yellow | Green | Green | Green | Green | 9 | Lower |
| Obasi 2001[132] | 1998 | Tanzania | Red | Green | Green | Red | Green | Red | Green | Green | Red | Red | 5 | Moderate |
| Msuya 2006[133] | 1999 | Tanzania | Red | Green | Red | Yellow | Green | Green | Green | Green | Red | Red | 5 | Moderate |
| Msuya 2002[134] | 1999 | Tanzania | Red | Green | Red | Yellow | Green | Green | Green | Green | Red | Red | 5 | Moderate |
| Hoyo 2005[135] | 1999 | Malawi | Green | Green | Red | Green | Green | Green | Green | Green | Green | Green | 9 | Lower |
| Chalamilla 2006[136] | 1999 | Tanzania | Green | Green | Green | Green | Yellow | Green | Red | Green | Green | Red | 7 | Moderate |
| Ahmed 2003[137] | 1999 | Tanzania | Green | Green | Green | Green | Green | Green | Green | Green | Green | Green | 10 | Lower |
| Mbizvo 2004[138] | 1999 | Zimbabwe | Red | Green | Green | Green | Green | Green | Red | Green | Red | Red | 6 | Moderate |
| Vuylsteke 2003[139] | 2000 | Côte d'Ivoire | Red | Green | Red | Yellow | Yellow | Red | Red | Green | Green | Red | 3 | Higher |
| Price 2004[140] | 2000 | Malawi | Green | Green | Green | Green | Green | Green | Green | Red | Red | Red | 7 | Moderate |
| Anorlu 2004[141] | 2000 | Nigeria | Red | Green | Green | Green | Red | Green | Green | Green | Green | Green | 8 | Lower |
| Bakare 2002[142] | 2000 | Nigeria | Green | Green | Red | Green | Yellow | Green | Green | Green | Red | Green | 7 | Moderate |
| Moodley 2002[143] | 2000 | South Africa | Green | Green | Green | Red | Yellow | Yellow | Green | Green | Green | Green | 7 | Moderate |
| Nilsen 2007[144] | 2000 | Tanzania | Green | Green | Green | Green | Green | Green | Green | Green | Green | Green | 10 | Lower |
| Riedner 2007[145] | 2000 | Tanzania | Red | Green | Green | Red | Yellow | Green | Red | Green | Green | Green | 6 | Moderate |
| Watson-Jones 2000[146] | 2000* | Tanzania | Red | Green | Red | Red | Yellow | Green | Red | Red | Red | Green | 3 | Higher |
| Pickering 2005[147] | 2000 | Uganda | Green | Green | Green | Yellow | Green | Green | Green | Green | Green | Red | 8 | Lower |
| Mason 2000[148] | 2000* | Zimbabwe | Red | Green | Red | Yellow | Yellow | Green | Green | Green | Green | Green | 6 | Moderate |
| Wolday 2004[149] | 2001 | Ethiopia | Red | Green | Green | Green | Green | Green | Red | Green | Green | Green | 8 | Lower |

| Reference | Year[1] | Country | Q1 | Q2 | Q3 | Q4 | Q5 | Q6 | Q7 | Q8 | Q9 | Q10 | Score | Rating |
|---|---|---|---|---|---|---|---|---|---|---|---|---|---|---|
| Pepin 2004[150] | 2001 | Benin, Burkina Faso, Ghana, Guinea, Mali | 🟢 | 🟢 | 🔴 | 🟡 | 🟢 | 🟢 | 🟢 | 🟢 | 🟢 | 🟢 | 8 | Lower |
| Romoren 2007[151] | 2001 | Botswana | 🔴 | 🟢 | 🔴 | 🔴 | 🟡 | 🟢 | 🟢 | 🟢 | 🔴 | 🔴 | 4 | Higher |
| Bankole 2001[152] | 2001* | Côte d'Ivoire | 🟢 | 🟢 | 🔴 | 🟡 | 🔴 | 🔴 | 🟢 | 🟢 | 🔴 | 🔴 | 4 | Higher |
| Tadesse 2001[153] | 2001* | Ethiopia | 🔴 | 🟢 | 🔴 | 🔴 | 🟢 | 🟢 | 🟢 | 🟢 | 🔴 | 🟢 | 6 | Moderate |
| Zachariah 2002[154] | 2001 | Malawi | 🟢 | 🟢 | 🔴 | 🟢 | 🟢 | 🔴 | 🟢 | 🟢 | 🔴 | 🟢 | 7 | Moderate |
| Ly 2006[155] | 2001 | Senegal | 🔴 | 🟢 | 🟢 | 🟡 | 🟡 | 🔴 | 🔴 | 🟢 | 🔴 | 🔴 | 3 | Higher |
| Moodley 2003[156] | 2001 | South Africa | 🟢 | 🟢 | 🟢 | 🔴 | 🟡 | 🟢 | 🟢 | 🟢 | 🟢 | 🟢 | 8 | Lower |
| Otuonye 2004[157] | 2002 | Nigeria | 🟢 | 🟢 | 🟢 | 🔴 | 🟡 | 🔴 | 🟢 | 🟢 | 🟢 | 🔴 | 6 | Moderate |
| Namkinga 2005[158] | 2002 | Tanzania | 🟢 | 🟢 | 🟢 | 🟡 | 🟢 | 🟢 | 🟢 | 🟢 | 🔴 | 🟢 | 8 | Lower |
| Mwansasu 2002[159] | 2002* | Tanzania | 🟢 | 🟢 | 🟢 | 🟢 | 🟢 | 🟢 | 🔴 | 🟢 | 🔴 | 🟢 | 8 | Lower |
| Rassjo 2006[160] | 2002 | Uganda | 🔴 | 🟢 | 🔴 | 🟢 | 🟡 | 🔴 | 🟢 | 🟢 | 🔴 | 🔴 | 4 | Higher |
| Fayemiwo 2011[161] | 2003 | Nigeria | 🔴 | 🟢 | 🟢 | 🔴 | 🟡 | 🟢 | 🔴 | 🟢 | 🔴 | 🔴 | 4 | Higher |
| Msuya 2009[162] | 2003 | Tanzania | 🔴 | 🟢 | 🔴 | 🟡 | 🟡 | 🟢 | 🟢 | 🟢 | 🟢 | 🔴 | 5 | Moderate |
| O'Farrell 2008[163] | 2004 | South Africa | 🟢 | 🟢 | 🟢 | 🟢 | 🟢 | 🟢 | 🟢 | 🟢 | 🟢 | 🟢 | 10 | Lower |
| Weiss 2011[164] | 2004 | Central African Republic | 🔴 | 🟢 | 🟢 | 🟢 | 🟢 | 🟢 | 🟢 | 🟢 | 🟢 | 🟢 | 9 | Lower |
| Legoff 2007[165] | 2004 | Central African Republic, Ghana | 🔴 | 🟢 | 🟢 | 🟢 | 🟢 | 🟢 | 🟢 | 🟢 | 🟢 | 🟢 | 9 | Lower |
| Abauleth 2006[166] | 2004 | Côte d'Ivoire | 🟢 | 🟢 | 🔴 | 🟡 | 🟡 | 🔴 | 🟢 | 🟢 | 🟢 | 🟢 | 6 | Moderate |
| Mehta 2012[167] | 2004 | Kenya | 🔴 | 🟢 | 🟢 | 🔴 | 🟢 | 🟢 | 🟢 | 🟢 | 🔴 | 🟢 | 7 | Moderate |
| Onyekonwu 2011[168] | 2004 | Nigeria | 🟢 | 🟢 | 🔴 | 🟢 | 🟢 | 🟢 | 🟢 | 🟢 | 🟢 | 🟢 | 9 | Lower |
| Suntoke 2009[169] | 2004 | Uganda | 🔴 | 🟢 | 🟢 | 🟢 | 🟢 | 🟢 | 🟢 | 🟢 | 🟢 | 🟢 | 9 | Lower |
| Pepin 2006[170] | 2005 | Ghana, Guinea, Mali, Togo | 🔴 | 🟢 | 🟢 | 🟡 | 🟢 | 🟢 | 🟢 | 🟢 | 🟢 | 🟢 | 8 | Lower |
| Pepin 2011[171] | 2005 | Ghana, Guinea, Mali, Togo | 🔴 | 🟢 | 🟢 | 🟡 | 🟢 | 🟢 | 🟢 | 🟢 | 🟢 | 🟢 | 8 | Lower |
| Phiri 2013[172] | 2005 | Malawi | 🟢 | 🟢 | 🟢 | 🟢 | 🟢 | 🟢 | 🟢 | 🟢 | 🟢 | 🟢 | 10 | Lower |
| Brankin 2009[173] | 2005 | Uganda | 🔴 | 🟢 | 🔴 | 🔴 | 🟡 | 🟢 | 🔴 | 🟢 | 🟢 | 🟢 | 5 | Moderate |
| Zimba 2011[174] | 2005 | Mozambique | 🟢 | 🟢 | 🟢 | 🟢 | 🟢 | 🟢 | 🔴 | 🟢 | 🟢 | 🟢 | 9 | Lower |
| Paz-Bailey 2010[175] | 2006 | South Africa | 🔴 | 🟢 | 🟢 | 🟡 | 🟢 | 🟢 | 🟢 | 🟢 | 🟢 | 🔴 | 7 | Moderate |
| Paz-Bailey 2009[176] | 2006 | South Africa | 🔴 | 🟢 | 🟢 | 🟡 | 🟢 | 🟢 | 🟢 | 🟢 | 🟢 | 🔴 | 7 | Moderate |
| Ojuromi 2007[177] | 2006 | Nigeria | 🟢 | 🟢 | 🔴 | 🟡 | 🟡 | 🔴 | 🟢 | 🟢 | 🟢 | 🟢 | 6 | Moderate |
| Nwadioha 2012[178] | 2007 | Nigeria | 🟢 | 🟢 | 🟢 | 🟡 | 🟡 | 🟡 | 🟢 | 🟢 | 🔴 | 🟢 | 6 | Moderate |
| Nwadioha 2011[179] | 2007 | Nigeria | 🟢 | 🟢 | 🟢 | 🟡 | 🟡 | 🟢 | 🟢 | 🟢 | 🔴 | 🟢 | 7 | Moderate |
| Bochner 2017[180] | 2007 | Botswana, Kenya, Rwanda, South Africa, Tanzania, Uganda | 🔴 | 🟢 | 🟢 | 🔴 | 🟢 | 🟢 | 🟢 | 🔴 | 🔴 | 🟢 | 6 | Moderate |
| Brown 2010[181] | 2007 | Malawi | 🔴 | 🟢 | 🔴 | 🟢 | 🟡 | 🟢 | 🟢 | 🟢 | 🔴 | 🟢 | 6 | Moderate |
| Nwadioha 2011[182] | 2007 | Nigeria | 🟢 | 🔴 | 🔴 | 🔴 | 🟡 | 🟡 | 🟢 | 🟢 | 🟢 | 🟢 | 5 | Moderate |
| Muvunyi 2009[183] | 2007 | Rwanda | 🟢 | 🟢 | 🔴 | 🟢 | 🔴 | 🔴 | 🔴 | 🟢 | 🔴 | 🔴 | 4 | Higher |

| Reference | Year[1] | Country | Q1 | Q2 | Q3 | Q4 | Q5 | Q6 | Q7 | Q8 | Q9 | Q10 | Score | Rating |
|---|---|---|---|---|---|---|---|---|---|---|---|---|---|---|
| Moodley 2013[184] | 2007 | South Africa | 🟢 | 🟢 | 🟢 | 🟡 | 🟢 | 🔴 | 🟢 | 🟢 | 🔴 | 🟢 | 7 | Moderate |
| LeRoux 2010[185] | 2008 | South Africa | 🟢 | 🟢 | 🔴 | 🟢 | 🟢 | 🟢 | 🔴 | 🟢 | 🟢 | 🟢 | 8 | Lower |
| Nwadioha 2010[186] | 2008 | Nigeria | 🟢 | 🟢 | 🟢 | 🟡 | 🟡 | 🟡 | 🟢 | 🟢 | 🟢 | 🔴 | 6 | Moderate |
| Nwadioha 2010[187] | 2008 | Nigeria | 🟢 | 🟢 | 🔴 | 🟡 | 🟡 | 🟡 | 🟢 | 🟢 | 🟢 | 🔴 | 5 | Moderate |
| Djohan 2012[188] | 2008 | Côte d'Ivoire | 🔴 | 🟢 | 🔴 | 🟢 | 🟡 | 🔴 | 🟢 | 🟢 | 🔴 | 🟢 | 5 | Moderate |
| Vandenhoudt 2013[189] | 2008 | Kenya | 🔴 | 🟢 | 🟢 | 🔴 | 🟡 | 🟢 | 🔴 | 🟢 | 🟢 | 🟢 | 6 | Moderate |
| Vandepitte 2012[190] | 2009 | Uganda | 🔴 | 🟢 | 🟢 | 🟢 | 🟢 | 🟢 | 🟢 | 🟢 | 🔴 | 🔴 | 7 | Moderate |
| Vandepitte 2011[191] | 2009 | Uganda | 🔴 | 🟢 | 🟢 | 🟢 | 🟡 | 🟢 | 🔴 | 🟢 | 🟢 | 🔴 | 6 | Moderate |
| Karou 2012[192] | 2009 | Burkina Faso | 🟢 | 🟢 | 🔴 | 🟡 | 🟡 | 🔴 | 🟢 | 🟢 | 🟢 | 🔴 | 5 | Moderate |
| Tine 2019[193] | 2009 | Senegal | 🟢 | 🟢 | 🔴 | 🟡 | 🟡 | 🟢 | 🟢 | 🔴 | 🔴 | 🟢 | 5 | Moderate |
| Nwadioha 2013[194] | 2009 | Nigeria | 🔴 | 🟢 | 🔴 | 🟡 | 🟡 | 🔴 | 🟢 | 🟢 | 🔴 | 🟢 | 4 | Higher |
| Okunlola 2009[195] | 2009 | Nigeria | 🔴 | 🟢 | 🔴 | 🟡 | 🟢 | 🟢 | 🔴 | 🟢 | 🟢 | 🟢 | 6 | Moderate |
| De Jongh 2009[196] | 2009* | South Africa | 🟢 | 🟢 | 🔴 | 🟢 | 🟢 | 🟡 | 🔴 | 🟢 | 🟢 | 🟢 | 7 | Moderate |
| Djomand 2016[197] | 2010 | Kenya | 🟢 | 🟢 | 🟢 | 🟢 | 🟡 | 🟢 | 🟢 | 🟢 | 🟢 | 🟢 | 9 | Lower |
| Singa 2013[198] | 2010 | Kenya | 🔴 | 🟢 | 🟢 | 🔴 | 🟢 | 🟢 | 🔴 | 🟢 | 🟢 | 🔴 | 6 | Moderate |
| De Jongh 2010[199] | 2010* | South Africa | 🔴 | 🟢 | 🔴 | 🟡 | 🟢 | 🟢 | 🟢 | 🟢 | 🟢 | 🟢 | 7 | Moderate |
| Makasa 2012[200] | 2010 | Zambia | 🟢 | 🟢 | 🟢 | 🟢 | 🟢 | 🟢 | 🟢 | 🟢 | 🟢 | 🟢 | 10 | Lower |
| Ibrahim 2014[201] | 2011 | Nigeria | 🟢 | 🟢 | 🔴 | 🟢 | 🟢 | 🔴 | 🟢 | 🟢 | 🔴 | 🔴 | 6 | Moderate |
| Ibrahim 2013[202] | 2011 | Nigeria | 🟢 | 🟢 | 🔴 | 🟢 | 🟢 | 🔴 | 🟢 | 🟢 | 🔴 | 🔴 | 6 | Moderate |
| Konate 2014[203] | 2011 | Côte d'Ivoire | 🟢 | 🟢 | 🔴 | 🟡 | 🟡 | 🔴 | 🟢 | 🟢 | 🔴 | 🔴 | 4 | Higher |
| Takuva 2014[204] | 2011 | Zimbabwe | 🟢 | 🟢 | 🔴 | 🟡 | 🟢 | 🟢 | 🟢 | 🟢 | 🟢 | 🔴 | 7 | Moderate |
| Van der Eem 2016[205] | 2012 | South Africa | 🟢 | 🟢 | 🟢 | 🟢 | 🟡 | 🟢 | 🔴 | 🟢 | 🟢 | 🟢 | 8 | Lower |
| Mulu 2017[206] | 2013 | Ethiopia | 🔴 | 🔴 | 🔴 | 🟢 | 🟡 | 🔴 | 🔴 | 🔴 | 🔴 | 🔴 | 1 | Higher |
| Kerubo 2016[207] | 2013 | Kenya | 🔴 | 🟢 | 🔴 | 🟢 | 🟡 | 🔴 | 🔴 | 🟢 | 🔴 | 🟢 | 4 | Higher |
| Tine 2019[208] | 2013 | Senegal | 🟢 | 🟢 | 🔴 | 🟡 | 🟡 | 🟢 | 🟢 | 🔴 | 🔴 | 🟢 | 5 | Moderate |
| NICD 2023[209] | 2014 | South Africa | 🟢 | 🟢 | 🟢 | 🟢 | 🟢 | 🟢 | 🟢 | 🟢 | 🟢 | 🟢 | 10 | Lower |
| Nkwabong 2015[210] | 2014 | Cameroon | 🔴 | 🟢 | 🔴 | 🟡 | 🟡 | 🟢 | 🔴 | 🟢 | 🟢 | 🟢 | 5 | Moderate |
| Fentaw 2020[211] | 2014 | Ethiopia | 🔴 | 🟢 | 🔴 | 🟡 | 🟡 | 🟢 | 🟢 | 🟢 | 🔴 | 🟢 | 5 | Moderate |
| Dela 2019[212] | 2014 | Ghana | 🔴 | 🟢 | 🔴 | 🟡 | 🟡 | 🔴 | 🟢 | 🟢 | 🔴 | 🟢 | 4 | Higher |
| Asmah 2018[213] | 2014 | Ghana | 🔴 | 🟢 | 🔴 | 🔴 | 🟡 | 🔴 | 🔴 | 🔴 | 🔴 | 🟢 | 2 | Higher |
| Rietmeijer 2019[214] | 2015 | Zimbabwe | 🟢 | 🟢 | 🟢 | 🟢 | 🟢 | 🟢 | 🟢 | 🟢 | 🟢 | 🟢 | 10 | Lower |
| Chirenje 2018[215] | 2015 | Zimbabwe | 🟢 | 🟢 | 🟢 | 🟢 | 🟢 | 🟢 | 🟢 | 🟢 | 🟢 | 🟢 | 10 | Lower |
| Kilmarx 2018[216] | 2015 | Zimbabwe | 🟢 | 🟢 | 🟢 | 🟢 | 🟢 | 🟢 | 🟢 | 🟢 | 🟢 | 🔴 | 9 | Lower |
| Mungati 2018[217] | 2015 | Zimbabwe | 🟢 | 🟢 | 🟢 | 🟢 | 🟢 | 🟢 | 🟢 | 🟢 | 🟢 | 🟢 | 10 | Lower |
| Rietmeijer 2018[218] | 2015 | Zimbabwe | 🟢 | 🟢 | 🟢 | 🟢 | 🟢 | 🟢 | 🟢 | 🟢 | 🟢 | 🟢 | 10 | Lower |
| Konadu 2019[219] | 2015 | Ghana | 🔴 | 🟢 | 🔴 | 🔴 | 🟢 | 🟢 | 🟢 | 🟢 | 🔴 | 🔴 | 5 | Moderate |
| Odunvbun 2018[220] | 2015 | Nigeria | 🔴 | 🟢 | 🔴 | 🟡 | 🟡 | 🔴 | 🔴 | 🟢 | 🔴 | 🔴 | 2 | Higher |

| Reference | Year[1] | Country | Q1 | Q2 | Q3 | Q4 | Q5 | Q6 | Q7 | Q8 | Q9 | Q10 | Score | Rating |
|---|---|---|---|---|---|---|---|---|---|---|---|---|---|---|
| Oyeyipo 2015[221] | 2015 | Nigeria | 🔴 | 🟢 | 🔴 | 🟡 | 🟢 | 🔴 | 🟢 | 🟢 | 🔴 | 🟢 | 5 | Moderate |
| Marcotte 2019[222] | 2015 | South Africa | 🔴 | 🟢 | 🟢 | 🟡 | 🟡 | 🔴 | 🔴 | 🟢 | 🔴 | 🔴 | 3 | Higher |
| Nabweyambo 2017[223] | 2015 | Uganda | 🔴 | 🟢 | 🔴 | 🟢 | 🟡 | 🔴 | 🟢 | 🟢 | 🔴 | 🟢 | 5 | Moderate |
| Sakala 2016[224] | 2015 | Zambia | 🔴 | 🔴 | 🟢 | 🟢 | 🟡 | 🟢 | 🔴 | 🟢 | 🔴 | 🔴 | 4 | Higher |
| Van Liere 2019[225] | 2016 | South Africa | 🔴 | 🟢 | 🟢 | 🔴 | 🟡 | 🔴 | 🔴 | 🟢 | 🟢 | 🟢 | 5 | Moderate |
| Latif 2018[226] | 2016 | Zimbabwe | 🔴 | 🟢 | 🔴 | 🟡 | 🟢 | 🟢 | 🟢 | 🟢 | 🔴 | 🟢 | 6 | Moderate |
| Mehta 2017[227] | 2017* | Kenya | 🟢 | 🟢 | 🟢 | 🟢 | 🟡 | 🟢 | 🔴 | 🟢 | 🟢 | 🔴 | 7 | Moderate |
| Gadoth 2020[228] | 2017 | Democratic Republic of Congo | 🔴 | 🟢 | 🟢 | 🔴 | 🟡 | 🔴 | 🔴 | 🟢 | 🔴 | 🔴 | 3 | Higher |
| Chen 2020[229] | 2017 | Malawi | 🔴 | 🟢 | 🟢 | 🟢 | 🟡 | 🟢 | 🔴 | 🟢 | 🔴 | 🟢 | 6 | Moderate |
| Aduloju 2019[230] | 2017 | Nigeria | 🔴 | 🟢 | 🔴 | 🟢 | 🟡 | 🟢 | 🟢 | 🔴 | 🔴 | 🟢 | 5 | Moderate |
| Kashibu 2018[231] | 2017 | Nigeria | 🔴 | 🟢 | 🔴 | 🟢 | 🟡 | 🟢 | 🔴 | 🟢 | 🔴 | 🟢 | 5 | Moderate |
| Verwijs 2019[232] | 2017 | Rwanda | 🔴 | 🟢 | 🟢 | 🔴 | 🟡 | 🟢 | 🟢 | 🟢 | 🟢 | 🟢 | 7 | Moderate |
| Chabata 2023[233] | 2017 | Zimbabwe | 🔴 | 🟢 | 🟢 | 🔴 | 🟢 | 🟢 | 🔴 | 🟢 | 🟢 | 🟢 | 7 | Moderate |
| Nacht 2020[234] | 2018 | Kenya | 🔴 | 🟢 | 🔴 | 🔴 | 🔴 | 🟢 | 🔴 | 🟢 | 🔴 | 🔴 | 3 | Higher |
| Wall 2021[235] | 2018 | Rwanda | 🟢 | 🟢 | 🟢 | 🔴 | 🟡 | 🟢 | 🟢 | 🟢 | 🔴 | 🟢 | 7 | Moderate |
| Happel 2020[236] | 2018 | South Africa | 🔴 | 🟢 | 🟢 | 🟢 | 🟢 | 🔴 | 🔴 | 🟢 | 🟢 | 🟢 | 7 | Moderate |
| Connolly 2020[237] | 2018 | Zambia | 🔴 | 🟢 | 🟢 | 🔴 | 🟡 | 🟢 | 🟢 | 🟢 | 🔴 | 🔴 | 5 | Moderate |
| Mabaso 2020[238] | 2019 | South Africa | 🔴 | 🟢 | 🔴 | 🟡 | 🟡 | 🔴 | 🔴 | 🟢 | 🔴 | 🟢 | 3 | Higher |
| Melendez 2022[239] | 2020 | Uganda | 🟢 | 🟢 | 🟢 | 🟡 | 🟢 | 🔴 | 🟢 | 🟢 | 🟢 | 🔴 | 7 | Moderate |
| Zenebe 2021[240] | 2020 | Ethiopia | 🔴 | 🟢 | 🔴 | 🟢 | 🟡 | 🔴 | 🔴 | 🟢 | 🟢 | 🟢 | 5 | Moderate |
| Ahmed 2022[241] | 2021 | Ethiopia | 🔴 | 🟢 | 🔴 | 🟡 | 🟡 | 🟢 | 🔴 | 🟢 | 🔴 | 🔴 | 3 | Higher |

**Q1:** Was the study specifically designed to estimate the outcome/s of interest? **Q2:** Were the criteria for inclusion in the sample clearly defined? **Q3:** Were study participants and their setting described in sufficient detail to determine representativeness of the target population? **Q4:** Were study participants recruited in an appropriate way? **Q5:** Was the participation rate adequate to minimise selection bias? **Q6:** Were objective, standard criteria used to define participant symptoms? **Q7:** Was the sample size sufficient to measure the outcome? **Q8**: Were the outcomes measured in a reliable and consistent way? **Q9:** Were participants tested for multiple pathogens to prevent misclassification of infection outcomes? **Q10:** Was the reporting of the outcome(s) unambiguous?

[1]Midpoint year of data collection. *Publication year used as a proxy for studies that did not report dates or year(s) of data collection.

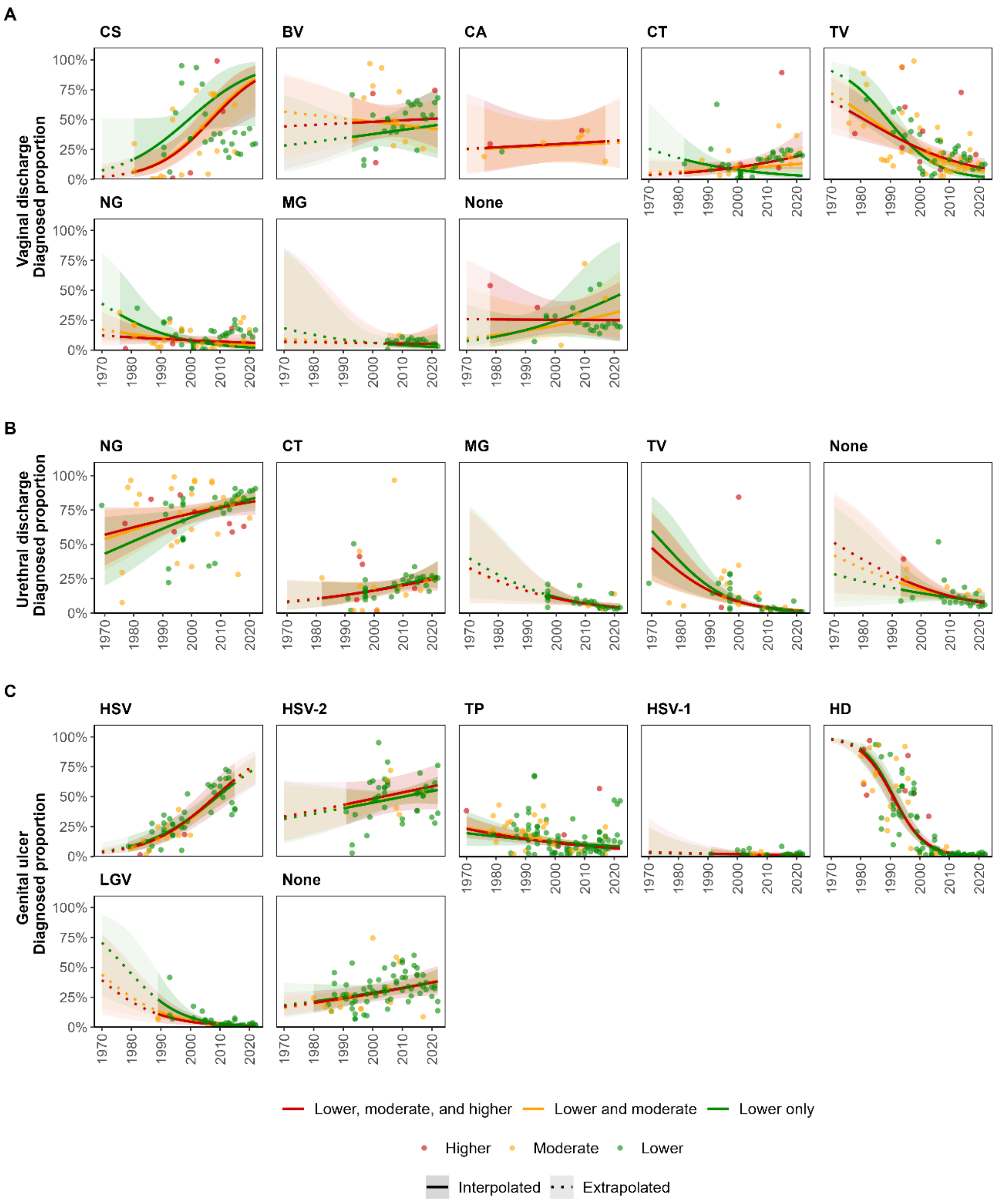

**Figure S3: Estimated diagnosed proportion per pathogen among symptomatic adults of mixed or unmeasured HIV-status in sub-Saharan Africa over time, according to risk of bias.**

Diagnosed proportion per pathogen among individuals symptomatic with (A) vaginal discharge, (B) urethral discharge, and (C) genital ulcer. Proportions estimated using generalised linear mixed-effects models for each symptom, by including studies with all levels of bias, moderate and lower risk of bias, and lower risk of bias only. Lines and shaded areas represent sex-matched population-weighted mean proportions and 95% confidence intervals. Solid lines and darker shading denote estimates and confidence intervals within the observed data range (interpolated), while dotted lines and lighter shading indicate estimates and confidence intervals beyond that time frame (extrapolated). Points represent study observations adjusted for diagnostic test performance and colour coded according to risk of bias.

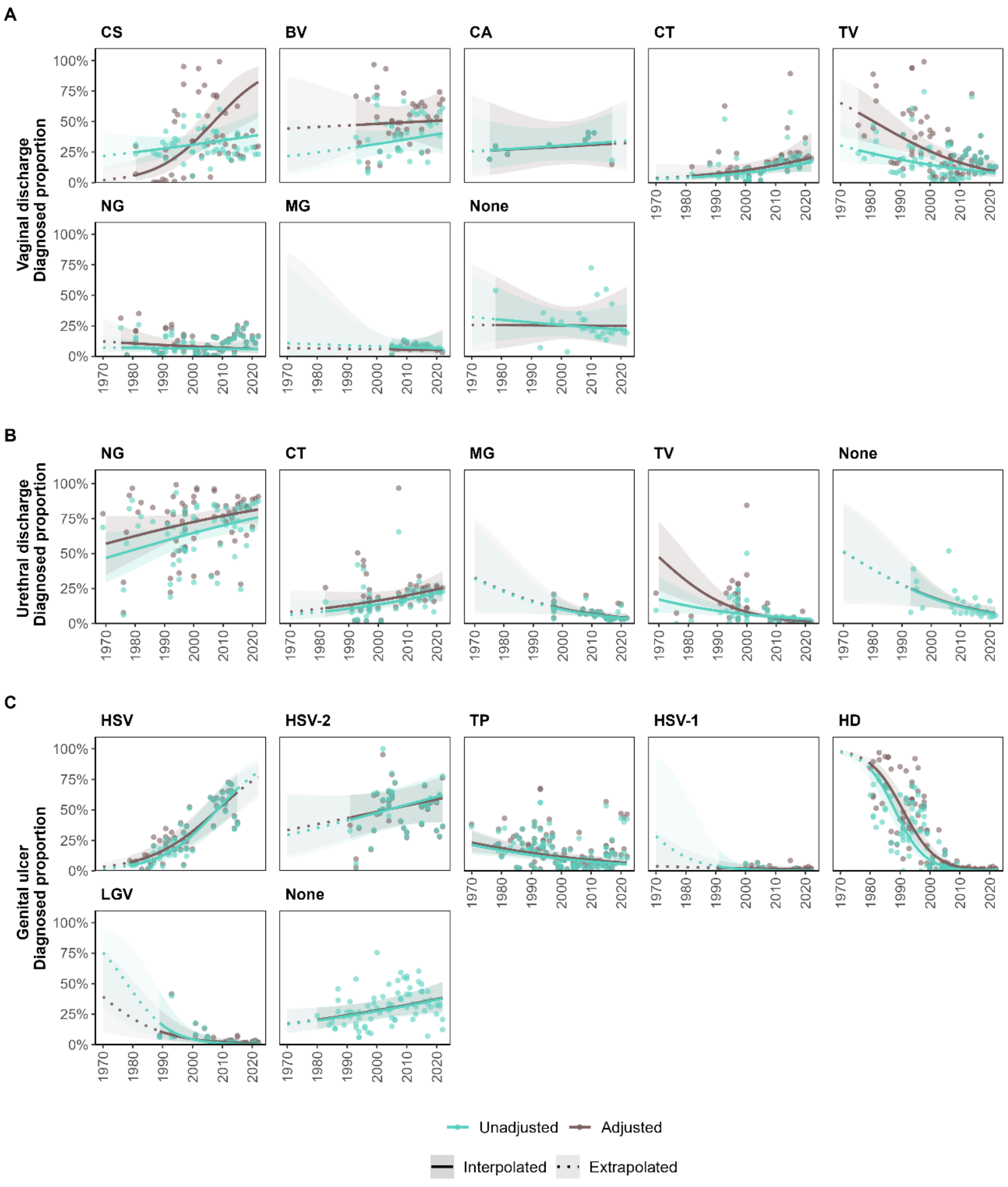

**Figure S4: Estimated diagnosed proportion per pathogen among symptomatic adults of mixed or unmeasured HIV-status in sub-Saharan Africa over time, with and without diagnostic test performance adjustment.**

Diagnosed proportion per pathogen among individuals symptomatic with (A) vaginal discharge, (B) urethral discharge, and (C) genital ulcer. Proportions estimated using generalised linear mixed-effects models for each symptom, using either observations as reported or observations adjusted for diagnostic test performance. Lines and shaded areas represent sex-matched population-weighted mean proportions and 95% confidence intervals. Solid lines and darker shading denote estimates and confidence intervals within the observed data range (interpolated), while dotted lines and lighter shading indicate estimates and confidence intervals beyond that time frame (extrapolated). Points represent study observations that are unadjusted or adjusted for diagnostic test performance.

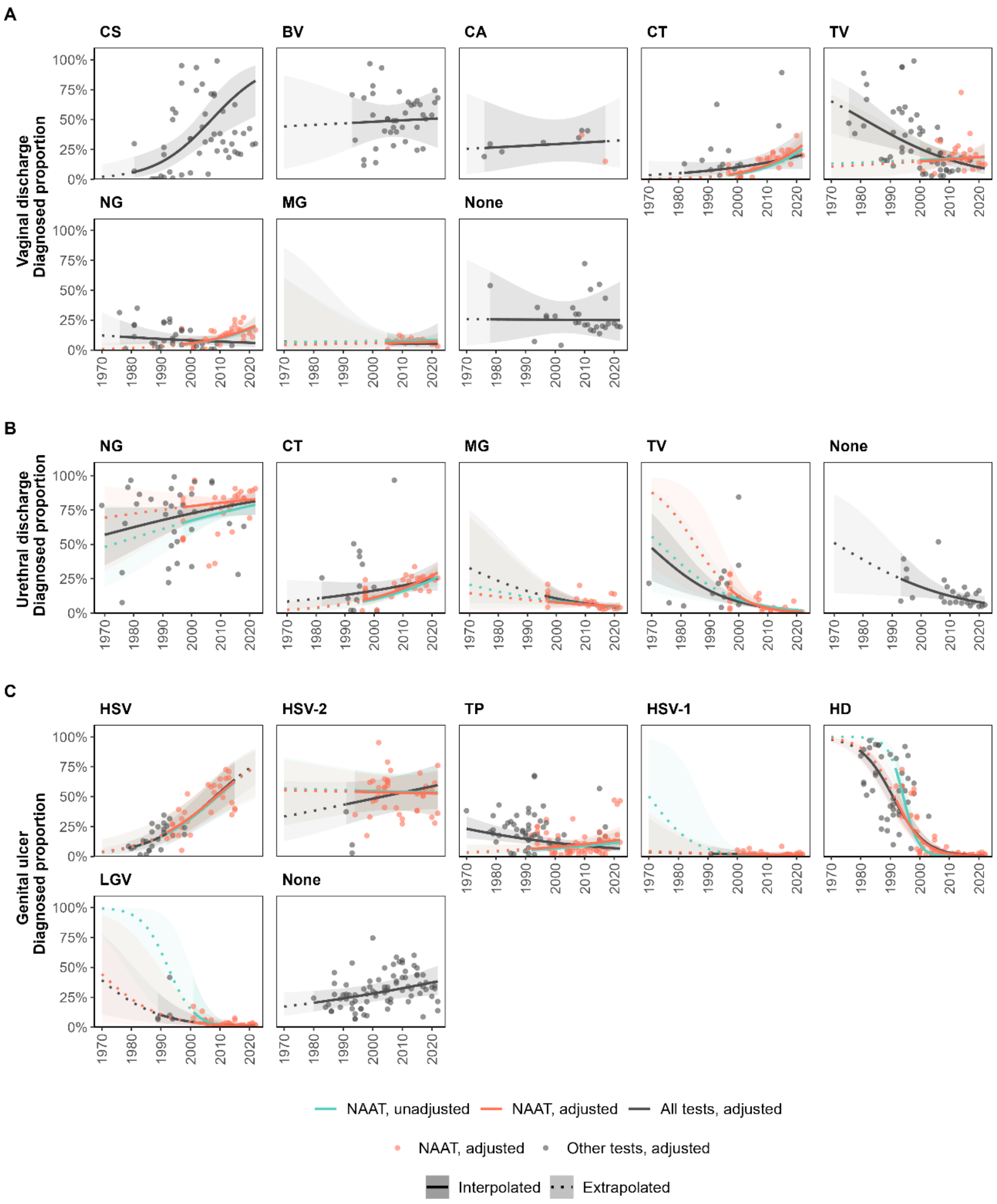

**Figure S5: Estimated diagnosed proportion per pathogen among symptomatic adults of mixed or unmeasured HIV-status in sub-Saharan Africa over time, using NAAT with and without diagnostic test performance adjustment.**

Diagnosed proportion per pathogen among individuals symptomatic with (A) vaginal discharge, (B) urethral discharge, and (C) genital ulcer. Proportions estimated using generalised linear mixed-effects models for each symptom, using observations attained using nucleic acid amplification testing (NAAT) as reported or adjusted for diagnostic test performance, compared to observations from all diagnostics adjusted for diagnostic test performance. Lines and shaded areas represent sex-matched population-weighted mean proportions and 95% confidence intervals. Solid lines and darker shading denote estimates and confidence intervals within the observed data range (interpolated), while dotted lines and lighter shading indicate estimates and confidence intervals beyond that time frame (extrapolated). Points represent study observations adjusted for diagnostic test performance and colour coded according to diagnostic test.

### References

1 UN Statistics Division. Standard country or area codes for statistical use (M49). https://unstats.un.org/unsd/methodology/m49/ (accessed June 14, 2023).

2 Kufa T, Gumede L, Maseko D V, Radebe F, Kularatne R. The demographic and clinical profiles of women presenting with vaginal discharge syndrome at primary care facilities in South Africa: Associations with age and implications for management. *S Afr Med J* 2018; **108**: 876–80.

3 Kufa T, Radebe F, Cutler E, *et al.* Recency of HIV infection, antiretroviral therapy use and viral loads among symptomatic sexually transmitted infection service attendees in South Africa. *South African Med J* 2022; **112**: 96–101.

4 Kularatne RS, Muller EE, Maseko D V, Kufa-Chakezha T, Lewis DA. Trends in the relative prevalence of genital ulcer disease pathogens and association with HIV infection in Johannesburg, South Africa, 2007-2015. *PLoS One* 2018; **13**: e0194125.

5 Kularatne R, Maseko V, Mahlangu P, Muller E, Kufa T. Etiological Surveillance of Male Urethritis Syndrome in South Africa: 2019 to 2020. *Sex Transm Dis* 2022; **49**: 560–4.

6 Kularatne R, Muller E, Maseko V, Dias BDC, Kufa T. Etiological Surveillance of Vaginal Discharge Syndrome in South Africa: 2019 to 2020. *Sex Transm Dis* 2022; **49**: 565–70.

7 Kularatne R, Venter JME, Maseko V, Muller E, Kufa T. Etiological Surveillance of Genital Ulcer Syndrome in South Africa: 2019 to 2020. *Sex Transm Dis* 2022; **49**: 571–5.

8 Mhlongo S, Magooa P, Müller EE, *et al.* Etiology and STI/HIV coinfections among patients with urethral and vaginal discharge syndromes in South Africa. *Sex Transm Dis* 2010; **37**: 566–70.

9 Mathebula RC, Kuonza LR, Musekiwa A, Kularatne R, Maseko V, Kufa T. Factors associated with repeat genital symptoms among sexually transmitted infection service attendees in South Africa, 2015 - 2016. *S Afr Med J* 2020; **110**: 661–6.

10 Lewis DA, Muller E, Steele L, *et al.* Prevalence and associations of genital ulcer and urethral pathogens in men presenting with genital ulcer syndrome to primary health care clinics in South Africa. *Sex Transm Dis* 2012; **39**: 880–5.

11 Lewis DA, Marsh K, Radebe F, Maseko V, Hughes G. Trends and associations of Trichomonas vaginalis infection in men and women with genital discharge syndromes in Johannesburg, South Africa. *Sex Transm Infect* 2013; **89**: 523–7.

12 Lewis DA, Scott L, Slabbert M, *et al.* Escalation in the relative prevalence of ciprofloxacin-resistant gonorrhoea among men with urethral discharge in two South African cities: association with HIV seropositivity. *Sex Transm Infect* 2008; **84**: 352–5.

13 Fearon E, Chabata ST, Magutshwa S, *et al.* Estimating the Population Size of Female Sex Workers in Zimbabwe: Comparison of Estimates Obtained Using Different Methods in Twenty Sites and Development of a National-Level Estimate. 2020. www.jaids.com (accessed Oct 1, 2023).

14 Stannah J, Soni N, Keng J, *et al.* Trends in HIV testing, the treatment cascade, and HIV incidence among men who have sex with men in Africa: a systematic review and meta-analysis. *Lancet HIV* 2023; **10**: E528–42.

15 World Health Organization. Laboratory tests for the detection of reproductive tract infections. 1999.

16 World Health Organization. Prevalence and incidence of selected sexually transmitted infections: Methods and results used by WHO to generate 2005 estimates. 2011 http://www.who.int/about/licensing/ (accessed Feb 18, 2022).

17 Kaydos-Daniels SC, Miller WC, Hoffman I, *et al.* The Use of Specimens from Various Genitourinary Sites in Men, to Detect Trichomonas vaginalis Infection. *J Infect Dis* 2004; **189**: 1926–31.

18 Hobbs MM, Lapple DM, Lawing LF, *et al.* Methods for detection of Trichomonas vaginalis in the male partners of infected women: implications for control of trichomoniasis. *J Clin Microbiol* 2006; **44**: 3994–9.

19 Rogan WJ, Gladen B. Estimating prevalence from the results of a screening test. *Am J Epidemiol* 1978; **107**: 71–6.

20 Flor M, Weiß M, Selhorst T, Müller-Graf C, Greiner M. Comparison of Bayesian and frequentist methods for prevalence estimation under misclassification. *BMC Public Health* 2020; **20**: 1135.

21 Speybroeck N, Devleesschauwer B, Joseph L, Berkvens D. Misclassification errors in prevalence estimation: Bayesian handling with care. *Int J Public Health* 2013; **58**: 791–5.

22 Dadhwal V, Hariprasad R, Mittal S, Kapil A. Prevalence of bacterial vaginosis in pregnant women and predictive value of clinical diagnosis. *Arch Gynecol Obstet* 2010; **281**: 101–4.

23 Thomason JL, Anderson RJ, Gelbart SM, *et al.* Simplified Gram stain interpretive method for diagnosis of bacterial vaginosis. *Am J Obstet Gynecol* 1992; **167**: 16–9.

24 Schwebke JR, Hillier SL, Sobel JD, McGregor JA, Sweet RL. Validity of the vaginal gram stain for the diagnosis of bacterial vaginosis. *Obstet Gynecol* 1996; **88**: 573–6.

25 Hoppe JE, Frey P. Evaluation of six commercial tests and the germ-tube test for presumptive identification of Candida albicans. *Eur J Clin Microbiol Infect Dis* 1999; **18**: 188–91.

26 Gaydos CA, Beqaj S, Schwebke JR, *et al.* Clinical Validation of a Test for the Diagnosis of Vaginitis. *Obstet Gynecol* 2017; **130**: 181–9.

27 Chatwani AJ, Mehta R, Hassan S, Rahimi S, Jeronis S, Dandolu V. Rapid testing for vaginal yeast

detection: a prospective study. *Obs Gynecol* 2007; **196**: 309–10.
28 Schwebke JR, Gaydos CA, Nyirjesy P, Paradis S, Kodsi S, Cooper CK. Diagnostic Performance of a Molecular Test versus Clinician Assessment of Vaginitis. 2018. https://doi.org/10 (accessed May 30, 2023).
29 Grillo-Ardila CF, Torres M, Gaitán HG. Rapid point of care test for detecting urogenital Chlamydia trachomatis infection in nonpregnant women and men at reproductive age. *Cochrane database Syst Rev* 2020; **1**. DOI:10.1002/14651858.CD011708.PUB2.
30 Gaydos CA, Manhart LE, Taylor SN, *et al.* Molecular testing for mycoplasma genitalium in the United States: Results from the AMES prospective multicenter clinical study. *J Clin Microbiol* 2019; **57**. DOI:10.1128/JCM.01125-19.
31 Paz-Bailey G, Rahman M, Chen C, *et al.* Changes in the etiology of sexually transmitted diseases in Botswana between 1993 and 2002: implications for the clinical management of genital ulcer disease. *Clin Infect Dis* 2005; **41**: 1304–12.
32 Legoff J, Péré H, Bélec L. Diagnosis of genital herpes simplex virus infection in the clinical laboratory. *Virol J* 2014; **11**. DOI:10.1186/1743-422X-11-83.
33 Arshad Z, Alturkistani A, Brindley D, Lam C, Foley K, Meinert E. Tools for the diagnosis of herpes simplex virus 1/2: Systematic review of studies published between 2012 and 2018. *JMIR Public Heal Surveill* 2019; **5**. DOI:10.2196/14216.
34 Diagnostic Hybrids. D3 DFA Chlamydiae Culture Confirmation Kit. REF: 01-040000. Athens, Ohio, 2008.
35 Touati A, Laurier-Nadalié C, Bébéar C, Peuchant O, de Barbeyrac B. Evaluation of four commercial real-time PCR assays for the detection of lymphogranuloma venereum in Chlamydia trachomatis-positive anorectal samples. *Clin Microbiol Infect* 2021; **27**: 909.e1-909.e5.
36 Peeling RW, Ye H. Diagnostic tools for preventing and managing maternal and congenital syphilis: An overview. *Bull World Health Organ* 2004; **82**: 439–46.
37 Hambie EA, Larsen SA, Perryman MW, Pettit DE, Mullally RL, Whittington W. Comparison of a new rapid plasma reagin card test with the standard rapid plasma reagin 18-mm circle card test and the venereal disease research laboratory slide test for serodiagnosis of syphilis. *J Clin Microbiol* 1983; **17**: 249–54.
38 Perryman MW, Larsen SA, Hambie EA, Pettit DE, Mullally RL, Whittington W. Evaluation of a new rapid plasma reagin card test as a screening test for syphilis. *J Clin Microbiol* 1982; **16**: 286–90.
39 Parham CE, Pettit DE, Larsen SA, Hambie EA, Perryman MW, McGrew BE. Interlaboratory comparison of the toluidine red unheated serum test antigen preparation. *J Clin Microbiol* 1984; **20**: 434–7.
40 Cantor AG, Pappas M, Daeges M, Nelson HD. Screening for Syphilis: Updated Evidence Report and Systematic Review for the US Preventive Services Task Force. *JAMA* 2016; **315**: 2328–37.
41 Sena AC, White BL, Sparling PF. Novel Treponema pallidum Serologic Tests: A Paradigm Shift in Syphilis Screening for the 21st Century. *Clin Infect Dis* 2010; **51**: 700–8.
42 Park IU, Fakile YF, Chow JM, *et al.* Performance of Treponemal Tests for the Diagnosis of Syphilis. *Clin Infect Dis* 2019; **68**: 913–8.
43 Munn Z, Moola S, Lisy K, Riitano D, Tufanaru C. Chapter 5: Systematic reviews of prevalence and incidence. In: Aromataris E, Munn Z, eds. JBI Manual for Evidence Synthesis. JBI, 2020.
44 Page MJ, McKenzie JE, Bossuyt PM, *et al.* The PRISMA 2020 statement: an updated guideline for reporting systematic reviews. *BMJ* 2021; **372**. DOI:10.1136/BMJ.N71.
45 Arya OP. Some highlights on the aetiology of the non gonococcal urethral discharge in males in Kampala, Uganda. *East Afr Med J* 1972; **49**: 817–24.
46 Ongom VL, Lwanga VN, Mugisha JK, Mafigiri JT. Social background to venereal disease at Kasangati. *East Afr Med J* 1971; **48**: 367–71.
47 Van Binsbergen JJ, Schulpen TWJ, Patel SC, Van Binsbergen-Ingelse A. Urethral or vaginal discharge at a rural hospital in Kenya. *East Afr Med J* 1980; **57**: 540–4.
48 Fakunle YM, Watkins B. Influence of self-medication on prevalence and antibiotic sensitivity of N. gonorrhoeae in Zaria (Nigeria). *East Afr Med J* 1976; **53**: 693–6.
49 Ballard RC, Schoub BD, Schneider J, Robins-Browne RM, Koornhof HJ. Urethritis in white men - a microbiological appraisal. *South African Med J* 1977; **51**: 702–6.
50 Rubin A, Russell JM, Mauff A. Efficacy of econazole in the treatment of candidiasis and other vaginal discharges. *South African Med J* 1980; **57**: 407–8.
51 Meheus A, Van Dyck E, Friedman F. Genital infections in Swaziland. *Ann Soc Belg Med Trop (1920)* 1982; **62**: 361–7.
52 Meheus A, Van Dyck E, Ursi JP, Ballard RC, Piot P. Etiology of genital ulcerations in Swaziland. *Sex Transm Dis* 1983; **10**: 33–5.
53 Ursi JP, van Dyck E, van Houtte C, *et al.* Syphilis in Swaziland: a serological survey. *Br J Vener Dis* 1981; **57**: 95–9.
54 Meheus A, Ballard R, Dlamini M, Ursi JP, Van Dyck E, Piot P. Epidemiology and aetiology of urethritis in Swaziland. *Int J Epidemiol* 1980; **9**: 239–45.
55 Nsanze H, Fast M V, D’Costa LJ, Tukei P, Curran J, Ronald A. Genital ulcers in Kenya. Clinical and laboratory study. *Br J Vener Dis* 1981; **57**: 378–81.

56 Fast M V, Nsanze H, Plummer FA, D'Costa LJ, MacLean IW, Ronald AR. Treatment of chancroid. A comparison of sulphamethoxazole and trimethoprim-sulphamethoxazole. *Br J Vener Dis* 1983; **59**: 320–4.
57 Yvert F, Riou JY, Frost E, Ivanoff B, Ossari S, Bouatsia P. [Gonococcal infections in Gabon (Haut-Ogooue)]. *Pathol Biol (Paris)* 1984; **32**: 80–4.
58 Hoosen AA, Ross SM, Mulla MJ, Patel M. The incidence of selected vaginal infections among pregnant urban Blacks. *South African Med J* 1981; **59**: 827–9.
59 Crewe-Brown HH. Genital ulceration in males at Ga-Rankuwa Hospital, Pretoria. *South African Med J* 1982; **62**.
60 D'Costa LJ, Plummer FA, Bowmer I, *et al.* Prostitutes are a major reservoir of sexually transmitted diseases in Nairobi, Kenya. *Sex Transm Dis* 1985; **12**: 64–7.
61 Mirza NB, Nsanze H, D'Costa LJ, Piot P. Microbiology of vaginal discharge in Nairobi, Kenya. *Br J Vener Dis* 1983; **59**: 186–8.
62 Mabey DC, Whittle HC. Genital and neonatal chlamydial infection in a trachoma endemic area. *Lancet* 1982; **2**: 300–1.
63 Mabey DC, Lloyd-Evans NE, Conteh S, Forsey T. Sexually transmitted diseases among randomly selected attenders at an antenatal clinic in The Gambia. *Br J Vener Dis* 1984; **60**: 331–6.
64 Plummer FA, Nsanze H, D'Costa LJ, *et al.* Single-dose therapy of chancroid with trimethoprim-sulfametrole. *N Engl J Med* 1983; **309**: 67–71.
65 Mauff AC, Ballard RC, Bilgeri YR, Koornhof HJ. Isolation of Haemophilus ducreyi from genital ulcerations in white men in Johannesburg. *South African Med J* 1983; **63**: 236–7.
66 Coovadia YM, Kharsany A, Hoosen A. The microbial aetiology of genital ulcers in black men in Durban, South Africa. *Genitourin Med* 1985; **61**: 266–9.
67 Greenblatt RM, Lukehart SA, Plummer FA, *et al.* Genital ulceration as a risk factor for human immunodeficiency virus infection. *AIDS* 1988; **2**: 47–50.
68 Plummer FA, D'Costa LJ, Nsanze H, *et al.* Clinical and microbiologic studies of genital ulcers in Kenyan women. *Sex Transm Dis* 1985; **12**: 193–7.
69 Kreiss JK, Koech D, Plummer FA, *et al.* AIDS virus infection in Nairobi prostitutes: spread of the epidemic to East Africa. *N Engl J Med* 1986; **314**: 414–8.
70 Dangor Y, Fehler G, Exposto FD, Koornhof HJ. Causes and treatment of sexually acquired genital ulceration in southern Africa. *South African Med J* 1989; **76**: 339–41.
71 Ismail SO, Ahmed HJ, Grillner L, Hederstedt B, Issa A, Bygdeman SM. Sexually transmitted diseases in men in Mogadishu, Somalia. *Int J STD AIDS* 1990; **1**: 102–6.
72 Meiring JA, Kemp E, Jennings DL, Mdhlovu M, Koornhof HJ. High-level penicillin-resistant gonococcal infections in Port Elizabeth. *South African Med J* 1989; **75**: 118–9.
73 Mabey DCW, Wall RA, Bello CSS. Aetiology of genital ulceration in the Gambia. *Genitourin Med* 1987; **63**: 312–5.
74 Leclerc A, Frost E, Collet M, Goeman J, Bedjabaga L. Urogenital Chlamydia trachomatis in Gabon: an unrecognised epidemic. *Genitourin Med* 1988; **64**: 308–11.
75 Bogaerts J, Kestens L, van Dyck E, Tello WM, Akingeneye J, Mukantabana V. Genital ulcers in a primary health clinic in Rwanda: impact of HIV infection on diagnosis and ulcer healing (1986-1992). *Int J STD AIDS* 1998; **9**: 706–10.
76 Faye-Kette YH, Kouassi AA, Sylla-Koko DF, *et al.* [Prevalence of 4 agents of sexually transmitted diseases in leukorrhea in Abidjan (Ivory Coast)]. *Bull la Société Pathol Exot* 1993; **86**: 245–7.
77 O'Farrell N, Hoosen AA, Coetzee KD, van den Ende J. Genital ulcer disease: accuracy of clinical diagnosis and strategies to improve control in Durban, South Africa. *Genitourin Med* 1994; **70**: 7–11.
78 Daly CC, Maggwa N, Mati JK, *et al.* Risk factors for gonorrhoea, syphilis, and trichomonas infections among women attending family planning clinics in Nairobi, Kenya. *Genitourin Med* 1994; **70**: 155–61.
79 Thior I, Diouf G, Diaw IK, *et al.* Sexually transmitted diseases and risk of HIV infection in men attending a sexually transmitted diseases clinic in Dakar, Senegal. *Afr J Reprod Health* 1997; **1**: 26–35.
80 Ndinya-Achola JO, Kihara AN, Fisher LD, *et al.* Presumptive specific clinical diagnosis of genital ulcer disease (GUD) in a primary health care setting in Nairobi. *Int J STD AIDS* 1996; **7**: 201–5.
81 Kamya MR, Nsubuga P, Grant RM, Hellman N. The high prevalence of genital herpes among patients with genital ulcer disease in Uganda. *Sex Transm Dis* 1995; **22**: 351–4.
82 Hanson S, Sunkutu RM, Kamanga J, Hojer B, Sandstrom E. STD care in Zambia: an evaluation of the guidelines for case management through a syndromic approach. *Int J STD AIDS* 1996; **7**: 324–32.
83 Le Bacq F, Mason PR, Gwanzura L, Robertson VJ, Latif AS. HIV and other sexually transmitted diseases at a rural hospital in Zimbabwe. *Genitourin Med* 1993; **69**: 352–6.
84 Kimani J, Bwayo JJ, Anzala AO, *et al.* Low dose erythromycin regimen for the treatment of chancroid. *East Afr Med J* 1995; **72**: 645–8.
85 Oni AA, Adu FD, Ekweozor CC, Bakare RA. Herpetic urethritis in male patients in Ibadan. *West Afr J Med* 1997; **16**: 27–9.
86 Totten PA, Kuypers JM, Chen CY, *et al.* Etiology of genital ulcer disease in Dakar, Senegal, and comparison of PCR and serologic assays for detection of Haemophilus ducreyi. *J Clin Microbiol* 2000; **38**:

268–73.
87 Htun Y, Radebe F, Fehler HG, Ballard RC. Changes in the patterns of sexually transmitted infection among South African mineworkers, associated with the emergence of the HIV/AIDS epidemic. *South African Med J* 2007; **97**: 1155–60.
88 Grosskurth H, Mayaud P, Mosha F, *et al.* Asymptomatic gonorrhoea and chlamydial infection in rural Tanzanian men. *Br Med J* 1996; **312**: 277–80.
89 Tswana SA, Nystrom L, Moyo SR, *et al.* Hospital-based study of sexually transmitted diseases at Murewa Rural district hospital, Zimbabwe 1991-1992. *Sex Transm Dis* 1995; **22**: 1–6.
90 Ghys PD, Diallo MO, Ettiegne-Traore V, *et al.* Genital ulcers associated with human immunodeficiency virus-related immunosuppression in female sex workers in Abidjan, Ivory Coast. *J Infect Dis* 1995; **172**: 1371–4.
91 Meless H, Abegaze B. Drug susceptibility of Neisseria isolates from patients attending clinics for sexually transmitted diseases in Addis Ababa. *East Afr Med J* 1995; **74**: 447–9.
92 Harms G, Matull R, Randrianasolo D, *et al.* Pattern of sexually transmitted diseases in a Malagasy population. *Sex Transm Dis* 1994; **21**: 315–20.
93 Dallabetta G, Behets F, Lule G, *et al.* Specificity of dysuria and discharge complaints and presence of urethritis in male patients attending an STD clinic in Malawi. *Sex Transm Infect* 1998; **74**: S34-7.
94 Behets FM, Liomba G, Lule G, *et al.* Sexually transmitted diseases and human immunodeficiency virus control in Malawi: a field study of genital ulcer disease. *J Infect Dis* 1995; **171**: 451–5.
95 Cossa HA, Gloyd S, Vaz RG, *et al.* Syphilis and HIV infection among displaced pregnant women in rural Mozambique. *Int J STD AIDS* 1994; **5**: 117–23.
96 Alary M, Baganizi E, Guedeme A, *et al.* Evaluation of clinical algorithms for the diagnosis of gonococcal and chlamydial infections among men with urethral discharge or dysuria and women with vaginal discharge in Benin. *Sex Transm Infect* 1998; **74**: S44–9.
97 Thomas T, Choudhri S, Kariuki C, Moses S. Identifying cervical infection among pregnant women in Nairobi, Kenya: limitations of risk assessment and symptom-based approaches. *Genitourin Med* 1996; **72**: 334–8.
98 King R, Choudhri SH, Nasio J, *et al.* Clinical and in situ cellular responses to Haemophilus ducreyi in the presence or absence of HIV infection. *Int J STD AIDS* 1998; **9**: 531–6.
99 Htun Y, Morse SA, Dangor Y, *et al.* Comparison of clinically directed, disease specific, and syndromic protocols for the management of genital ulcer disease in Lesotho. *Sex Transm Infect* 1998; **74**: S23-8.
100 Morse SA, Trees DL, Htun Y, *et al.* Comparison of clinical diagnosis and standard laboratory and molecular methods for the diagnosis of genital ulcer disease in Lesotho: association with human immunodeficiency virus infection. *J Infect Dis* 1997; **175**: 583–9.
101 Lo BB, Philippon M, Cunin P, Meynard D, TandiaDiagana M. Sexually transmitted diseases agents of genital discharges in Nouakchott, Mauritania. *Bull la Société Pathol Exot* 1997; **90**: 81–2.
102 Chen CY, Ballard RC, Beck-Sague CM, *et al.* Human immunodeficiency virus infection and genital ulcer disease in South Africa: the herpetic connection. *Sex Transm Dis* 2000; **27**: 21–9.
103 Schneider H, Coetzee DJ, Fehler HG, *et al.* Screening for sexually transmitted diseases in rural South African women. *Sex Transm Infect* 1998; **74**: S147-52.
104 Pillay DG, Hoosen AA, Vezi B, Moodley C. Diagnosis of Trichomonas vaginalis in male urethritis. *Trop Geogr Med* 1994; **46**: 44–5.
105 Mayaud P, Msuya W, Todd J, *et al.* STD rapid assessment in Rwandan refugee camps in Tanzania. *Genitourin Med* 1997; **73**: 33–8.
106 Aseffa A, Ishak A, Stevens R, *et al.* Prevalence of HIV, syphilis and genital chlamydial infection among women in north-west Ethiopia. *Epidemiol Infect* 1998; **120**: 171–7.
107 MacDonald KS, Malonza I, Chen DK, *et al.* Vitamin A and risk of HIV-1 seroconversion among Kenyan men with genital ulcers. *AIDS* 2001; **15**: 635–9.
108 Jackson DJ, Rakwar JP, Chohan B, *et al.* Urethral infection in a workplace population of East African men: evaluation of strategies for screening and management. *J Infect Dis* 1997; **175**: 833–8.
109 Gateau T, Zeller HG. Epidemiological approach for sexually transmitted diseases in Antsiranana (north Madagascar). Between prevention and treatment, the choice of a strategy against sexually transmitted diseases. *Arch Inst Pasteur Madagascar* 1996; **63**: 8–11.
110 Bakare RA, Oni AA, Umar US, *et al.* Non-gonococcal urethritis due to Chlamydia trachomatis: the Ibadan experience. *Afr J Med Med Sci* 2002; **31**: 17–20.
111 Ramuthaga TN, Mahomed FM, Greeff AS, Crewe-Brown HH, Vermeulen R. Comparison of urine with urethral swabs for the detection of Chlamydia trachomatis in men attending an STD clinic. *South African Med J* 1995; **85**: 1287–9.
112 Ahmed HJ, Borrelli S, Jonasson J, *et al.* Monoclonal antibodies against Haemophilus ducreyi lipooligosaccharide and their diagnostic usefulness. *Eur J Clin Microbiol Infect Dis* 1995; **14**: 892–8.
113 Latif AS, Mason PR, Marowa E, Gwanzura L, Chingono A, Mbengeranwa OL. Risk factors for gonococcal and chlamydial cervical infection in pregnant and non-pregnant women in Zimbabwe. *Cent Afr J Med* 1999; **45**: 252–8.

114 Farina C, Malighetti V, Lombart JP, *et al.* Yeasts from vaginal exudates in the Comoros islands. *J Mycol Med* 2000; **10**: 91–3.
115 Malonza IM, Tyndall MW, Ndinya-Achola JO, *et al.* A randomized, double-blind, placebo-controlled trial of single-dose ciprofloxacin versus erythromycin for the treatment of chancroid in Nairobi, Kenya. *J Infect Dis* 1999; **180**: 1886–93.
116 Cohen MS, Hoffman IF, Royce RA, *et al.* Reduction of concentration of HIV-1 in semen after treatment of urethritis: implications for prevention of sexual transmission of HIV-1. AIDSCAP Malawi Research Group. *Lancet* 1997; **349**: 1868–73.
117 Dieye AM, Samb NGD, Ba A, *et al.* Effectiveness of syndromic approach for management of uretral discharge in Senegal. *Med Trop* 2003; **63**: 45–8.
118 Lai W, Chen CY, Morse SA, *et al.* Increasing relative prevalence of HSV-2 infection among men with genital ulcers from a mining community in South Africa. *Sex Transm Infect* 2003; **79**: 202–7.
119 Ballard RC, Ye H, Matta A, Dangor Y, Radebe F. Treatment of chancroid with azithromycin. *Int J STD AIDS* 1996; **7**: S9-12.
120 Pepin J, Sobela F, Deslandes S, *et al.* Etiology of urethral discharge in West Africa: the role of Mycoplasma genitalium and Trichomonas vaginalis. *Bull World Health Organ* 2001; **79**: 118–26.
121 Morency P, Dubois MJ, Gresenguet G, *et al.* Aetiology of urethral discharge in Bangui, Central African Republic. *Sex Transm Infect* 2001; **77**: 125–9.
122 Gomes JP, Tavira L, Exposto F, Prieto E, Catry MA. Neisseria gonorrhoeae and Chlamydia trachomatis infections in patients attending STD and family planning clinics in Bissau, Guinea-Bissau. *Acta Trop* 2001; **80**: 261–4.
123 Fonck K, Mwai C, Rakwar J, Kirui P, Ndinya-Achola JO, Temmerman M. Healthcare-seeking behavior and sexual behavior of patients with sexually transmitted diseases in Nairobi, Kenya. *Sex Transm Dis* 2001; **28**: 367–71.
124 Ballard RC, Koornhof HJ, Chen CY, Radebe F, Fehler HG, Htun Y. The influence of concomitant HIV infection on the serological diagnosis of primary syphilis in southern Africa. *South African Med J* 2007; **97**: 1151–4.
125 Behets FM, Andriamiadana J, Randrianasolo D, *et al.* Chancroid, primary syphilis, genital herpes, and lymphogranuloma venereum in Antananarivo, Madagascar. *J Infect Dis* 1999; **180**: 1382–5.
126 Rajagopal M, Hoosen AA, Moodley J, Kharsany ABM, Moodley P, Sturm AW. Analysis of genital tract infections at a dedicated sexually transmitted diseases clinic. *South African J Epidemiol Infect* 1999; **14**: 77–82.
127 Hoosen AA, Moodley J, Maitin P, Sturm AW. Bacterial vaginosis in symptomatic women attending a gynaecology outpatient clinic. *South African J Epidemiol Infect* 1997; **12**: 119–21.
128 Ramjee G, Karim SS, Sturm AW. Sexually transmitted infections among sex workers in KwaZulu-Natal, South Africa. *Sex Transm Dis* 1998; **25**: 346–9.
129 Nzila N, Laga M, Thiam MA, *et al.* HIV and other sexually transmitted diseases among female prostitutes in Kinshasa. *AIDS* 1991; **5**: 715–21.
130 Anorlu RI, Fagbenro Beyioku AF, Fagorala T, Abudu OO, Galadanci HS. Prevalence of trichomonas vaginalis in patients with vaginal discharge in Lagos, Nigeria. *Niger Postgrad Median J* 2001; **8**: 183–6.
131 Ballard RC, Fehler HG, Htun Y, Radebe F, Jensen JS, Taylor-Robinson D. Coexistence of urethritis with genital ulcer disease in South Africa: influence on provision of syndromic management. *Sex Transm Infect* 2002; **78**: 274–7.
132 Obasi AI, Balira R, Todd J, *et al.* Prevalence of HIV and Chlamydia trachomatis infection in 15--19-year olds in rural Tanzania. *Trop Med Int Heal* 2001; **6**: 517–25.
133 Msuya SE, Mbizvo EM, Stray-Pedersen B, Sundby J, Sam NE, Hussain A. Risk assessment at the primary health care level in Moshi, Tanzania: limits in predicting sexually transmitted infections among women. *Cent Afr J Med* 2006; **52**: 97–104.
134 Msuya SE, Mbizvo E, Stray-Pedersen B, Sundby J, Sam NE, Hussain A. Reproductive tract infections among women attending primary health care facilities in Moshi, Tanzania. *East Afr Med J* 2002; **79**: 16–21.
135 Hoyo C, Hoffman I, Moser BK, *et al.* Improving the accuracy of syndromic diagnosis of genital ulcer disease in Malawi. *Sex Transm Dis* 2005; **32**: 231–7.
136 Chalamilla G, Mbwana J, Mhalu F, *et al.* Patterns of sexually transmitted infections in adolescents and youth in Dar es Salaam, Tanzania. *BMC Infect Dis* 2006; **6**: 22.
137 Ahmed HJ, Mbwana J, Gunnarsson E, *et al.* Etiology of genital ulcer disease and association with human immunodeficiency virus infection in two tanzanian cities. *Sex Transm Dis* 2003; **30**: 114–9.
138 Mbizvo ME, Musya SE, Stray-Pedersen B, Chirenje Z, Hussain A. Bacterial vaginosis and intravaginal practices: association with HIV. *Cent African Med J* 2004; **50**: 41–6.
139 Vuylsteke BL, Ettiegne-Traore V, Anoma CK, *et al.* Assessment of the validity of and adherence to sexually transmitted infection algorithms at a female sex worker clinic in Abidjan, Cote d'Ivoire. *Sex Transm Dis* 2003; **30**: 284–91.
140 Price MA, Miller WC, Kaydos-Daniels SC, *et al.* Trichomoniasis in men and HIV infection: data from 2 outpatient clinics at Lilongwe Central Hospital, Malawi. *J Infect Dis* 2004; **190**: 1448–55.

141 Anorlu R, Imosemi D, Odunukwe N, Abudu O, Otuonye M. Prevalence of HIV among Women with Vaginal Discharge in a Gynecological Clinic. *J Natl Med Assoc* 2004; **96**: 367–71.
142 Bakare RA, Oni AA, Umar US, Kehinde AO, Fayemiwo SA, Fasina NA. Ureaplasma urealyticum as a cause of non-gonococcal urethritis: the Ibadan experience. *Niger Postgrad Med J* 2002; **9**: 140–5.
143 Moodley P, Wilkinson D, Connolly C, Moodley J, Sturm AW. Trichomonas vaginalis is associated with pelvic inflammatory disease in women infected with human immunodeficiency virus. *Clin Infect Dis* 2002; **34**: 519–22.
144 Nilsen A, Kasubi MJ, Mohn SC, Mwakagile D, Langeland N, Haarr L. Herpes simplex virus infection and genital ulcer disease among patients with sexually transmitted infections in Dar es Salaam, Tanzania. *Acta Derm Venereol* 2007; **87**: 355–9.
145 Riedner G, Todd J, Rusizoka M, *et al.* Possible reasons for an increase in the proportion of genital ulcers due to herpes simplex virus from a cohort of female bar workers in Tanzania. *Sex Transm Infect* 2007; **83**: 91–6.
146 Watson-Jones D, Mugeye K, Mayaud P, *et al.* High prevalence of trichomoniasis in rural men in Mwanza, Tanzania: results from a population based study. *Sex Transm Infect* 2000; **76**: 355–62.
147 Pickering JM, Whitworth JA, Hughes P, *et al.* Aetiology of sexually transmitted infections and response to syndromic treatment in southwest Uganda. *Sex Transm Infect* 2005; **81**: 488–93.
148 Mason PR, Gwanzura L, Gregson S, Katzenstein DA. Chlamydia trachomatis in symptomatic and asymptomatic men: detection in urine by enzyme immunoassay. *Cent Afr J Med* 2000; **46**: 62–5.
149 Wolday D, Z GM, Mohammed Z, *et al.* Risk factors associated with failure of syndromic treatment of sexually transmitted diseases among women seeking primary care in Addis Ababa. *Sex Transm Infect* 2004; **80**: 392–4.
150 Pepin J, Deslandes S, Khonde N, *et al.* Low prevalence of cervical infections in women with vaginal discharge in west Africa: implications for syndromic management. *Sex Transm Infect* 2004; **80**: 230–5.
151 Romoren M, Velauthapillai M, Rahman M, Sundby J, Klouman E, Hjortdahl P. Trichomoniasis and bacterial vaginosis in pregnancy: inadequately managed with the syndromic approach. *Bull World Heal Organ* 2007; **85**: 297–304.
152 Bankole H, Faye-Kette H, Laruche G, Dabis F, Welffens-Ekra C, Dosso M. [Cell culture for active Chlamydia trachomatis infection in a population of symptomatic women in Abidjan]. *Bull la Société Pathol Exot* 2001; **94**: 235–8.
153 Tadesse A, Mekonnen A, Kassu A, Asmelash T. Antimicrobial sensitivity of Neisseria gonorrhoeae in Gondar, Ethiopia. *East Afr Med J* 2001; **78**: 259–61.
154 Zachariah R, Harries AD, Nkhoma W, *et al.* Behavioural characteristics, prevalence of Chlamydia trachomatis and antibiotic susceptibility of Neisseria gonorrhoeae in men with urethral discharge in Thyolo, Malawi. *Trans R Soc Trop Med Hyg* 2002; **96**: 232–5.
155 Ly F, Gueye N, Samb ND, Sow PS, Ndiaye B, Mahe A. [Prospective study of sexually transmitted infections in Dakar, Senegal]. *Médecine Trop* 2006; **66**: 64–8.
156 Moodley P, Sturm PDJ, Vanmali T, Wilkinson D, Connolly C, Sturm AW. Association between HIV-1 infection, the etiology of genital ulcer disease, and response to syndromic management. *Sex Transm Dis* 2003; **30**: 241–5.
157 Otuonye NM, Odunukwe NN, Idigbe EO, *et al.* Aetiological agents of vaginitis in Nigerian women. *Br J Biomed Sci* 2004; **61**: 175–8.
158 Namkinga LA, Matee MI, Kivaisi AK, Moshiro C. Prevalence and risk factors for vaginal candidiasis among women seeking primary care for genital infections in Dar es Salaam, Tanzania. *East Afr Med J* 2005; **82**: 138–43.
159 Mwansasu A, Mwakagile D, Haarr L, Langeland N. Detection of HSV-2 in genital ulcers from STD patients in Dar es Salaam, Tanzania. *J Clin Virol* 2002; **24**: 183–92.
160 Rassjo EB, Kambugu F, Tumwesigye MN, Tenywa T, Darj E. Prevalence of sexually transmitted infections among adolescents in Kampala, Uganda, and theoretical models for improving syndromic management. *J Adolesc Heal* 2006; **38**: 213–21.
161 Fayemiwo SA, Odaibo GN, Oni AA, Ajayi AA, Bakare RA, Olaleye DO. Genital ulcer diseases among HIV-infected female commercial sex workers in Ibadan, Nigeria. *Afr J Med Med Sci* 2011; **40**: 39–46.
162 Msuya SE, Uriyo J, Stray-Pedersen B, Sam NE, Mbizvo EM. The effectiveness of a syndromic approach in managing vaginal infections among pregnant women in northern Tanzania. *East Afr J Public Health* 2009; **6**: 263–7.
163 O'Farrell N, Morison L, Moodley P, *et al.* Genital ulcers and concomitant complaints in men attending a sexually transmitted infections clinic: implications for sexually transmitted infections management. *Sex Transm Dis* 2008; **35**: 545–9.
164 Weiss HA, Paz Bailey G, Phiri S, *et al.* Episodic therapy for genital herpes in sub-saharan Africa: a pooled analysis from three randomized controlled trials. *PLoS One* 2011; **6**: e22601.
165 LeGoff J, Mayaud P, Gresenguet G, *et al.* Performance of HerpeSelect and Kalon assays in detection of antibodies to herpes simplex virus type 2. *J Clin Microbiol* 2008; **46**: 1914–8.
166 Abauleth R, Boni S, Kouassi-Mbengue A, Konan J, Deza S. [Causation and treatment of infectious

leucorrhoea at the Cocody University Hospital (Abidjan, Cote d'Ivoire)]. *Sante* 2006; **16**: 191–5.
167 Mehta SD, Moses S, Parker CB, Agot K, Maclean I, Bailey RC. Circumcision status and incident herpes simplex virus type 2 infection, genital ulcer disease, and HIV infection. *AIDS* 2012; **26**: 1141–9.
168 Onyekonwu CL, Olumide YM, Oresanya FA, Onyekonwu GC. Vaginal discharge: aetiological agents and evaluation of syndromic management in Lagos. *Niger J Med* 2011; **20**: 155–62.
169 Suntoke TR, Hardick A, Tobian AA, *et al.* Evaluation of multiplex real-time PCR for detection of Haemophilus ducreyi, Treponema pallidum, herpes simplex virus type 1 and 2 in the diagnosis of genital ulcer disease in the Rakai District, Uganda. *Sex Transm Infect* 2009; **85**: 97–101.
170 Pepin J, Sobela F, Khonde N, *et al.* The syndromic management of vaginal discharge using single-dose treatments: a randomized controlled trial in West Africa. *Bull World Heal Organ* 2006; **84**: 729–38.
171 Pepin J, Deslandes S, Giroux G, *et al.* The complex vaginal flora of West African women with bacterial vaginosis. *PLoS One* 2011; **6**: e25082.
172 Phiri S, Zadrozny S, Weiss HA, *et al.* Etiology of genital ulcer disease and association with HIV infection in Malawi. *Sex Transm Dis* 2013; **40**: 923–8.
173 Brankin AE, Tobian AA, Laeyendecker O, *et al.* Aetiology of genital ulcer disease in female partners of male participants in a circumcision trial in Uganda. *Int J STD AIDS* 2009; **20**: 650–1.
174 Zimba TF, Apalata T, Sturm WA, Moodley P. Aetiology of sexually transmitted infections in Maputo, Mozambique. *J Infect Dev Ctries* 2011; **5**: 41–7.
175 Paz-Bailey G, Sternberg M, Puren AJ, Steele L, Lewis DA. Determinants of HIV type 1 shedding from genital ulcers among men in South Africa. *Clin Infect Dis* 2010; **50**: 1060–7.
176 Paz-Bailey G, Sternberg M, Puren AJ, *et al.* Improvement in healing and reduction in HIV shedding with episodic acyclovir therapy as part of syndromic management among men: a randomized, controlled trial. *J Infect Dis* 2009; **200**: 1039–49.
177 Ojuromi OT, Oyibo WA, Tayo AO, *et al.* Reliance on microscopy in Trichomonas vaginalis diagnosis and its prevalence in females presenting with vaginal discharge in Lagos, Nigeria. *J Infect Dev Ctries* 2007; **1**: 210–3.
178 Nwadioha SI, Bako IA, Onwuezobe I, Egah DZ. Vaginal trichomoniasis among HIV patients attending primary health care centers of Jos, Nigeria. *Asian Pacific J Trop Dis* 2012; **2**: 337–41.
179 Nwadioha S, Egah D, Banwat E, Egesie J, Onwuezobe I. Prevalence of bacterial vaginosis and its risk factors in HIV/AIDS patients with abnormal vaginal discharge. *Asian Pacific J Trop Dis* 2011; **4**: 156–8.
180 Bochner AF, Baeten JM, Rustagi AS, *et al.* A cross-sectional analysis of Trichomonas vaginalis infection among heterosexual HIV-1 serodiscordant African couples. *Sex Transm Infect* 2017; **93**: 520–9.
181 Brown LB, Krysiak R, Kamanga G, *et al.* Neisseria gonorrhoeae antimicrobial susceptibility in Lilongwe, Malawi, 2007. *Sex Transm Dis* 2010; **37**: 169–72.
182 Nwadioha S, Egah D, Nwokedi E, Onwuezobe I. A study of female genital swabs in primary health care centres in Jos, Nigeria. *Asian Pacific J Trop Dis* 2011; **1**: 52–4.
183 Muvunyi CM, Hernandez TC. Prevalence of bacterial vaginosis in women with vaginal symptoms in South Province, Rwanda. *African J Clin Exp Microbiol* 2009; **10**. http://ajol.info/index.php/ajcem/article/view/43408 NS -.
184 Moodley P, Wilkinson D, Connolly C, Sturm AW. Influence of HIV-1 coinfection on effective management of abnormal vaginal discharge. *Sex Transm Dis* 2003; **30**: 1–5.
185 Le Roux MC, Ramoncha MR, Adam A, Hoosen AA. Aetiological agents of urethritis in symptomatic South African men attending a family practice. *Int J STD AIDS* 2010; **21**: 477–81.
186 Nwadioha S, Egesie JO, Emejuo H, Iheanacho E. A study of female genital swabs in a Nigerian Tertiary Hospital. *Asian Pac J Trop Med* 2010; **3**: 577–9.
187 Nwadioha S, Egesie JO, Emejuo H, Iheanacho E. Prevalence of pathogens of abnormal vaginal discharges in a Nigerian tertiary hospital. *Asian Pac J Trop Med* 2010; **3**: 483–5.
188 Djohan V, Angora KE, Vanga-Bosson AH, *et al.* [In vitro susceptibility of vaginal Candida albicans to antifungal drugs in Abidjan (Ivory Coast)]. *J Mycol Med* 2012; **22**: 129–33.
189 Vandenhoudt HM, Langat L, Menten J, *et al.* Prevalence of HIV and other sexually transmitted infections among female sex workers in Kisumu, Western Kenya, 1997 and 2008. *PLoS One* 2013; **8**: e54953.
190 Vandepitte J, Bukenya J, Hughes P, *et al.* Clinical characteristics associated with Mycoplasma genitalium infection among women at high risk of HIV and other STI in Uganda. *Sex Transm Dis* 2012; **39**: 487–91.
191 Vandepitte J, Bukenya J, Weiss HA, *et al.* HIV and other sexually transmitted infections in a cohort of women involved in high-risk sexual behavior in Kampala, Uganda. *Sex Transm Dis* 2011; **38**: 316–23.
192 Karou SD, Djigma F, Sagna T, *et al.* Antimicrobial resistance of abnormal vaginal discharges microorganisms in Ouagadougou, Burkina Faso. *Asian Pac J Trop Biomed* 2012; **2**: 294–7.
193 Tine RC, Sylla K, Ka R, *et al.* A Study of Trichomonas vaginalis infection and correlates in women with vaginal discharge referred at fann teaching hospital in Senegal. *J Parasitol Res* 2019; : 1–8.
194 Nwadioha SI, Nwokedi EO, Egesie J, Enejuo H. Vaginal candidiasis and its risk factors among women attending a Nigerian teaching hospital. *Niger Postgrad Med J* 2013; **20**: 20–3.
195 Okunlola MA, Stella-Maris ON, Tokzaka AA, Ojengbede Oladosu A. Study on Vaginitis Among Intrauterine Contraceptive Device Users in Ibadan, South-western Nigeria. *J Reprod Contracept* 2009; **20**: 247–55.

196 De Jongh M, Le Roux M, Adam A, Caliendo AM, Hoosen AA. Co-Infection with Neisseria gonorrhoeae, Chlamydia trachomatis and Trichomonas vaginalis in Symptomatic South African Men with Urethritis: Implications for Syndromic Management. *Open Trop Med J* 2009; **2**: 13–6.
197 Djomand G, Gao H, Singa B, *et al.* Genital infections and syndromic diagnosis among HIV-infected women in HIV care programmes in Kenya. *Int J STD AIDS* 2016; **27**: 19–24.
198 Singa B, Glick SN, Bock N, *et al.* Sexually transmitted infections among HIV-infected adults in HIV care programs in Kenya: a national sample of HIV clinics. *Sex Transm Dis* 2013; **40**: 148–53.
199 De Jongh M, Lekalakala MR, Le Roux M, Hoosen AA. Risk of having a sexually transmitted infection in women presenting at a termination of pregnancy clinic in Pretoria, South Africa. *J Obstet Gynaecol (Lahore)* 2010; **30**: 480–3.
200 Makasa M, Buve A, Sandøy IF. Etiologic pattern of genital ulcers in Lusaka, Zambia: Has chancroid been eliminated? *Sex Transm Dis* 2012; **39**: 787–91.
201 Ibrahim SM, Bukar M, Galadima GB, Audu BM, Ibrahim HA. Prevalence of bacterial vaginosis in pregnant women in Maiduguri, North-Eastern Nigeria. *Niger J Clin Pract* 2014; **17**: 154–8.
202 Ibrahim SM, Mohammed B, Yahaya M, Audu BM, Ibrahim HA. Prevalence of vaginal candidiasis among pregnant women with abnormal vaginal discharge in Maiduguri. *Niger J Med* 2013; **22**: 138–42.
203 Konate A, Yavo W, Kassi FK, *et al.* Aetiologies and contributing factors of vulvovaginal candidiasis in Abidjan (Cote d'Ivoire). *J Mycol Med* 2014; **24**: 93–9.
204 Takuva S, Mugurungi O, Mutsvangwa J, *et al.* Etiology and antimicrobial susceptibility of pathogens responsible for urethral discharge among men in Harare, Zimbabwe. *Sex Transm Dis* 2014; **41**: 713–7.
205 van der Eem L, Dubbink JH, Struthers HE, *et al.* Evaluation of syndromic management guidelines for treatment of sexually transmitted infections in South African women. *Trop Med Int Heal* 2016; **21**: 1138–46.
206 Mulu W, Abera B, Yimer M, Hailu T, Ayele H, Abate D. Bacterial agents and antibiotic resistance profiles of infections from different sites that occurred among patients at Debre Markos Referral Hospital, Ethiopia: a cross-sectional study. *BMC Res Notes* 2017; **10**: 254.
207 Kerubo E, Laserson KF, Otecko N, *et al.* Prevalence of reproductive tract infections and the predictive value of girls' symptom-based reporting: findings from a cross-sectional survey in rural western Kenya. *Sex Transm Infect* 2016; **92**: 251–6.
208 Tine RC, Dia L, Sylla K, Sow D, Lelo S, Ndour CT. Trichomonas vaginalis and Mycoplasma infections among women with vaginal discharge at Fann teaching hospital in Senegal. *Trop Parasitol* 2019; **9**: 45–53.
209 National Institute for Communicable Diseases. Database of aetiological surveillance of vaginal discharge, urethral discharge and genital ulcer syndromes in South Africa. 2023.
210 Nkwabong E, Dingom MAN. Acute pelvic inflammatory disease in Cameroon: a cross sectional descriptive study. *Afr J Reprod Health* 2015; **19**: 87–91.
211 Fentaw S, Abubeker R, Asamene N, Assefa M, Bekele Y, Tigabu E. Antimicrobial susceptibility profile of Gonococcal isolates obtained from men presenting with urethral discharge in Addis Ababa, Ethiopia: Implications for national syndromic treatment guideline. *PLoS One* 2020; **15**: e0233753.
212 Dela H, Attram N, Behene E, *et al.* Risk factors associated with gonorrhea and chlamydia transmission in selected health facilities in Ghana. *BMC Infect Dis* 2019; **19**: 425.
213 Asmah RH, Agyeman RO, Obeng-Nkrumah N, *et al.* Trichomonas vaginalis infection and the diagnostic significance of detection tests among Ghanaian outpatients. *BMC Womens Heal* 2018; **18**: 206.
214 Rietmeijer CA, Mungati M, Kilmarx PH, *et al.* Serological markers for syphilis among persons presenting with syndromes associated with sexually transmitted infections: Results From the Zimbabwe STI etiology study. *Sex Transm Dis* 2019; **46**: 579–83.
215 Chirenje ZM, Dhibi N, Handsfield HH, *et al.* The etiology of vaginal discharge syndrome in Zimbabwe: Results from the Zimbabwe STI etiology study. *Sex Transm Dis* 2018; **45**: 422–8.
216 Kilmarx PH, Gonese E, Lewis DA, *et al.* HIV infection in patients with sexually transmitted infections in Zimbabwe: Results from the Zimbabwe STI etiology study. *PLoS One* 2018; **13**: e0198683.
217 Mungati M, Machiha A, Mugurungi O, *et al.* The etiology of genital ulcer disease and coinfections with Chlamydia trachomatis and Neisseria gonorrhoeae in Zimbabwe: Results from the Zimbabwe STI etiology study. *Sex Transm Dis* 2018; **45**: 61–8.
218 Rietmeijer CA, Mungati M, MacHiha A, *et al.* The etiology of male urethral discharge in Zimbabwe: Results from the Zimbabwe STI Etiology Study. *Sex Transm Dis* 2018; **45**: 56–60.
219 Konadu DG, Owusu-Ofori A, Yidana Z, *et al.* Prevalence of vulvovaginal candidiasis, bacterial vaginosis and trichomoniasis in pregnant women attending antenatal clinic in the middle belt of Ghana. *BMC Pregnancy Childbirth* 2019; **19**: 341.
220 Odunvbun WO. Prevalence of chlamydia infection using chlamydia antigen detection test and it's predictive value in tubal disease among infertile women attending the gynaecological clinic of the Eku Baptist Hospital, Eku, Delta State, South-South Nigeria. *Ann Biomed Sci* 2018; **17**. https://www.ajol.info/index.php/abs/article/view/169175 NS -.
221 Oyeyipo OO, Onasoga MF. Incidence and speciation of Candida species among non-gravid young females in Ilorin, North Central, Nigeria. *J Appl Sci Environ Manag* 2015; **19**: 680–5.
222 Marcotte H, Larsson PG, Andersen KK, *et al.* An exploratory pilot study evaluating the supplementation of

standard antibiotic therapy with probiotic lactobacilli in south African women with bacterial vaginosis. *BMC Infect Dis* 2019; **19**: 824.
223 Nabweyambo S, Kakaire O, Sowinski S, *et al.* Very low sensitivity of wet mount microscopy compared to PCR against culture in the diagnosis of vaginal trichomoniasis in Uganda: a cross sectional study. *BMC Res Notes* 2017; **10**: 259.
224 Sakala J, Chizuni N, Nzala S. A study on usefulness of a set of known risk factors in predicting maternal syphilis infections in three districts of Western Province, Zambia. *Pan Afr Med J* 2016; **24**: 75.
225 van Liere G, Kock MM, Radebe O, *et al.* High rate of repeat sexually transmitted diseases among men who have sex with men in South Africa: a prospective cohort study. *Sex Transm Dis* 2019; **46**: e105–7.
226 Latif AS, Gwanzura L, Machiha A, *et al.* Antimicrobial susceptibility in Neisseria gonorrhoeae isolates from five sentinel surveillance sites in Zimbabwe, 2015-2016. *Sex Transm Infect* 2018; **94**: 62–6.
227 Mehta SD, Pradhan AK, Green SJ, *et al.* Microbial Diversity of Genital Ulcers of HSV-2 Seropositive Women. *Sci Rep* 2017; **7**: 15475.
228 Gadoth A, Shannon CL, Hoff NA, *et al.* Prenatal chlamydial, gonococcal, and trichomonal screening in the Democratic Republic of Congo for case detection and management. *Int J STD AIDS* 2020; **31**: 221–9.
229 Chen JS, Matoga M, Khan S, *et al.* Estimating syphilis seroprevalence among patients in a sexually transmitted infections clinic in Lilongwe, Malawi. *Int J STD AIDS* 2020; **31**: 359–63.
230 Aduloju OP, Akintayo AA, Aduloju T. Prevalence of bacterial vaginosis in pregnancy in a tertiary health institution, south western Nigeria. *Pan Afr Med J* 2019; **33**: 9.
231 Kashibu E, Victor O, Ojumah I, Akafa R, Rikwentishe E. *Trichomonas vaginalis* infection: Prevalence and risk factors among ante-natal attendees in a tertiary facility in Taraba State, north-east Nigeria. *Niger J Parasitol* 2018; **39**: 199–203.
232 Verwijs MC, Agaba SK, Sumanyi JC, *et al.* Targeted point-of-care testing compared with syndromic management of urogenital infections in women (WISH): a cross-sectional screening and diagnostic accuracy study. *Lancet Infect Dis* 2019; **19**: 658–69.
233 Chabata S, Fearon E, Musemburi S, *et al.* Sexually transmitted infections among female sex workers in Zimbabwe - high and rising prevalenec blocks progress towards global elimination targets. *Unpublished* 2023.
234 Nacht C, Agingu W, Otieno F, Odhiambo F, Mehta SD. Antimicrobial resistance patterns in Neisseria gonorrhoeae among male clients of a sexually transmitted infections clinic in Kisumu, Kenya. *Int J STD AIDS* 2020; **31**: 46–52.
235 Wall KM, Nyombayire J, Parker R, *et al.* Etiologies of genital inflammation and ulceration in symptomatic Rwandan men and women responding to radio promotions of free screening and treatment services. *PLoS One* 2021; **16**: e0250044.
236 Happel AU, Singh R, Mitchev N, *et al.* Testing the regulatory framework in South Africa - a single-blind randomized pilot trial of commercial probiotic supplementation to standard therapy in women with bacterial vaginosis. *BMC Infect Dis* 2020; **20**: 491.
237 Connolly S, Wall KM, Parker R, *et al.* Sociodemographic factors and STIs associated with Chlamydia trachomatis and Neisseria gonorrhoeae infections in Zambian female sex workers and single mothers. *Int J STD AIDS* 2020; **31**: 364–74.
238 Mabaso N, Naicker C, Nyirenda M, Abbai N. Prevalence and risk factors for Trichomonas vaginalis infection in pregnant women in South Africa. *Int J STD AIDS* 2020; **31**: 351–8.
239 Melendez JH, Hardick J, Onzia A, *et al.* Retrospective analysis of Ugandan men with urethritis reveals Mycoplasma genitalium and associated macrolide resistance. *Microbiol Spectr* 2022; **10**: e0230421.
240 Zenebe MH, Mekonnen Z, Loha E, Padalko E. Prevalence, risk factors and association with delivery outcome of curable sexually transmitted infections among pregnant women in Southern Ethiopia. *PLoS One* 2021; **16**: e0248958.
241 Ahmed M, Admassu Ayana D, Abate D. Bacterial Vaginosis and associated factors among pregnant women attending antenatal care in Harar City, eastern Ethiopia. *Infect Drug Resist* 2022; **15**: 3077–86.
